# Non-medical COVID-19-related personal impact in medical ecological perspective: A global multileveled, mixed method study

**DOI:** 10.1101/2020.12.26.20248865

**Authors:** Timothy Dye, Brooke Levandowski, Shazia Siddiqi, José Pérez Ramos, Dongmei Li, Saloni Sharma, Erin Muir, Sophia Wiltse, Rebecca Royzer, Tiffany Panko, Wyatte Hall, Monica Barbosu, Carrie Irvine, Eva Pressman

**Affiliations:** University of Rochester, School of Medicine and Dentistry, Department of Obstetrics and Gynecology, Rochester, New York USA

## Abstract

**Background:** The COVID-19 pandemic has led to widespread public health measures to reduce transmission, morbidity, and mortality attributed to the SARS-CoV-2 virus. While much research and focus surrounds COVID-19 vaccine development, testing, and supportive management, little is known about the determinants of non-medical, personal impact of COVID-19 prevention policies. We aimed to understand determinants of non-medical COVID-19 impact and to account for its multileveled, intersectional nature of associations.

**Methods:** This cross-sectional, multi-level, convergent mixed-methods study assessed a range of beliefs, practices, and experiences relating to COVID-19. We recruited a global sample (n=7,411) using both Facebook and Amazon mTURK platforms. We constructed a novel data-driven non-medical COVID-19 Impact Score and four subcomponents (“Personal Action,” “Supply-related,” “Cancellations,” and “Livelihood” impacts). We used generalized estimating equation models with identity link functions to determine concomitant association of individual, household, and country-level variables on the impact scores. We also classified 20,015 qualitative excerpts from 6859 respondents using an 80-code codebook.

**Results:** Total and component impact scores varied significantly by region with Asia, Africa, and Latin America and the Caribbean observing the highest impact scores. Multilevel modeling indicated that individual-level sociocultural variables accounted for much of this variation with COVID-related worry, knowledge, struggles in accessing food and supplies, and worsening mental health most strongly associated with non-medical impact. Family responsibilities, personal COVID medical experience, and health locus of control – in addition to country-level variables reflecting social and health challenge – were also significantly and independently associated with non-medical impact.

**Discussion:** Non-medical personal impact of COVID-19 affects most people internationally, largely in response to shutdowns, implementing prevention requirements, and through economic consequences. In the context where most of the world’s population does not have direct medical experience with COVID-19, this phenomena of non-medical impact is profound, and likely impacts sustainability of public health interventions aimed at containing COVID-19.

## INTRODUCTION

The world has seen unprecedented cooperation in implementing prevention strategies and policies aimed at reducing the transmission and impact of SARS-CoV-2 infection, or COVID-19 (the resulting clinical disease).(1) People in their households and communities face challenges not only from the perceived and real threats of SARS-CoV-2 infecting them or their families, but additionally from state policies aimed at reducing the spread and impact of disease, including the closing of schools and workplaces, mandating masking and social-physical distancing, and limitations on congregation. Consequently, dramatic shifts in personal lives, households, and communities have extended beyond the medical experience (i.e., disease detection, treatment, follow-up) of COVID-19 extending into non-medical effects that impact macro and micro-economies, social interaction, work, and maintenance of household needs. Layered upon these personal responses to government policies are pre-existing inequities, pre-existing maldistribution of resources and supplies, complex personal and household dynamics, and beliefs – all of which when combined create differential impacts on personal, lived experience and non-medical dimensions of COVID-19 prevention.(2, 3)

Traditional approaches to infectious disease focus on individual level factors and preventive strategies such as vaccine development or social-physical distancing (primary), early and mass testing (secondary), and curative treatment (tertiary). Western medical practice prioritizes focus on the individual’s illness, detection, identification of risks, and prevention strategies. (4) The centrality of the individualized aspect of disease has shifted more recently to a public health approach, by determining economic and social factors and studying population characteristics. (5) Populations encompass several domains at the country, community, and cultural levels that frequently influence individual-level health outcomes. Focused attention on upstream determinants can affect health outcomes downstream. (6) These upstream country-level effects fall within three areas: social determinants of health, behavioral influences, and power dynamics shaping governing and socio-political actions.

The transmission rate of COVID-19 is characterized by its propensity to travel rapidly throughout the world and has globally transformed the lives of people. A pandemic of this scale was forewarned by international entities and several studies as early as 2007, foreshadowing an explosive combination of interaction between coronavirus, bats, mammals, and humans. (7) Even with numerous restrictions on travel, the number of COVID-19 cases had reached more than 2 million worldwide by April 2020. (8) The intense focus on controlling the spread of COVID-19 has called for social-physical distancing methods, avoidance of crowds, and respiratory and hand hygiene by the World Health Organization (WHO). (9) Not all countries have responded the same, however, to WHO mandates of preventing the spread of COVID-19. (10)

The WHO defines social determinants of health as “conditions in which people are born, grow, live, work and age, and which are shaped by the distribution of money, power and resources at global, national and local levels,” or, more broadly, to any non-medical factor that influences a person’s health. (6) Social determinants of health include socioeconomic and political factors such as a person’s income/wealth or poverty/economic instability; access to housing, education, health care, food, employment; stress; and race/ethnicity. Poverty and education have been well-documented to have strong interrelationships with poorer health outcomes. (11, 12) Many who have a lower socioeconomic status (SES) fear medical costs and loss of income if they test positive and are unable to quarantine for two weeks, or fear losing their job. (13) Mental health and stress coping skills act in complex, interdependent relationships with the other determinants. (14) For example, essential employees such as grocery store workers may feel stressed by their higher risk of exposure. (15) (9)

Defining how social determinants of health impact COVID-19 transmission is essential to incorporate into behavioral prevention efforts,,such as social-physical distancing. Social-physical distancing poses difficulty in many places of the world where people live in close quarters (16). When the first COVID-19 case was reported in China in a familial cluster, focused attention shifted to overcrowding, decreased sanitation, and poor living conditions, which showed an increased risk in transmission of COVID-19. (16) This prompted immediate social-physical isolation practices and policies. In low-middle income countries (LMIC), however, many live in overcrowded households and neighborhoods and find it impossible to isolate older adults or vulnerable people who live together. (16) People experiencing homelessness have a higher risk of infection as well, due to the inability to appropriately social-physical distance.

Cultural differences contribute to the effects of the social determinants of health. Some high-risk populations have strong religious beliefs about God protecting them from disease and do not comply with social-physical distancing practices. (17) Some countries emphasize religious gatherings and social cohesion that impede mitigation strategies. (18) However, when the Malaysian government ordered lockdown, 95% of the country’s population complied with the order within a week. (19) China, Malaysia, Taiwan, South Korea, and Singapore reacted quickly and collectively due to learnings from SARS in 2003 and H1N1 influenza in 2009. (19-21)

The most important domain of influence that drives most of the effects of an individual’s health outcomes is behavioral, that accounts for upwards of 40% of health effects. (22) Behavioral influence includes daily choices and coping strategies such as handwashing, using sanitizers, social-physical distancing, getting tested, and wearing facial masks. However, millions of people around the world do not have access to clean running water which makes frequent handwashing nearly impossible. (23) Psychological reactions (23) such as fear and anxiety can determine people’s behaviors and even more so when there are strong social norms and expectations around these behaviors. (24) People are driven by societal norms at the community level to conform to what they perceive as social approval and that determines their compliance to preventive measures. (25)

One of the most surprising and perplexing conundrums of this pandemic have been the varied responses of governments as power dynamics emerged in their handling of the COVID-19 pandemic. As COVID-19 was identified in China and the virus spread to high-income countries, many faced baffling strategies motivated by politics and economics. Countries that had plans were better prepared. South Korea and Taiwan prioritized prevention of COVID-19 by immediately starting testing people and implementing contact tracing methods to isolate asymptomatic individuals. (26, 27) In the UK, it was not until the Imperial College London released a report estimating 7 billion infections and 40 million deaths, that the UK government began t pursue mitigating efforts at the policy level. (28, 29) African countries started stepping up on their efforts as soon as the COVID-19 outbreak was declared a pandemic, acting on their experience of the 2013-2016 Ebola epidemic. (30)

Economic disruptions and ineffective government responses have influenced the community’s compliance in following public health and policy measures. Citizens want to feel protected and in some populations the mishandling of government responses has eroded their trust and mental well-being. Sweden relied on voluntary compliance, while most countries used legal measures (31) (Sweden, however, has one of the highest numbers of COVID-19 cases now (32)). Communities where government leaders resisted public health guidelines calling for wearing masks and social-physical distancing have increased the risk of spreading COVID-19. (33) Australians who reported more trust in governmental authorities reported a higher inclination to adopt social-physical distancing strategies. (34) This leads to the concept of governmental influence on how much its citizens trust government policies and thereby positively comply with preventive strategies. It therefore appears that trust in one’s government coupled with clear and measured guidance is important in managing the COVID-19 pandemic.(35)

Ultimately, the fate of an individual’s health is multi-disciplinary and -dimensional; the domains intersect and influence one’s susceptibility to become infected with COVID-19. Amid fear and uncertainty, social, behavioral, and power dynamics drive variations in COVID-19 outcomes at both the individual and country levels. As a theoretical framework, Critical Medical Ecology is a multilevel model that accounts for the intersectional experiences in which people live and are exposed to infections, potentially interact with the medical system, and experience clinical outcomes. Specifically, the model considers sociocultural, biological, health care, and abiotic circumstances at individual, household, and community levels and is well-suited for analysis of COVID-19 and multilevel modeling.(36, 37)

To account for the multileveled, intersectional nature of influences on non-medical COVID-19 impact – in addition to its emergent, unstructured conceptual influences –we designed and implemented a global mixed-methods study. The goal was to identify and to understand determinants and lived experience of non-medical impact resulting from COVID-19 policies, practices, and effects, using the Critical Medical Ecology framework (CMEF) as a heuristic device to organize concepts and interpret findings.

## MATERIALS AND METHODS

### Study Design

This cross-sectional, multi-level, convergent mixed-methods study assessed a range of beliefs, practices, and experiences relating to COVID-19, prevention efforts, and ancillary health and non-medical issues that may impact these beliefs, practices, and experiences. The overall paradigm guiding the design is the Critical Medical Ecological model (36) that accounts for the multilevel, multidimensional quantitative and qualitative data comprising this study (see Figure 1). The aim of the quantitative component is to explain contributions and associations of various medical ecological variables across multidimensional domains and levels of organization on COVID-19-related non-medical personal impact. The qualitative component’s aim is to identify themes and patterns of emic and etic constructs that provide explanatory insight regarding how unstructured variables relate to non-medical COVID-19 impact. Further, we were interested in a global comparative approach across both qualitative and quantitative data, integrating findings across six world regions (Africa, Asia, Europe, Latin America and the Caribbean, Northern America, and Oceania) to help identify potential variation by region of the world. We adhered to the Reporting of Studies Conducted using Observational Routinely-collected Data (RECORD) guidelines (38) and the Consolidated Criteria for Reporting Qualitative Research (COREQ)(39) reporting guidelines in the reporting of this study. Shown in Figure 1, quantitative and qualitative variables that were assessed for association with respondents’ non-medical COVID-19 Impact Score were applied to the Critical Medical Ecology framework previously described.(36) This model and conceptual structure of variables impacts the selection of analytic methods used to develop multilevel quantitative and qualitative explanatory models of non-medical COVID-19 impact.

**Figure 1:**
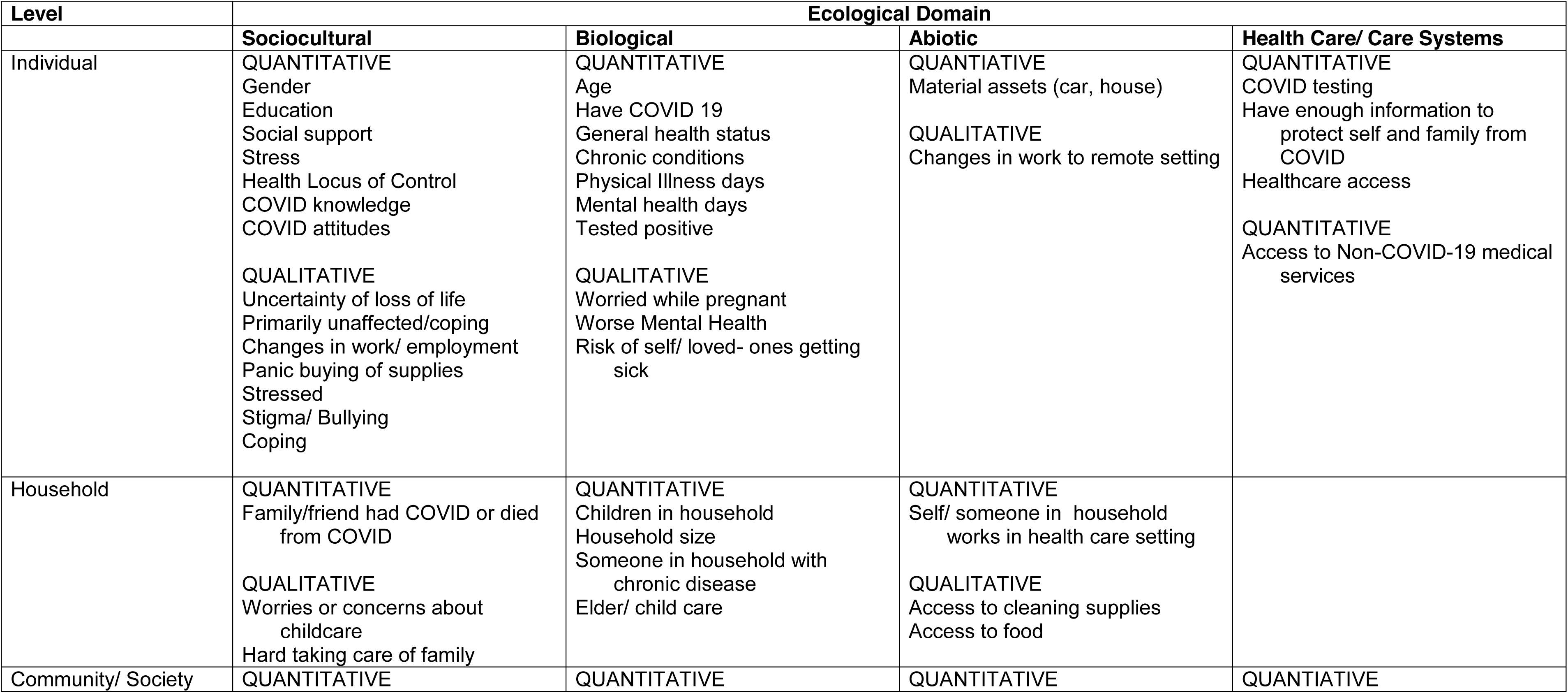
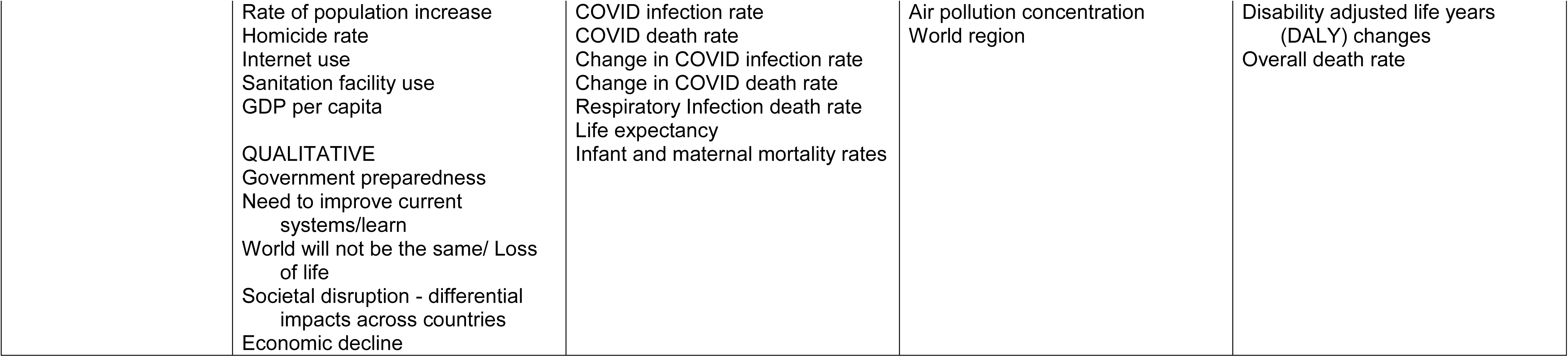
Critical Medical Ecological Framework for Global COVID-19 non-medical Impact Study, Integrating Both Quantitative and Qualitative Data Sources.

### Funding

Funding for this study was provided by a grant from the Richard & Mae Stone Goode Foundation, which was not involved in the design, collection, analyses, nor writing of this study.

### Setting

This study was worldwide in scope, with a survey developed and deployed in English, Spanish, French, and Italian through the University of Rochester’s REDCap system. At the time of this study, known COVID cases were most commonly found (see Dong et al. 2020) (52) in the USA (1,072,667), Spain (213,435), Italy (205,463), the UK (177,543), and France (165,764), hence the priority of conducting the survey in the major languages spoken in those regions of the world. Data collection occurred online during an eight-week period from April 6, 2020 to May 29, 2020.

### Sample Size

We decided *a priori* to recruit based on geographic region of the world using the United Nations M49 Standard Regional Codes(42) for Africa, Asia, Europe, Northern America, Latin America and the Caribbean, and Oceania to aggregate the International Standard Organization (ISO) 3166 Country Codes(43) of where respondents indicated they resided. Sample size was based on the standardized T-score computed for the major indices of this study, namely the primary outcome of interest: the non-medical-related COVID-19 Personal Impact Score. The effect size we aimed to detect was a mean difference of 3 points from the Impact Index mean of 50 (SD=10). Assuming confidence of 99.9%, power of 80.0%, and a 1:1 ratio of regions to one another, we estimated 380 individuals were required per geographic region. Additionally, we inflated sample size by 50% (to 570), minimally, to account for multivariate analysis, missing data, and planned sub-analyses. JMP Pro 14.1.0 (SAS Institute Inc., Cary, NC) was used to compute sample size. Recruitment occurred in all regions throughout the period of the survey until all regions had reached or exceeded the per-region inflated sample size target (570).

### Respondents

The final sample is presented in Figure 2, Global COVID Study Sample Disposition. Research respondents were recruited in all four languages through two global, multilingual mechanisms: 1) the Amazon Mechanical Turk (“mTURK” online workforce)(40) and 2) through Facebook, Instagram, and the Facebook Audience Network.(41) Facebook’s platforms have broad global reach and are frequently accessed for survey research.(41) mTURK’s platform provides access to a global workforce in multiple languages and provides a mechanism to reach reliable respondents around the world, including those with minimal use of or no access to Facebook platforms.(40) Participation was limited to individuals self-identifying as age 18 and older who could read English, Spanish, French, or Italian. Potential respondents through both mechanisms were directed to the REDCap survey in the language of recruitment (English, Spanish, French, Italian), were presented with the RSRB-approved Information Sheet in that language, provided consent to continue, and were asked to confirm their age and country of residence. Because of mTURK’s existence as a digital workforce, mTURK respondents were paid between $1.00 to $3.00 for completing the survey, depending on the difficulty in locating respondents. Facebook or Instagram respondents were not compensated. Respondents were recruited through multilingual (English, Spanish, French, Italian) advertisements on Facebook’s multiple platforms and the Amazon Mechanical Turk platform. The advertisements included a University of Rochester REDCap link for potential respondents to access the survey in multiple languages. Upon reaching the REDCap site (n=18,825), respondents were presented with an IRB-approved information sheet, and after having confirmed that they were 18 years old or older, asked if they consented to continue with the survey (n=15,638). Respondents were asked to self-identify their country of residence (n=11,757), the only required field in the survey. Overall 6,125 respondents reached the end of the survey and an additional 1,286 respondents completed the survey at least partially. In total, these 7,411 individuals form the base sample (2,969), or 40%, from the mTURK portal, and (4,442), or 60%, from the Facebook portals. Respondents from 173 countries were included in the final sample.

**Figure 2:**
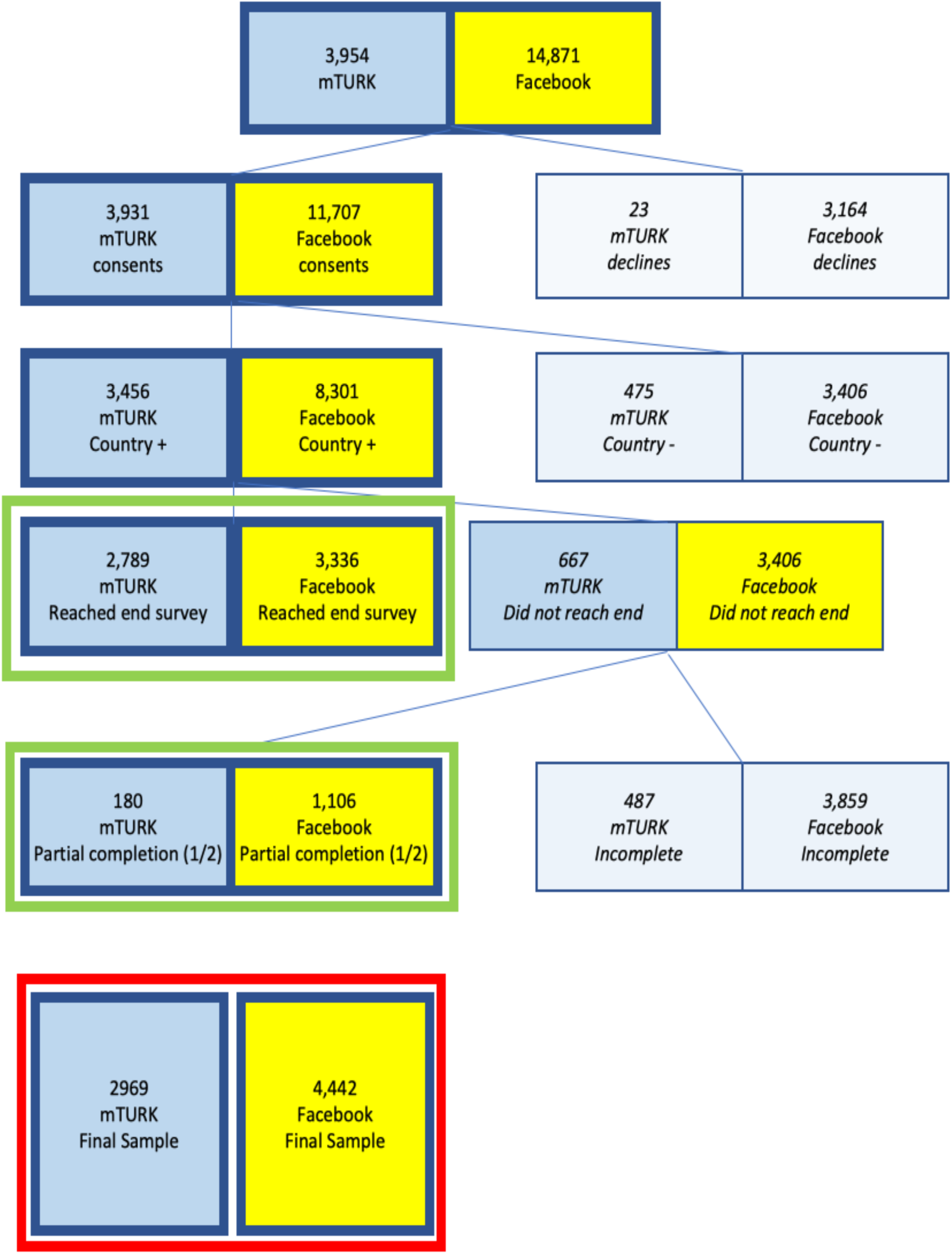
COVID Global Survey Sample Size Disposition.

### Measures

The Global COVID Survey included several standardized psychosocial indices including: the Multidimensional Health Locus of Control,(44) the Perceived Stress Scale,(45) and the Multidimensional Scale of Perceived Social Support.(46) Additionally, we incorporated elements of the CDC’s Health-Related Quality of Life 14 module(47) (HRQOL-14; 4-item Healthy Days Core Module, HRQOL– 4) and a standardized NHIS (National Health Interview Survey) question for assessing Health Access/Cost.(48) Most items for the coronavirus/ COVID-19 questions of the survey originated from the Kaiser Family Foundation (KFF)’s Coronavirus Poll(49) or were constructed and tested by the project team for flow and understandability. These COVID-19 questions ascertained knowledge, attitudes, and non-medical impact, and assembled into summary indices.

### Outcome: COVID-related non-medical Personal Impact Score and Subscores

We were guided by Streiner et al.’s method(50) for constructing the main outcome of this analysis: the COVID-related non-medical Personal Impact Score. Items assessed for inclusion were binary (yes/no) responses to the following question (items marked with (*) were added by the research team; all other items originated with the Kaiser Family Foundation, KFF Coronavirus Poll March 2020): “*Please tell us if you have taken any of the following actions because of the recent coronavirus outbreak*” (Decided not to travel or changed travel plans; Bought or worn a protective mask; Stocked up on items such as food and household supplies; Postponed or canceled health care visits;* Got extra refills on prescription medication;* Stayed home instead of going to work, school, or other regular activities; Postponed or canceled a medical procedure or surgery;* Canceled plans to attend large gatherings such as concerts or sporting events; Quit my job*) in addition to yes/no responses to “*Have you experienced any of the following because of coronavirus*?” (Lost income from a job or business; Been unable to get groceries; Been unable to get cleaning supplies or hand sanitizer; Been unable to get prescription medication; Have you or a family member been harassed, bullied, or hurt because of coronavirus*).

Items achieving 0.2 or higher corrected item-total correlation were retained in the Impact Score (all 14 items met this criteria) and the overall standardized Chronbach’s alpha for the resulting score was 0.65. To adequately account for completion of items, the final Impact Score constructed for analysis averaged individual participant responses (number of “yes” responses to items answered divided by the total number of items answered) for respondents answering at least half (7) of the individual impact items. That raw mean was then converted to a z-score and transformed to a standardized T-score with a mean of 50 and standard deviation of 10. The resulting score was the outcome measure for this study (the COVID-related non-medical Personal Impact T-Score). Additionally, we used Principal Components Analysis to cluster the fourteen individual impact items, resulting in four subscores of non-medical personal impact: Personal Actions, Supply-related, Cancellations and postponements, and Livelihood. As with the main Impact Scores, these subscores were computed using Streiner et al.’s approach,(50) and their standardized T-score values are used in analysis.

### Country-level Data

We included selected data at the country level regarding economic classifications from the World Bank, including income (High, High Middle, Low Middle, Low), development status (Least Developed Countries), and Fragile and Conflict-affected Situations; from the United Nations (Small Island Developing States);(51) and health, workforce, and environmental data from the WHO’s *World Health Statistics 2018: Monitoring health for the SDGs*.(52) Data regarding the COVID-19 pandemic at the country level is included from the COVID-19 Data Repository from the Johns Hopkins Center for Systems Science and Engineering,(53) which organizes data from WHO, national health surveillance organizations, universities, and other institutions tracking coronavirus metrics. COVID-19 data were included in analysis from the May 1, 2020 point-in-time (during data collection) and from August 1, 2020 (at the time of analysis). Data from the US Virgin Islands, Guam, American Samoa, Northern Marianas, and Puerto Rico were separated from the USA and reported separately for those jurisdictions.(54)

### Data Cleaning

All records were scanned (visually, and through algorithms) for aberrant and non-sensical responses, underage (less than 18 years old) or age missing, clearly fabricated response patterns, and for duplication. To minimize bias, a subcommittee from the research team was formed to review questionable records, discuss, and make decisions about deletion, correction, and inclusion. Any identifying details that respondents included in open-ended responses were deleted from the record.

### Statistical Analysis

Summary statistics and frequency distributions were used to characterize the demographics of respondents, households, and social norms, as well as the impact of COVID-19 on respondents. The differences in demographics and impact of COVID-19 across the six regions were examined based on 95% confidence intervals. For each impact outcome, Generalized Estimating Equation (GEE) models with identity link functions were used to examine the association of demographics of individuals, households, and social norms with personal impact score and subscores. Exchangeable variance-covariance structure was used to model the correlations between subjects coming from the same country. Purposeful model selection procedure was used to select important predictor variables in the GEE model. The Variance inflation factor (VIF) values were used to examine whether potential multicollinearity existed among predictor variables. Quasi-likelihood under the independence model criterion (QIC) values were used to examine the goodness-of-fit of the GEE models. All data analysis was conducted using the statistical analysis software SAS v9.4 (SAS Institute Inc., Cary, NC). The significance level of all tests was set at 5%.

The effect size of each predictor variable in the multivariable GEE models was estimated using the absolute value of the estimated coefficients divided by associated standard deviations, which is mathematically equivalent to the absolute value of the Z scores from the GEE models divided by the square root of the sample size used in the GEE model. The effect sizes were estimated for each predictor variable associated with each outcome variable in total samples, as well as samples in each region. Effect sizes of predictor variables in different levels and different dimensions were added together to denote the contributions of different levels and different dimensions to the variation of each outcome variable. Relative contributions were calculated using effect size of each predictor variable divided by the summed effect size of all predictor variables in the GEE model.

Missing data patterns were examined in the data, and missing at random pattern was found to be the possible missing mechanisms. To minimize the effect of missing data, predictor variables with over 10% of missing data and high correlations (VIF)>5) with other predictor variables were excluded during the model selection procedure. Maximum likelihood function estimates were used in parameter estimates to obtain unbiased estimates for associations of predictor variables with each outcome variable.

### Qualitative Coding

Respondent-level open-ended responses were uploaded to Dedoose (SocioCultural Research Consultants, Manhattan Beach, CA) and linked with key quantitative variables (including the Personal Impact Score and its subscores) as descriptors. All open-ended responses were translated into English prior to coding, and coding was conducted in English. A qualitative codebook was created based on review of the first 1,300 respondents to identify respondent-driven (emic) themes and also incorporating investigator-driven (etic) and theoretical priorities. A tiered codebook with 23 parent codes, 35 child codes, and 22 grandchild codes was finalized and applied to all final open-ended responses (see Supplementary Material). Three coders were engaged in the coding process, each completing a coding test of 400 randomly sampled qualitative excerpts and accomplishing moderate-high Kappa scores for each code and exceeding 90% agreement before initiating coding work. Coders were blinded to data descriptors (e.g., Impact Scores, region) when applying codes to excerpts.

We used JMP Pro 14.1.0 (SAS Institute, Cary, NC)’s Cluster Variables platform to reduce the 80 qualitative codes from this study to clusters of qualitative code variables, identified by the most representative variable (highest correlation) in the cluster.

### Mixed Method Integration

Using the building technique,(55) initial screening of all qualitative codes were conducted by level of the previously-described quantitative Impact Score. An initial listing of themes was explored through code co-occurrence, content review of coded excerpts, and cross-tabulation of data descriptors. Key thematic findings are presented generalizing these analyses and presenting original quotations from coded excerpts in their original language (along with translated English) to illustrate. Where feasible, the study’s ecological model guides presentation and interpretation of mixed methods results by level and dimension, integrating quantitative and qualitative results. Additional themes arising from this study that are not explicitly suited for presentation in a medical ecological framework are also identified as additional findings to explore.

### Ethical Review

This study was performed in accordance with the ethical standards established by the 1964 Declaration of Helsinki and its later amendments. The University of Rochester’s Research Subjects Review Board determined that this study met federal and University criteria for exemption. Respondents consented to participate in this research after a review of a detailed Information Sheet presented at the beginning of the REDCap survey and a confirmation of country of residence. Leaving any question they wished unanswered was allowed.

## RESULTS

This study included participants from 173 countries and a total participation of 7,411 individuals between Facebook and mTurk platforms.

### Demographics

#### Individual demographics

As shown in Table 1, most demographic characteristics of respondents varied across the six regions of this study. For example, more women participated in the study in Latin America and Oceania, while more men participated in Africa, Asia, and Europe. Latin America, Northern America, and Oceania generally had a higher proportion of older respondents while Africa, Asia, and Europe typically had more younger respondents. In all regions, more than half of all respondents who participated had attended at least some college. Missing demographic data included age from 16.9% of respondents and gender from 17.9% of respondents, with the highest missing data rate for age and gender in Africa (33.4% and 36.0%, respectively) and the lowest missing data rate on age and gender in Europe (8.9% and 9.4%, respectively).

**Table 1:**
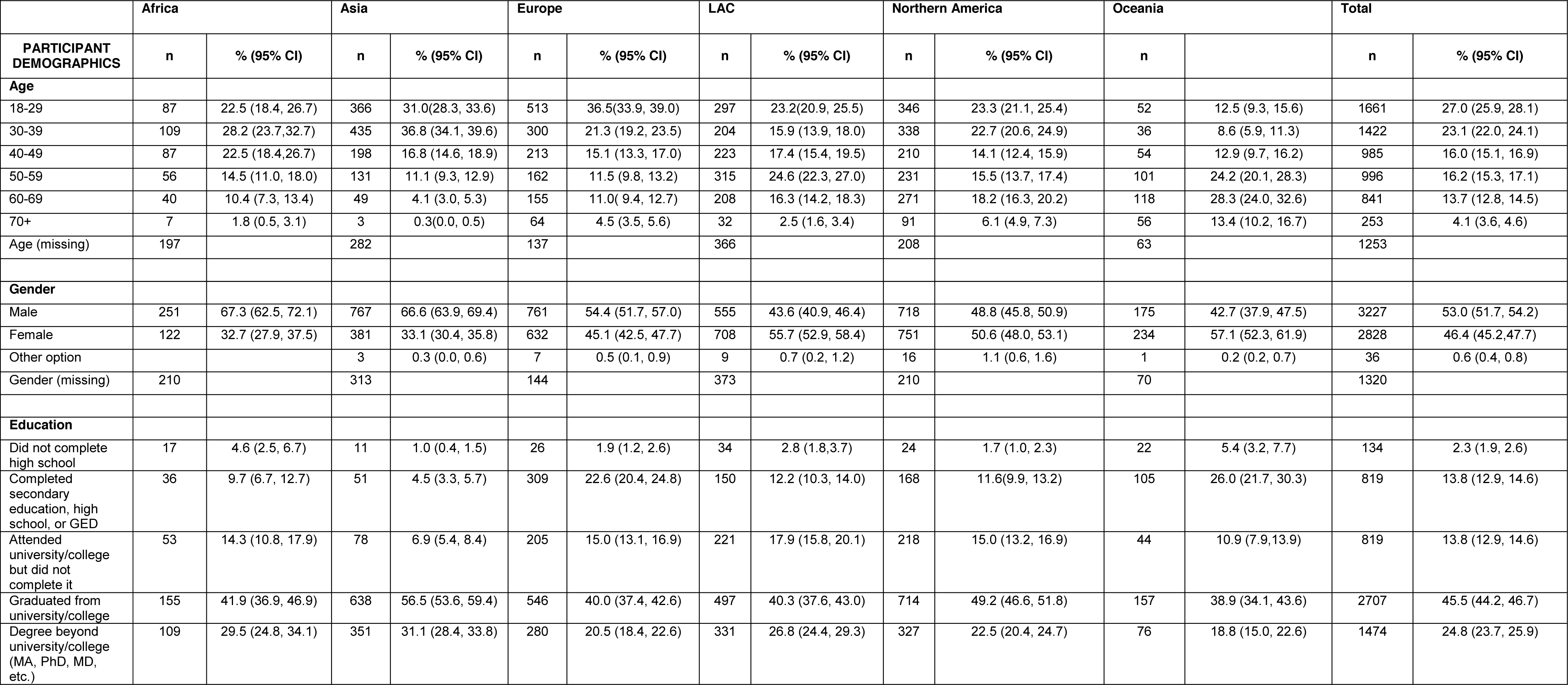
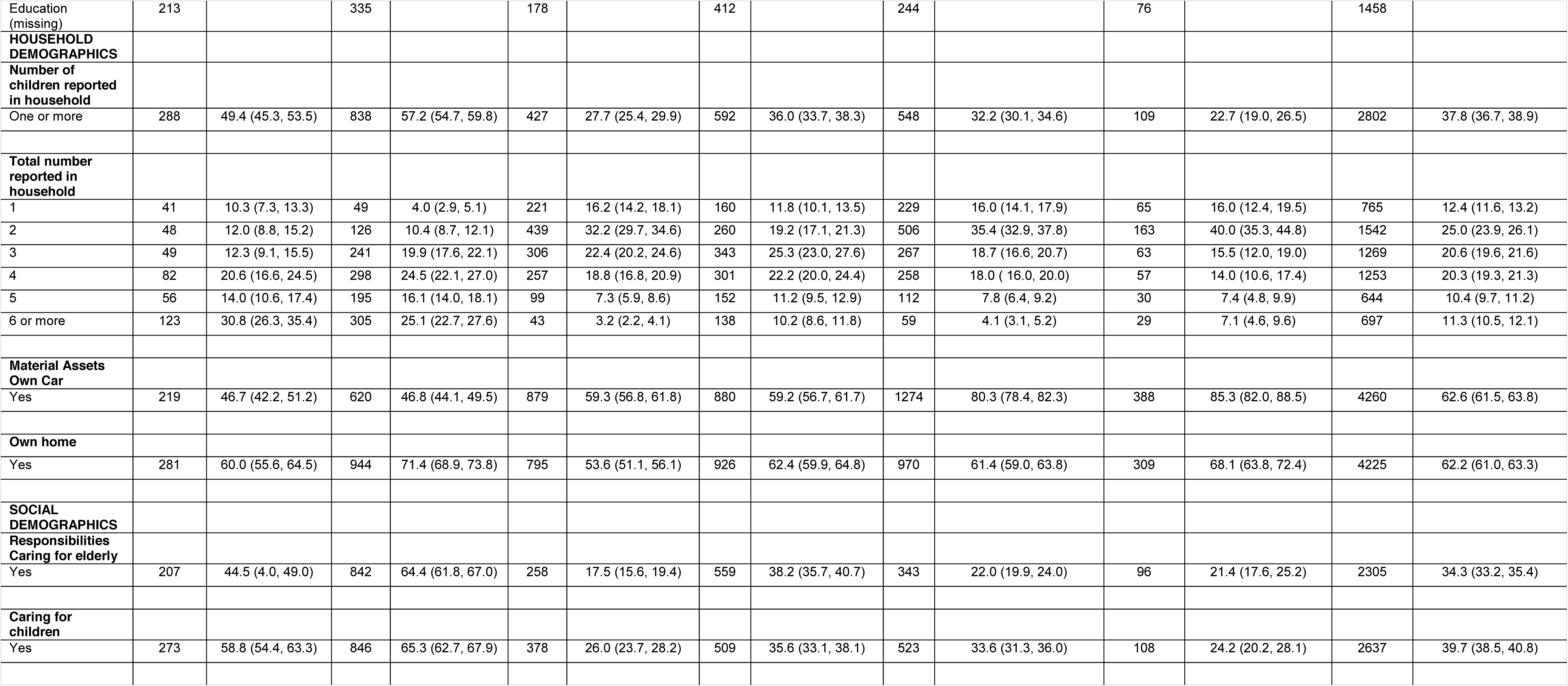
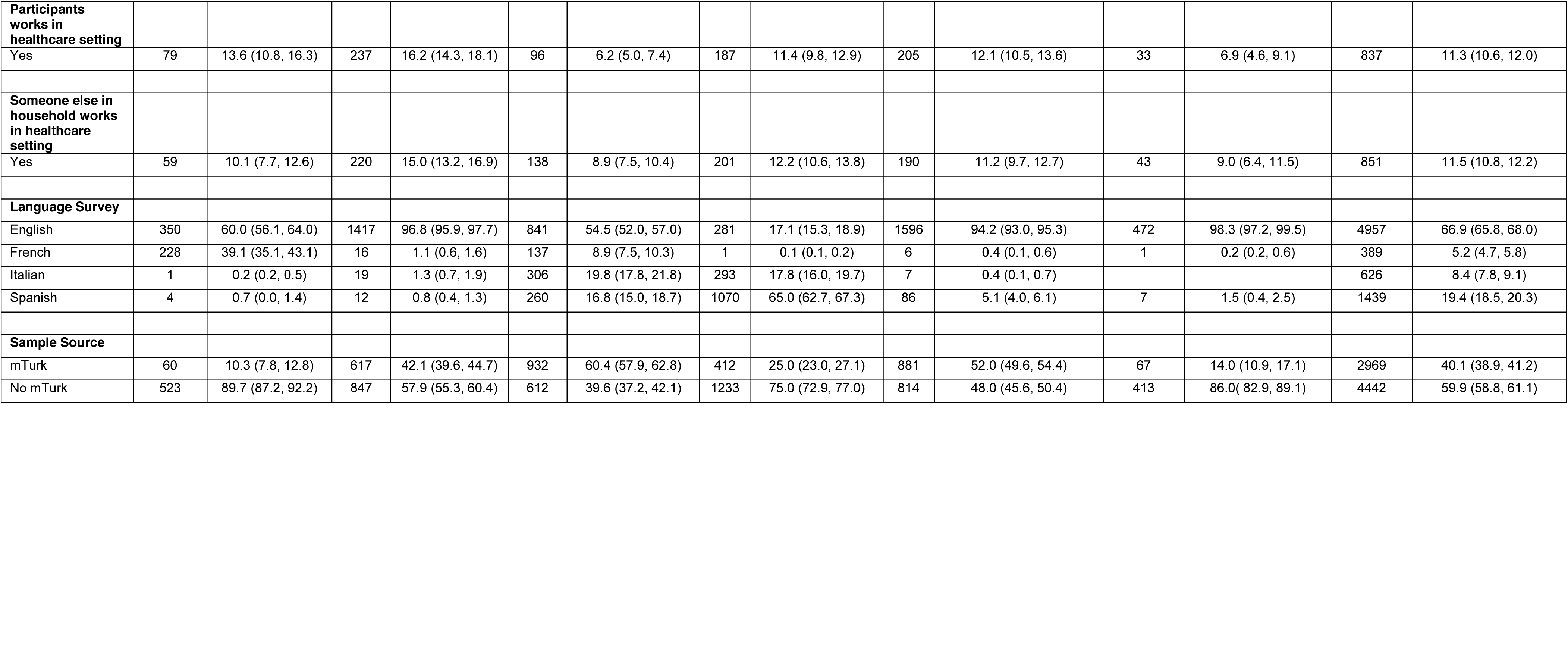
Global COVID Study participants demographic, household, and social characteristics, by UN region code.

#### Household demographics

Overall, 37.8% of respondents indicated they had one or more children under age 18 in their home, with respondents in Asia most likely to report having one or more children in the home (57.2%) and with Oceania being the least likely to report children in the home (22.7%). Household sizes were largest (6 or more members) in Africa (30.8%) and in Asia (25.1%), and were smallest in Oceania (56% with one or two in household) and Northern America (51.4%). In each region, more than half of respondents indicated they owned their own home, and in Latin America, Northern America, Europe, and Oceania, most respondents indicated they owned their own car.

#### Social demographics

In total, 34.3% of respondents indicated they had responsibilities for caring for elderly people, most commonly in Asia (64.4%), Africa (44.5%) and in Latin America and the Caribbean (38.2%). Similarly, respondents in Asia (65.3%) and in Africa (58.8%) were more likely to indicate they had responsibilities providing care for children. Respondents from Asia were more likely than respondents from other regions to indicate they (16.2%) or someone in their household (15.0%) worked in a healthcare setting. Most respondents (66.9%) completed the survey in English. In Latin America and the Caribbean, 82.8% of respondents completed the survey in Spanish or Italian, in Europe 45.5% completed the survey in Spanish, Italian, or French, and in Africa 39.1% of respondents completed the survey in French. Most respondents were reached through Facebook (59.9%) especially in Africa (89.7%), Oceania (86.0%), and in Latin America and the Caribbean (75%).

### COVID-19 Experience

Table 2 presents a variety of COVID-19-related experiences by region. Respondents (with children under 18) in Africa, Asia, and Latin America were more likely to report closure of daycare and schools for their children as a result of COVID-19 compared to similar parents in Europe, Northern America, and Oceania. About half of all respondents with children indicated they had difficulty with alternate arrangements for childcare, particularly in Africa and in Asia. Similar proportions of respondents indicated they would be able to do at least part of their job from home if required.

**Table 2:**
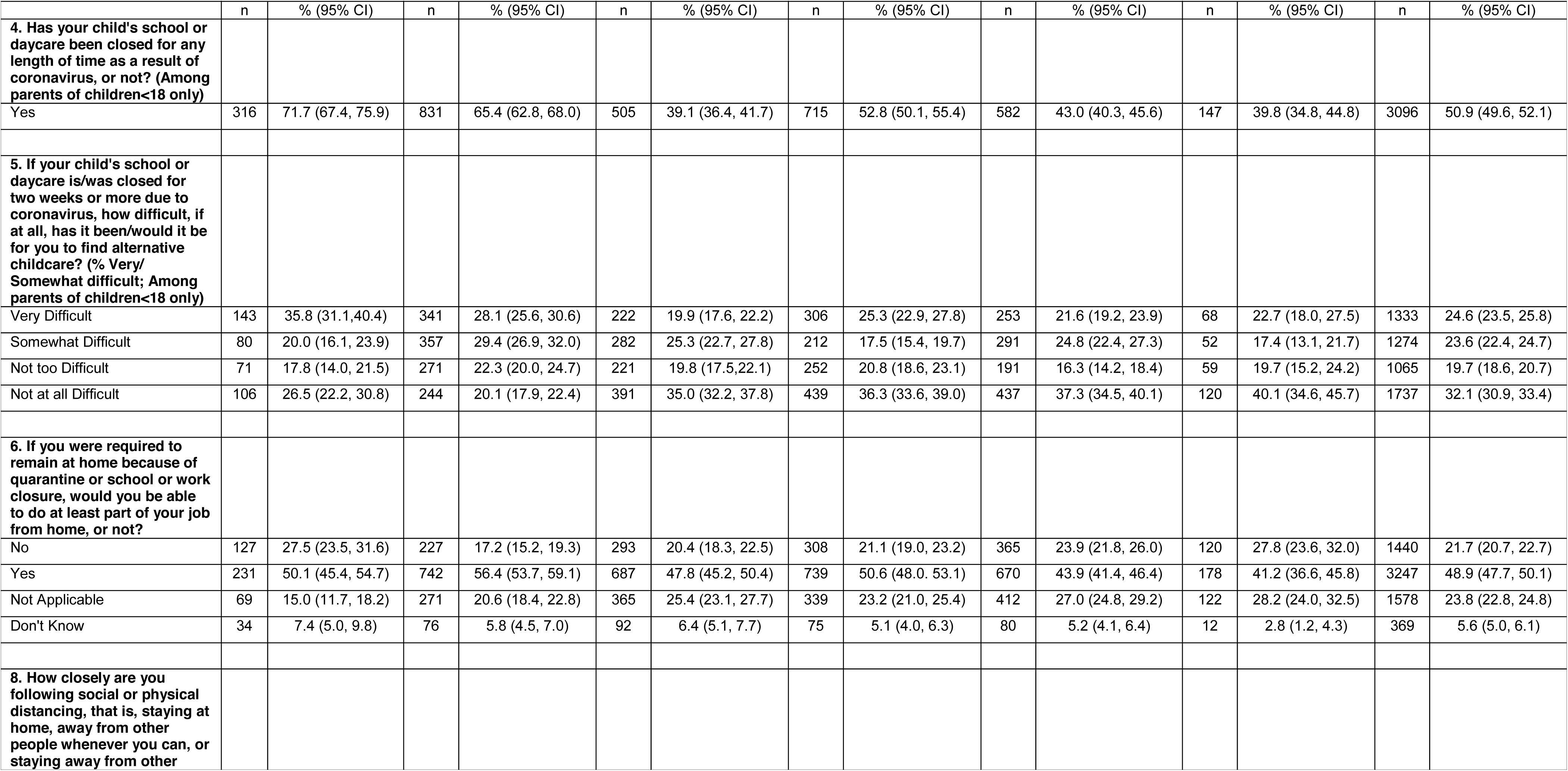
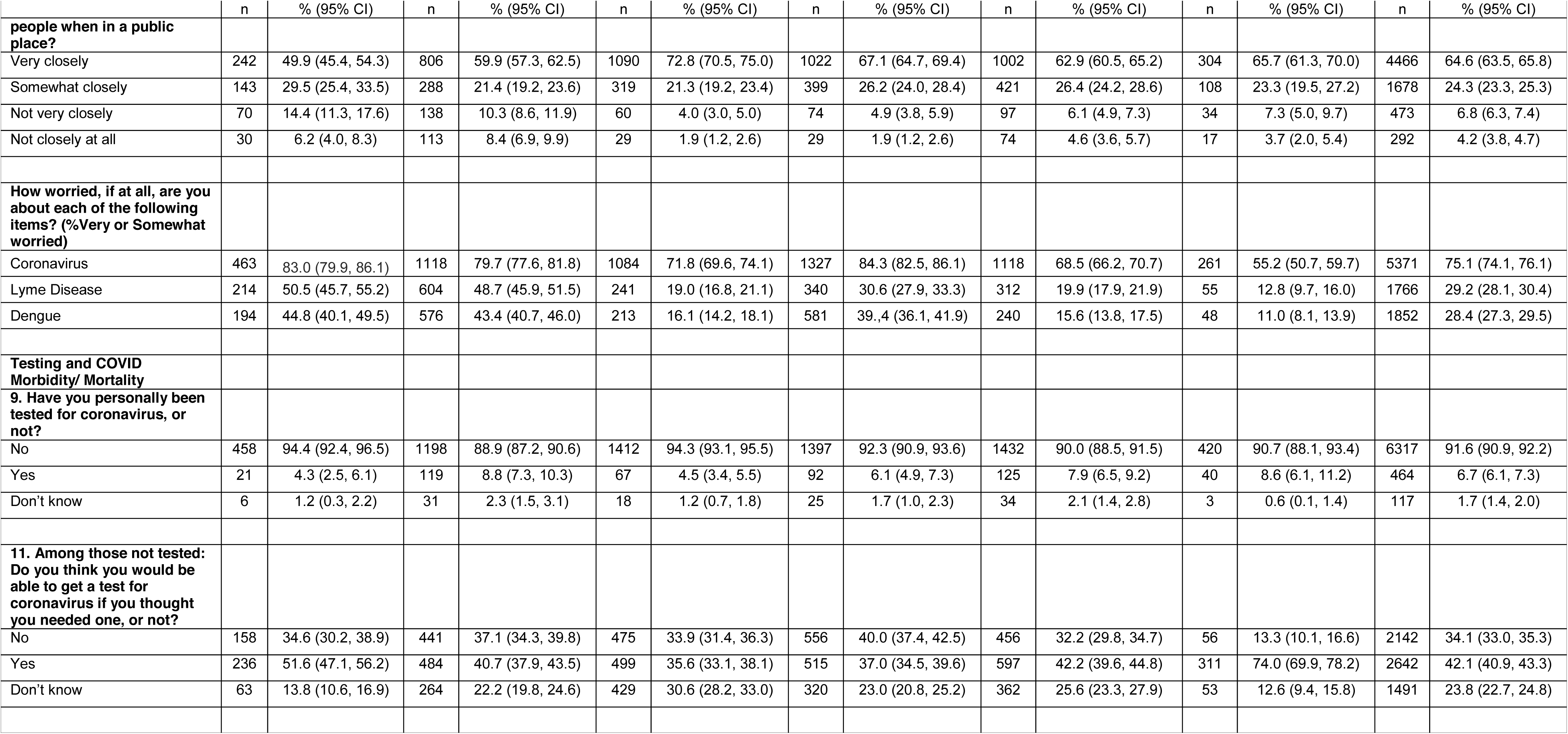
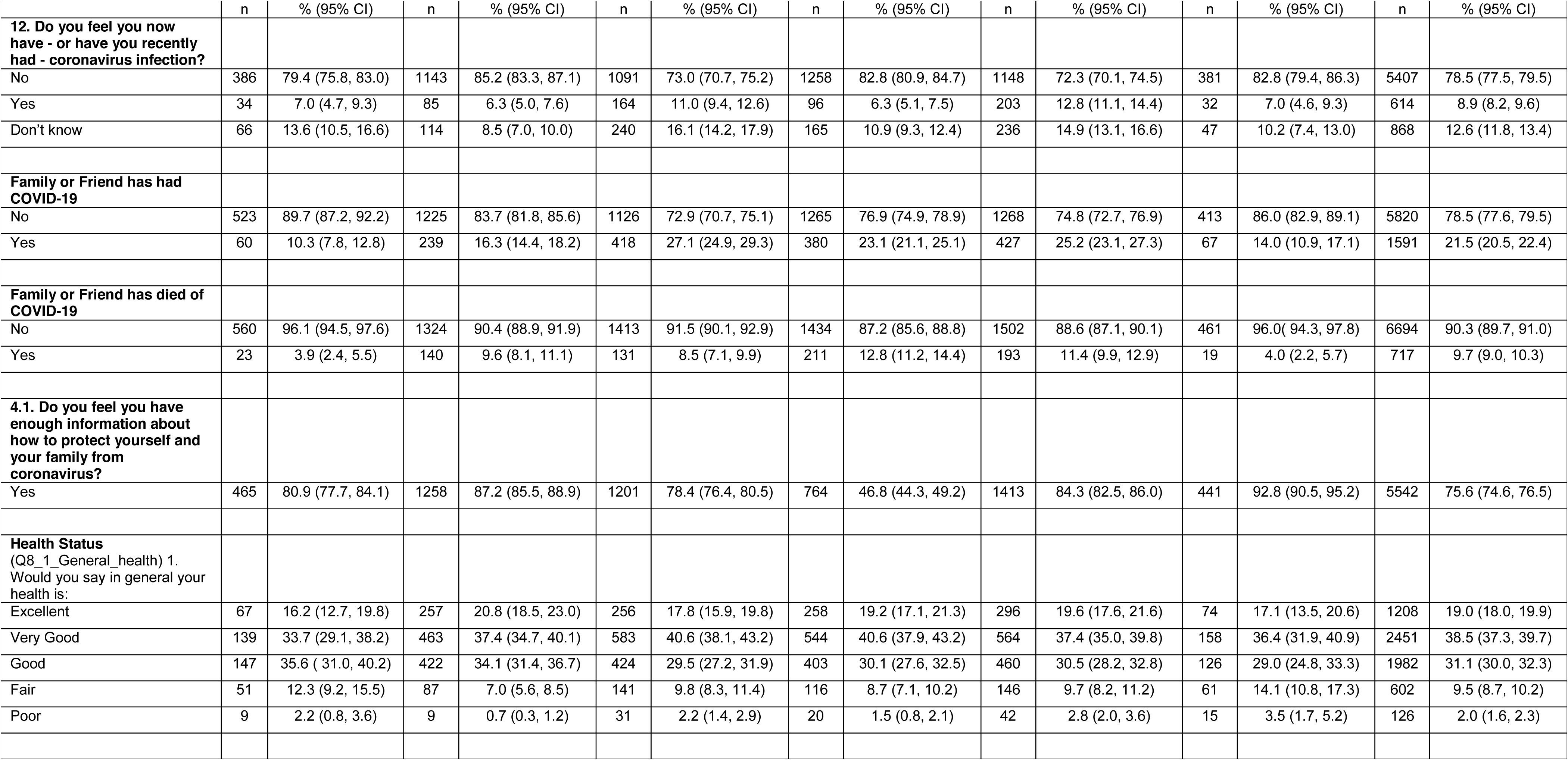
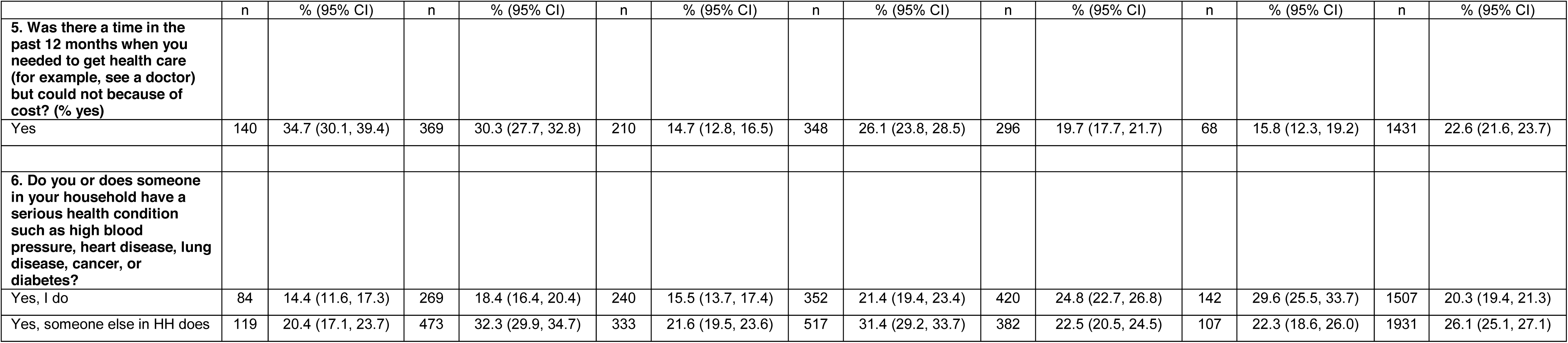
Global COVID Study participant COVID-19-related experiences, by UN region code.

Overall, 75% of respondents indicated they were worried about getting sick from COVID-19, with significantly higher proportions in Latin America and the Caribbean (84.3%), Africa (83.0%), and Asia (79.7%) than in Europe (71.8%), Northern America (68.5%), or Oceania (55.2%). Comparatively, respondents across regions were more than twice as likely to worry about getting sick from COVID-19 than from other significant infectious diseases (Lyme Disease and dengue).

Additionally, 64.6% of respondents indicated they were following social-physical distancing guidelines very closely, though respondents in Africa and in Asia reported they were most likely not to follow social distancing guidelines closely or at all (20.6% and 18.7%, respectively). Testing for COVID-19 was uncommon everywhere, with 6.7% of all respondents indicating they had been tested. Among those not tested, respondents in Oceania (74.0%) and in Africa (51.6%) were more likely to believe than were respondents from other regions that they would be able to obtain a test if needed. In total, 8.9% of the global sample believed they had coronavirus infection, with the highest proportion in Northern America (12.8%) and in Europe (11.0%). Respondents from Europe (27.1%), Northern America (25.2%), and Latin America (23.1%) were most likely to report they knew a family member, friend, or neighbor who had COVID-19, while respondents from Latin America (12.8%) and Northern America (11.4%) were more likely to report knowing someone who died from COVID-19. Finally, with the exception of Latin America and the Caribbean, more than most respondents indicated they had enough information to prevent coronavirus infection in their families. In Latin America, however, less than half (46.8%) felt they had sufficient information to prevent coronavirus.

#### Health and Health Care

Overall, 58.4% of respondents indicated that their health was “Excellent” or “Very Good” with little variation by region. Respondents from Africa (34.7%), Asia (30.3%), and Latin America (26.1%) were more likely to report they had difficulties obtaining health care when needed because of the cost. Respondents from Northern America (24.8%) and Oceania (29.6%) were more likely to indicate they had a chronic health condition, with respondents from Asia (32.3%) and Latin America (31.4%) most likely to indicate that someone in their household had a chronic health condition.

#### Psychosocial Scales

From Table 3, respondents from Africa reported the highest Perceived Stress Scale (PSS-Stress) levels – significantly higher than Latin America and the Caribbean, Northern America, and Oceania – followed by respondents from Asia and from Europe. In all regions, respondents reported a stronger “internal” health locus of control (meaning, respondents perceived they were in control their own health) than either “chance” or “powerful others” as health locus of control. Respondents from Africa, Asia, and Latin America reported higher levels of “internal” health locus of control than their counterparts from Europe, Northern America, and Oceania, while respondents from Asia and Northern America had the highest levels of “chance” health locus of control (i.e., one’s health is up to fate or luck). Respondents from Asia, Africa, and Latin America had higher levels of “powerful others” in control of their health (i.e., one’s health is influenced by external entities) when compared with respondents from Europe, Northern America, and Oceania. No differences in overall Perceived Social Support (PSS-Support) were noted among respondents from Asia, Europe, Latin America, Northern America, and Oceania, though respondents from Africa reported significantly lower overall levels of social support. Similar geographic patterns were observed, generally, for the subcomponents of PSS-Support (Family, Friends, Significant Others subscales).

**Table 3:** Global COVID Study psychosocial and COVID-related composite scores, by UN region code.

|  |  |  |  |  |  |  |  |  |  |  |  |  |  |  |
| --- | --- | --- | --- | --- | --- | --- | --- | --- | --- | --- | --- | --- | --- | --- |
|  | Africa |  | Asia |  | Europe |  | LAC |  | Northern America |  | Oceania |  | Total |  |
|  | n | Mean<br>(95% CI) | n | Mean<br>(95% CI) | n | Mean<br>(95% CI) | n | Mean<br>(95% CI) | n | Mean<br>(95% CI) | n | Mean<br>(95% CI) | n | Mean<br>(95% CI) |
| Perceived Stress Scale<br>(higher score = more stress) | 412 | 19.2<br>(18.5, 19.9) | 1190 | 18.6<br>(18.3, 18.9) | 1376 | 18.5<br>(18.1, 18.9) | 1363 | 18.1<br>(17.7, 18.4) | 1456 | 17.7<br>(17.3, 18.0) | 426 | 15.5<br>(14.8,16.2) | 6223 | 18.1<br>(17.9, 18.3) |
| Multidimensional Health<br>Locus of Control (Internal) | 547 | 22.2<br>(21.9, 22.5) | 1398 | 22.8<br>(22.6, 23.0) | 1488 | 20.4<br>(20.2, 20.6) | 1565 | 22.2<br>( 22.0, 22.4) | 1613 | 21.4<br>(21.3, 21.6) | 461 | 21.1<br>(20.8, 21.5) | 7072 | 21.7<br>(21.6, 21.8) |
| Multidimensional Health<br>Locus of Control (Chance) | 554 | 16.0<br>(15.6, 16.3) | 1402 | 17.5<br>(17.3, 17.8) | 1501 | 15.5<br>(15.3, 15.7) | 1553 | 13.9<br>(13.7, 14.1) | 1617 | 16.7<br>(16.5, 16.9) | 464 | 15.6<br>(15.3, 16.0) | 7091 | 15.9<br>(15.8, 16.0) |
| Multidimensional Health<br>Locus of Control (Powerful<br>Others) | 554 | 19.0<br>(18.6, 19.4) | 1404 | 20.6<br>(20.4, 20.9) | 1503 | 16.6<br>(16.4, 16.8) | 1574 | 18.9<br>(18.7, 19.1) | 1622 | 16.9<br>(16.7, 17.1) | 466 | 15.4<br>(15.0, 15.8) | 7123 | 18.1<br>(18.0, 18.2) |
| Mutlidimensional Score of<br>Perceived Social Support<br>(Significant Others) | 413 | 19.8<br>(19.2, 20.5) | 1211 | 20.6<br>(20.3, 21.0) | 1415 | 21.2<br>(20.9, 21.5) | 1340 | 21.2<br>(20.8, 21.5) | 1477 | 21.3<br>(21.0, 21.6) | 434 | 20.8<br>(20.2, 21.4) | 6290 | 21.0<br>(20.8, 21.1) |
| Mutlidimensional Score of<br>Perceived Social Support<br>(Family) | 414 | 19.6<br>(19.0, 20.3) | 1227 | 21.4<br>(21.1, 21.7) | 1403 | 20.5<br>(20.2, 20.8) | 1356 | 20.6<br>(20.3, 21.0) | 1480 | 20.7<br>(20.4, 21.0) | 432 | 20.3<br>(19.7, 20.8) | 6312 | 20.7<br>(20.5, 20.8) |
| Mutlidimensional Score of<br>Perceived Social Support<br>(Friend) | 416 | 18.0<br>(17.4, 18.5) | 1229 | 19.9<br>(19.6, 20.2) | 1417 | 19.8<br>(19.5, 20.1) | 1345 | 19.6<br>(19.3, 19.9) | 1482 | 20.2<br>(19.9, 20.5) | 428 | 19.9<br>(19.3, 20.4) | 6317 | 19.8<br>(19.6, 19.9) |
| Mutlidimensional Score of<br>Perceived Social Support<br>(Total) | 402 | 57.4<br>(55.8, 59.0) | 1178 | 61.9<br>(61.1, 62.8) | 1361 | 61.6<br>(60.8, 62.4) | 1304 | 61.3<br>(60.3, 62.2) | 1424 | 62.2<br>(61.4, 63.0) | 419 | 61.0<br>(59.5, 62.5) | 6088 | 61.4<br>(61.0, 61.8) |
| Standardized COVID-19<br>Knowledge Score (T-score;<br>higher score = more COVID-19 knowledge items<br>answered correctly) | 570 | 50.0<br>(49.3, 50.6) | 1446 | 45.6<br>(45.0, 46.1) | 1534 | 53.8<br>(53.4, 54.2) | 1629 | 50.3<br>(49.9,50.7) | 1668 | 49.8<br>(49.2, 50.4) | 478 | 50.9<br>(50.0,51.8) | 7325 | 50.0<br>(49.8, 50.2) |
| Standardized COVID-19<br>Attitudes Score (T-score;<br>higher score = more COVID-19-related worry) | 570 | 53.7<br>(52.9, 54.5) | 1432 | 53.3<br>(52.8, 53.8) | 1531 | 47.3<br>(46.8, 47.8) | 1633 | 52.6<br>(52.1, 53.0) | 1661 | 47.5<br>(47.0, 47.9) | 479 | 44.2<br>(43.3, 45.1) | 7306 | 50.0<br>(49.8, 50.2) |

Respondents from Europe reported the highest levels of the COVID-19 Knowledge Score while respondents from Asia reported the lowest; no differences in COVID-19-related knowledge were detected among respondents from Africa, Latin America, Northern America, and Oceania. Respondents from Africa, Asia, and Latin America reported higher levels of COVID-19-related worry (COVID-19 Attitudes Score) than did respondents in Europe, Northern America, and Oceania, with respondents from Oceania significantly least like to worry about COVID-19.

#### Non-medical COVID-19 related Impact

Table 4 presents items of non-medical personal impact relating to COVID-19 and the standardized non-medical COVID-19-related Impact Score with four sub-scores derived from those items. Respondents from Asia reported the highest overall non-medical personal impact, followed by respondents from Latin America and Africa (respectively). Respondents from Oceania reported the lowest personal impact. While little significant difference was observed in the relative strength of impact subcomponents within regions, respondents in Asia and Latin America had significantly higher “Livelihood” personal impact scores and “Personal Action” impact scores than did respondents in other regions.

**Table 4:** Global COVID Study non-medical COVID-19-related impact scores and items, by UN region code.

|  |  |  |  |  |  |  |  |  |  |  |  |  |  |  |
| --- | --- | --- | --- | --- | --- | --- | --- | --- | --- | --- | --- | --- | --- | --- |
|  | Africa |  | Asia |  | Europe |  | LAC |  | Northern America |  | Oceania |  | Total |  |
|  | n | % (95% CI) | n | % (95% CI) | n | % (95% CI) | n | % (95% CI) | n | % (95% CI) | n | % (95% CI) | n | % (95% CI) |
| **4.6. Please tell us if you have taken any of the following actions because of the recent coronavirus outbreak.** |  |  |  |  |  |  |  |  |  |  |  |  |  |  |
| a. Decided not to travel or changed travel plans (CANCEL) | 455 | 78.0(74.7,81.4) | 1262 | 86.2(84.4, 88.0) | 1162 | 75.3(73.1, 77.4) | 1228 | 74.7(72.5, 76.8) | 1177 | 69.4(67.2,71.6) | 364 | 75.8(72.0, 79.7) | 5648 | 76.2(75.2,77.2) |
| b. Bought or worn a protective mask (PERSONAL ACTION) | 442 | 75.8(72.3, 72.3) | 1270 | 86.7(85.0,88.5) | 1026 | 66.5(64.1,68.8) | 1433 | 87.1(85.5,88.7) | 1221 | 72.0(69.9,74.2) | 233 | 48.5(44.1, 53.0) | 5625 | 75.9(74.9,76.9) |
| c. Stocked up on items such as food and household supplies (PERSONAL ACTION) | 372 | 63.8(59.9, 67.7) | 1026 | 70.1(67.7,72.4) | 827 | 53.6(51.1, 56.0) | 1189 | 72.3(70.1,74.4) | 1138 | 67.1(64.9, 69.4) | 243 | 50.6(46.2,55.1) | 4795 | 64.7(63.6, 65.8) |
| d. Postponed or canceled health care visits (PERSONAL ACTION) | 226 | 38.8(34.8,42.7) | 909 | 62.1(59.6,64.6) | 738 | 47.8(45.3, 50.3) | 1048 | 63.7(61.4, 66.0) | 946 | 55.8(53.4,58.2) | 175 | 36.5(32.2, 40.8) | 4042 | 54.5(53.4, 55.7) |
| e. Got extra refills on prescription medication (PERSONAL ACTION) | 159 | 27.3(23.7,30.9) | 764 | 52.2(49.6, 54.7) | 327 | 21.2(19.1, 23.2) | 575 | 35.0(32.7,37.3) | 514 | 30.3(28.1, 32.5) | 144 | 30.0(25.9, 34.1) | 2483 | 33.5(32.4, 34.6) |
| f. Stayed home instead of going to work school, or other regular activities (PERSONAL ACTION) | 410 | 70.3(66.6, 74.0) | 1228 | 83.9(82.0, 85.8) | 1284 | 83.2(81.3, 85.0) | 1341 | 81.5(79.6, 83.4) | 1278 | 75.4(73.3, 77.4) | 376 | 78.3(74.6, 82.0) | 5917 | 79.8(78.9, 80.8) |
| g. Postponed or canceled a medical procedure or surgery (CANCEL) | 125 | 21.4(18.4, 24.8) | 601 | 41.1(38.5, 43.6) | 280 | 18.1(16.2, 20.1) | 467 | 28.4(26.2, 30.6) | 467 | 27.6(25.4, 29.7) | 95 | 19.8(16.2, 23.4) | 2035 | 27.5(26.4, 28.5) |
|  | Africa |  | Asia |  | Europe |  | LAC |  | Northern America |  | Oceania |  | Total |  |
|  | n | % (95% CI) | n | % (95% CI) | n | % (95% CI) | n | % (95% CI) | n | % (95% CI) | n | % (95% CI) | n | % (95% CI) |
| h. Canceled plans to attend large gatherings such as concerts or sporting events (CANCEL) | 412 | 70.7(67.0, 74.4) | 1138 | 77.7(75.6, 79.9) | 1008 | 65.3(62.9,67.7) | 1247 | 75.8(73.7, 77.9) | 1044 | 61.6(59.3, 63.9) | 277 | 57.7(53.3, 62.1) | 5126 | 69.2(68.1, 70.2) |
| i. Quit my job (LIVELIHOOD) | 59 | 10.1(7.7, 12.6) | 254 | 17.3(15.4, 19.3) | 122 | 7.9(6.6, 9.2) | 262 | 15.9(14.2, 17.7) | 163 | 9.6(8.2, 11.0) | 31 | 6.5(4.3, 8.7) | 891 | 12.0(11.3, 12.8) |
| **7. Have you experienced any of the following because of coronavirus?** |  |  |  |  |  |  |  |  |  |  |  |  |  |  |
| 7a. Lost income from a job or business (LIVELIHOOD) | 240 | 41.2(37.2,45.2) | 631 | 43.1, 40.6, 45.6) | 515 | 33.4(31.0, 35.7) | 795 | 48.3(45.9, 50.7) | 557 | 32.9 (30.6, 35.1) | 198 | 41.3(36.8, 45.7) | 2936 | 39.6( 38.5, 40.7) |
| 7b. Been unable to get groceries (SUPPLIES) | 160 | 27.4(23.8, 31.1) | 387 | 26.4(24.2, 28.7) | 262 | 17.0(15.1, 18.8) | 309 | 18.8(16.9, 20.7) | 395 | 23.3(21.3, 25.3) | 148 | 30.8(26.7, 35.0) | 1661 | 22.4(21.5, 23.4) |
| 7c. Been unable to get cleaning supplies or hand sanitizer (SUPPLIES) | 144 | 24.7(21.2, 28.2) | 447 | 30.5(28.2, 32.9) | 464 | 30.1(27.8, 32.3) | 438 | 26.6(24.5,28.8) | 748 | 44.1(41.8, 46.5) | 211 | 44.0(39.5, 48.4) | 2452 | 33.1(32.0,34.2) |
| 7d. Been unable to get prescription medication (SUPPLIES) | 62 | 10.6 (8.1, 13.2) | 207 | 14.1(12.4, 15.9) | 65 | 4.2(3.2, 5.2) | 240 | 14.6(12.9, 16.3) | 120 | 7.1(5.9, 8.3) | 44 | 9.2(6.6, 11.7) | 738 | 10.0(9.3, 10.6) |
| 7e. Have you or a family member been harassed, bullied, or hurt because | 59 | 10.1 (7.7, 12.6) | 141 | 9.6 (8.1, 11.1) | 76 | 4.9 (3.8, 6.0) | 99 | 6.0 (4.9, 7.2) | 172 | 10.1 (8.7, 11.6) | 48 | 10.0 (7.3, 12.7) | 595 | 8.0 (7.4, 8.6) |
|  | Africa |  | Asia |  | Europe |  | LAC |  | Northern America |  | Oceania |  | Total |  |
|  | n | % (95% CI) | n | % (95% CI) | n | % (95% CI) | n | % (95% CI) | n | % (95% CI) | n | % (95% CI) | n | % (95% CI) |
| of coronavirus<br>(LIVELIHOOD) |  |  |  |  |  |  |  |  |  |  |  |  |  |  |
| **Standardized non-medical COVID-19 Personal Impact Score (T-score; higher score =more impact items noted)** | 529 | 50.5<br>(49.7, 51.3) | 1397 | 54.5<br>(53.4, 55.0) | 1516 | 46.0<br>(45.6, 46.5) | 1576 | 51.9<br>(51.4, 52.3) | 1635 | 48.9<br>(48.4, 49.4) | 473 | 46.4<br>(45.5, 47.4) | 7126 | 50.0<br>(49.8,50.2) |
| **Personal Actions Impact Subscore** | 529 | 49.0<br>(48.2, 49.8) | 1400 | 54.0<br>(53.5, 54.4) | 1516 | 46.7<br>(46.2, 47.1) | 1578 | 52.5<br>(52.1, 53.0) | 1638 | 49.2<br>(48.7, 49.7) | 474 | 44.3<br>(43.4, 45.3) | 7135 | 50.0<br>(49.8, 50.2) |
| **Cancelled Impact Subscore** | 521 | 51.0<br>(50.2, 51.8) | 1381 | 54.3<br>(53.8, 54.7) | 1508 | 47.8<br>(47.3, 48.3) | 1544 | 50.8<br>(50.4, 51.3) | 1618 | 48.1<br>(47.6, 48.6) | 470 | 47.2<br>(46.3, 48.1) | 7042 | 50.0<br>(49.8, 50.2) |
| **Supplies Impact Subscore** | 505 | 50.4<br>(49.5, 51.4) | 1362 | 50.8<br>(50.2, 51.4) | 1502 | 48.1<br>(47.7, 48.5) | 1535 | 49.4<br>(48.9, 49.9) | 1599 | 51.0<br>(50.5, 51.5) | 463 | 51.9<br>(50.9, 52.8) | 6966 | 50.0<br>(49.8, 50.2) |
| **Livelihood Impact Subscore** | 498 | 51.2<br>(50.2, 52.1) | 1354 | 51.7<br>(51.1, 52.3) | 1489 | 47.9<br>(47.4, 48.3) | 1537 | 51.6<br>(51.1, 52.0) | 1600 | 48.9<br>(48.4, 49.3) | 468 | 49.5<br>(48.6, 50.4) | 6946 | 50.0<br>(49.8, 50.2) |

The most commonly noted impact item (Table 4) was that the respondent “stayed home instead of going to work or school,” with 79% of the sample indicating positive agreement. The proportion of respondents staying home was highest in Asia (83.9%) and lowest in Africa (70.3%). The next most common impact item noted is “bought or worn a protective mask” (75.9%), with the highest proportion of mask use found in Latin America and the Caribbean (87.1%) and in Asia (86.7%). The lowest proportion of mask use was in Oceania (48.5%). The third most common impact item was that the respondent decided to cancel travel plans (76.2%), most commonly found in Asia where 86.2% of respondents cancelled travel plans.

### Univariate Qualitative Findings

In total, 6859 respondents (92.6% of the final sample size) generated 20,015 qualitative excerpts for coding. Table 5 presents the top 10 codes most commonly applied to the open-ended responses, with exemplary quotations selected by region. Statements reflecting worsening mental health (62.2%) and stress (54.0%) were most frequently mentioned by respondents, respectively, in all regions. Non-medical impacts (namely, changes in work (38.5%), social-physical distancing and lockdown (30.3%), and struggle to access basic goods and services (27.6%)) were the next codes most commonly applied. Some respondents (22.6%) were concerned about their limitations on freedoms brought on by shutdowns, prevention strategies, and policies. Issues obtaining cleaning supplies (17.4%) and shortages (15.2%) were also commonly noted, although typically less frequently than livelihood and personal impacts. Unemployment-related issues were noted by 15.1% of respondents. Almost one in four respondents (25.5%) indicated they were essentially unaffected by COVID-19 or were coping with COVID-19, the 5^th^ most common code used in Asia, Europe, and Latin America, 6^th^ in Africa, and 7^th^ Northern America and Oceania. Generally, we observed little regional variation in the ranking of the top 10 commonly-applied codes, with the exception that “supply shortages” were more commonly cited (9^th^ rank in each) in Europe, Northern America, and Oceania, and with “worry about finances” ranking 9^th^ in Africa and 10^th^ in Asia.

**Table 5:**
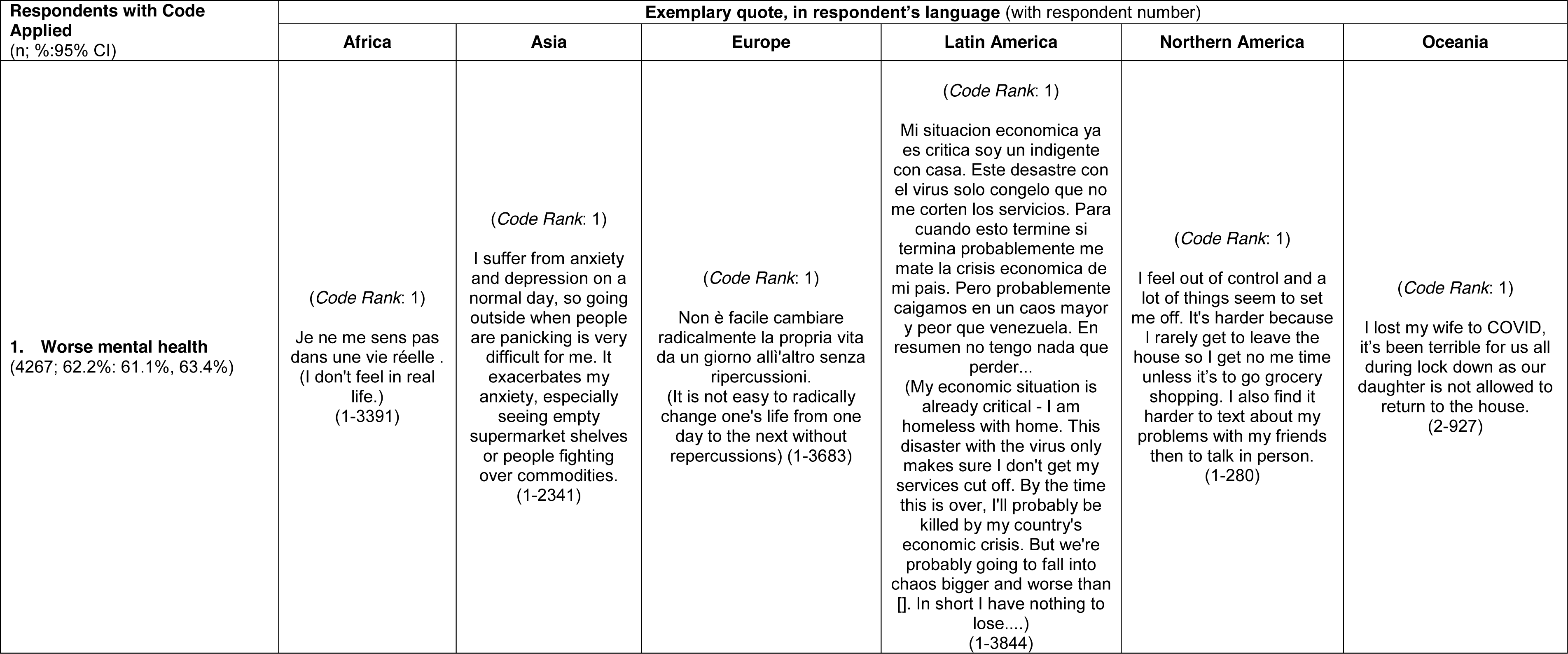
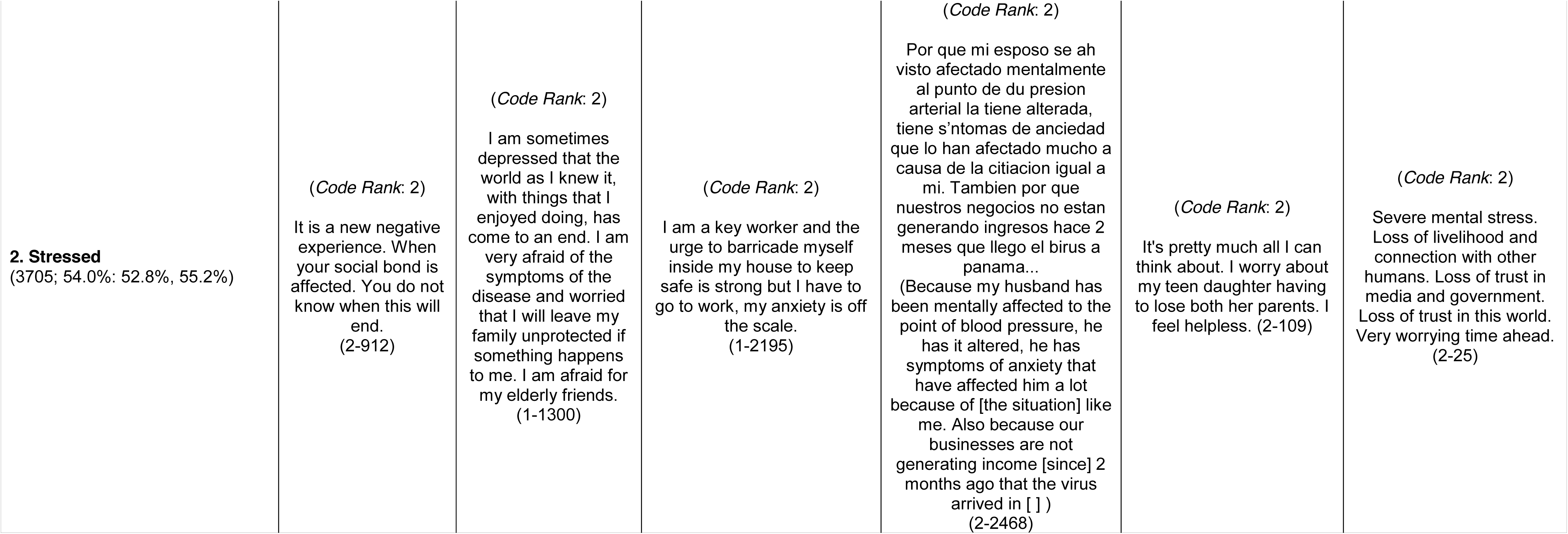
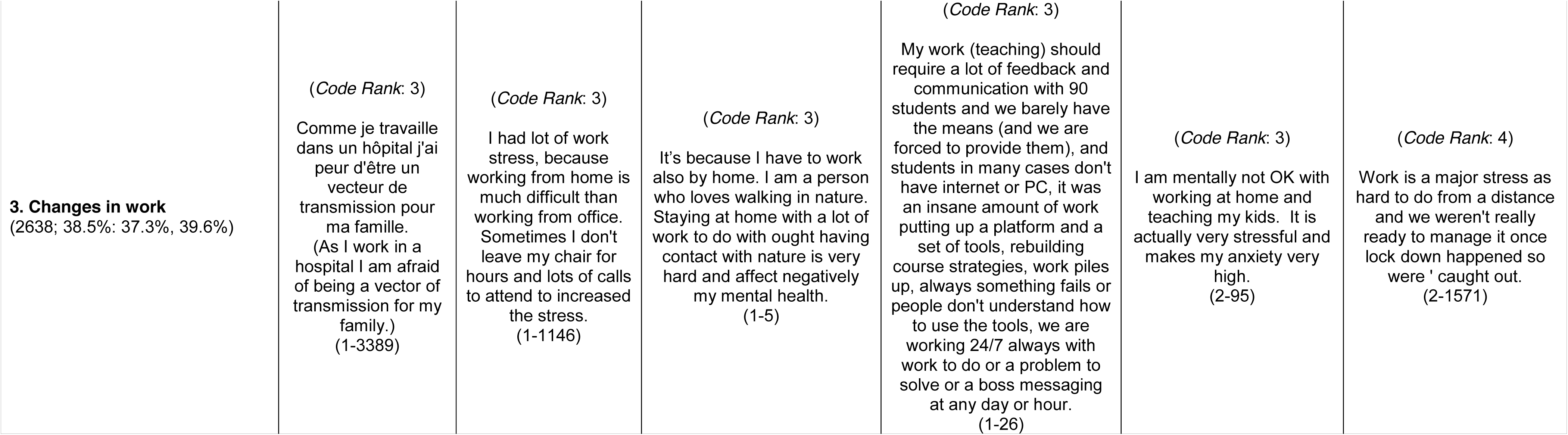
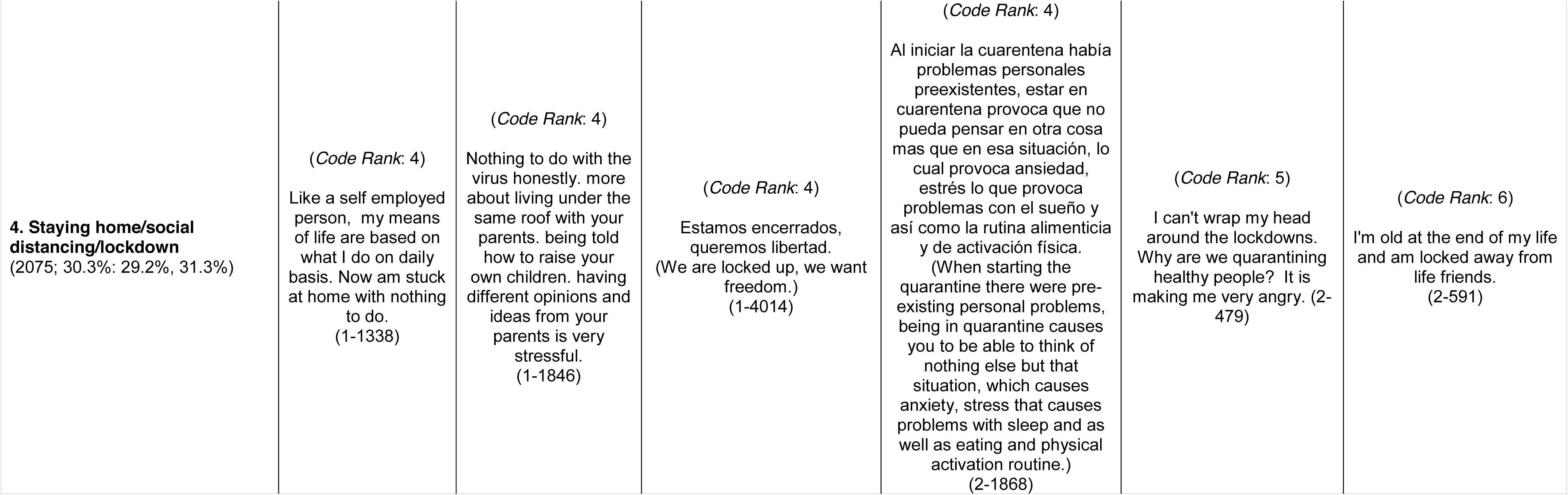
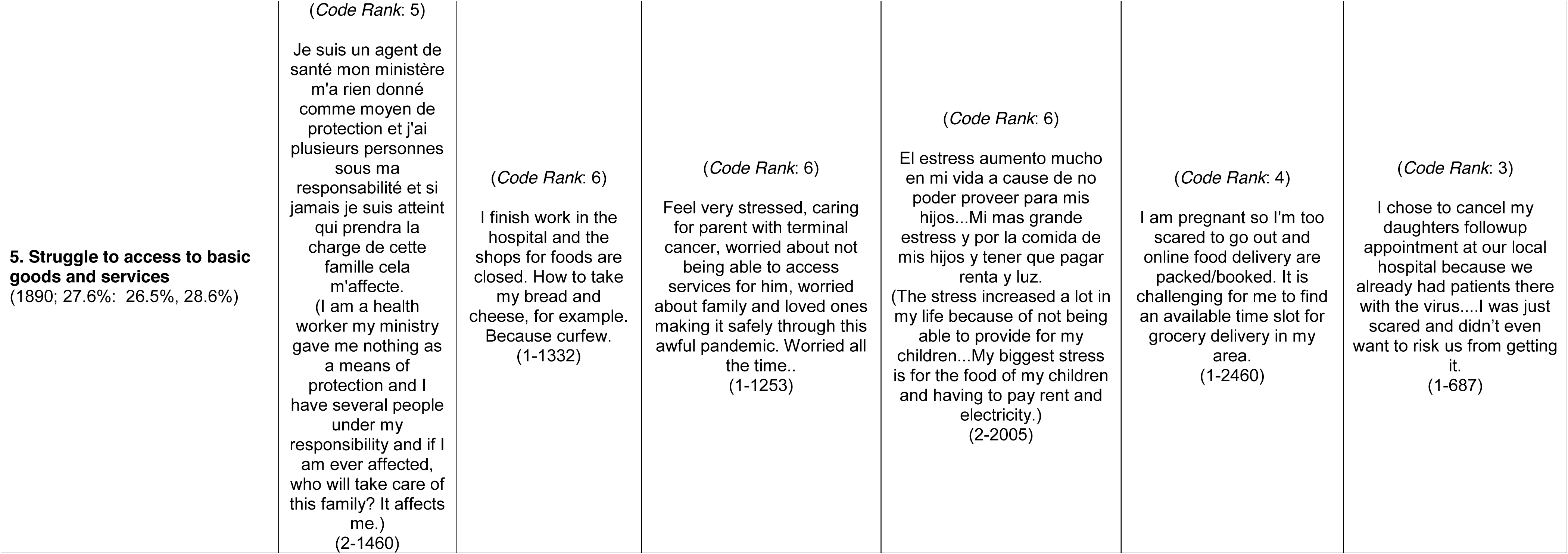
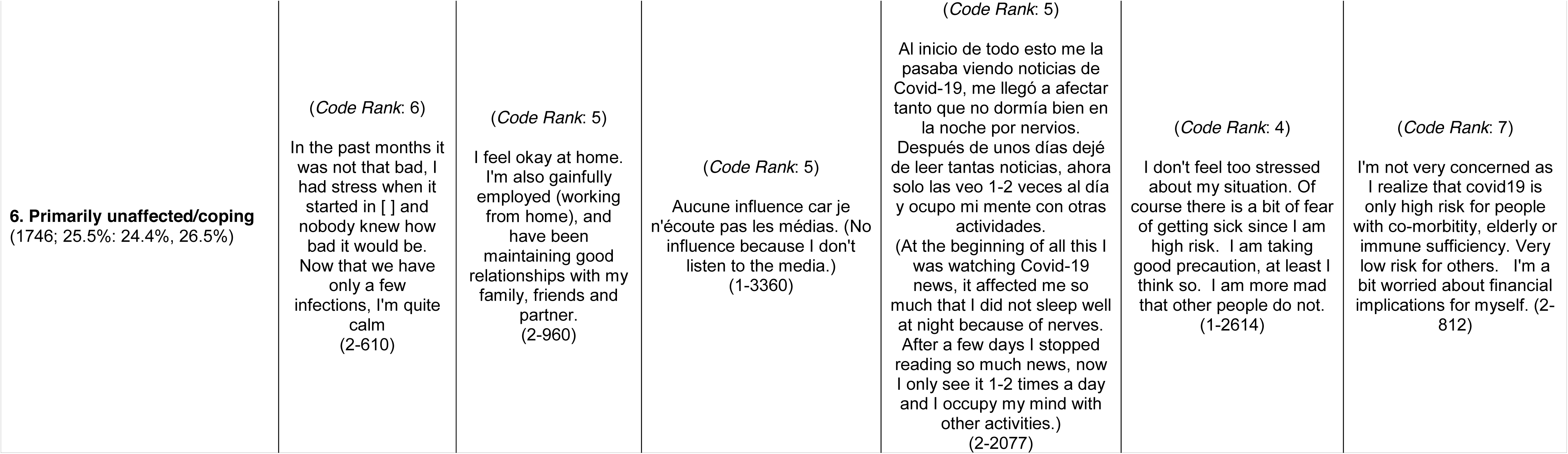
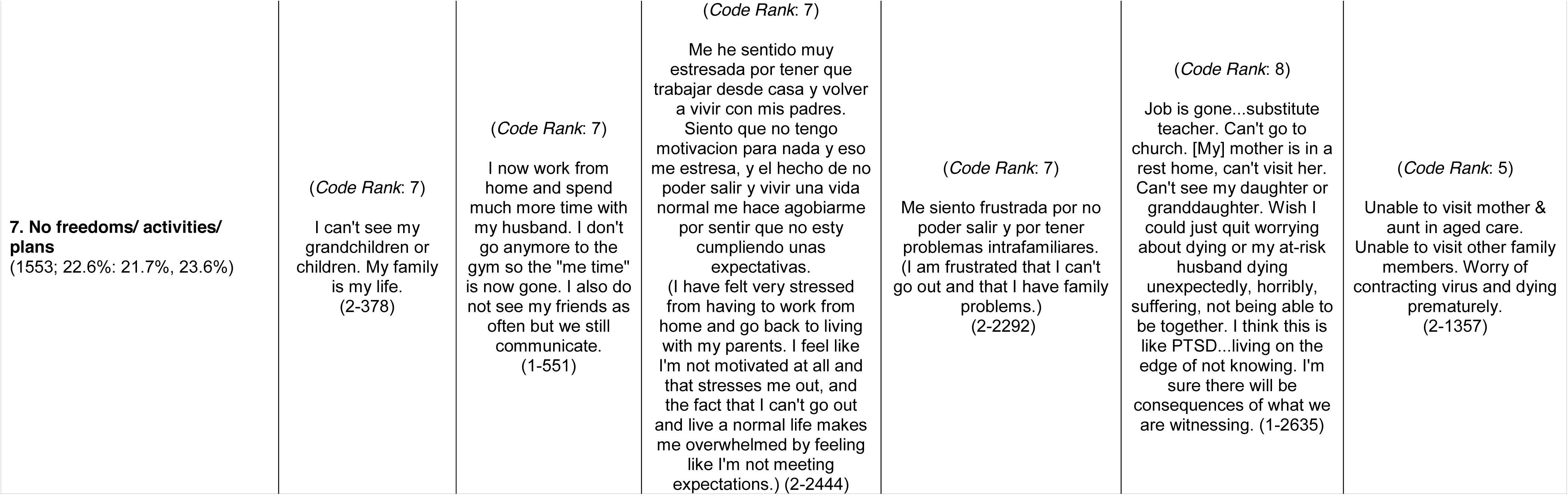
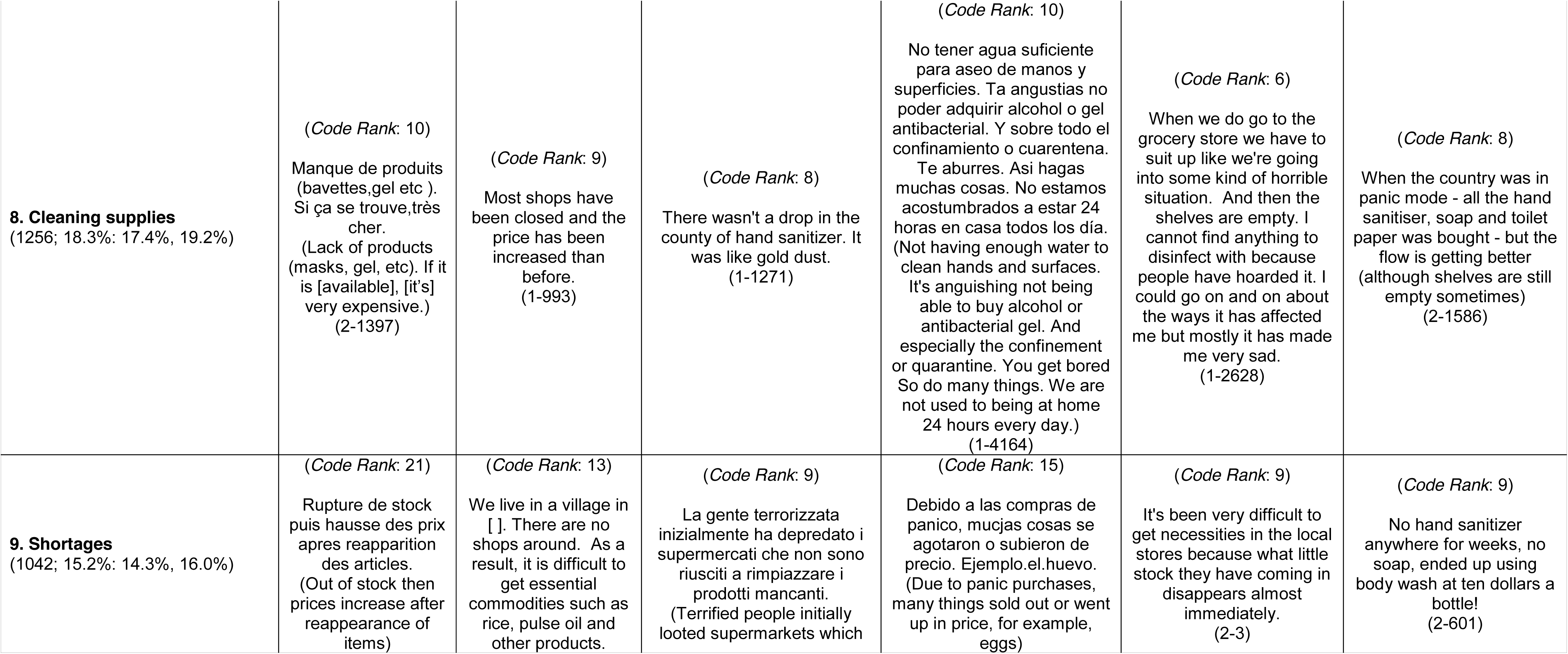
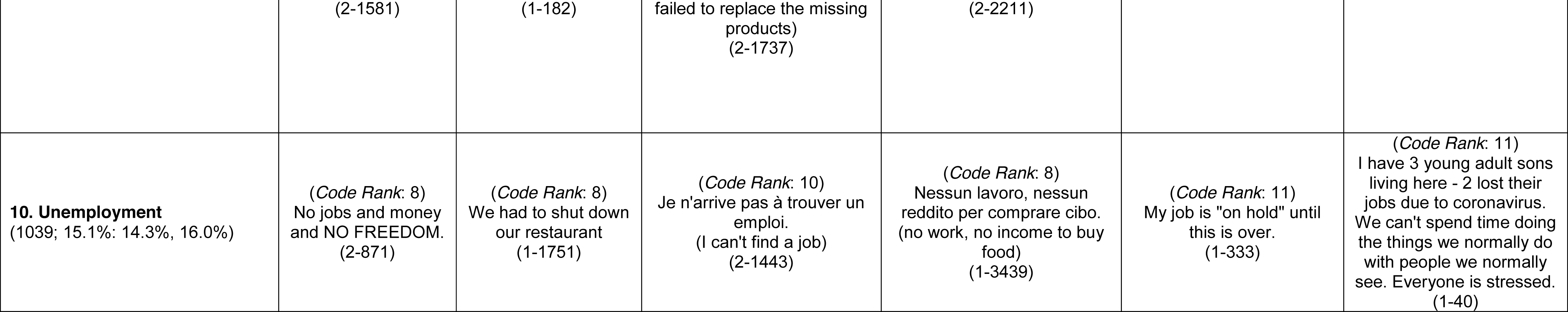
10 Most Commonly-applied Open-ended Codes in descending order with exemplary quotes, by World Region.

| Respondents with Code Applied<br>(n; %:95% CI) | Exemplary quote, in respondent's language (with respondent number) |  |  |  |  |  |
| --- | --- | --- | --- | --- | --- | --- |
|  | Africa | Asia | Europe | Latin America | Northern America | Oceania |
| **1. Worse mental health**<br>(4267; 62.2%: 61.1%, 63.4%) | (Code Rank: 1)<br>Je ne me sens pas dans une vie réelle .<br>(I don't feel in real life.)<br>(1-3391) | (Code Rank: 1)<br>I suffer from anxiety and depression on a normal day, so going outside when people are panicking is very difficult for me. It exacerbates my anxiety, especially seeing empty supermarket shelves or people fighting over commodities.<br>(1-2341) | (Code Rank: 1)<br>Non è facile cambiare radicalmente la propria vita da un giorno all'altro senza ripercussioni.<br>(It is not easy to radically change one's life from one day to the next without repercussions) (1-3683) | (Code Rank: 1)<br>Mi situacion economica ya es critica soy un indigente con casa. Este desastre con el virus solo congelo que no me corten los servicios. Para cuando esto termine si termina probablemente me mate la crisis economica de mi pais. Pero probablemente caigamos en un caos mayor y peor que venezuela. En resumen no tengo nada que perder...<br>(My economic situation is already critical - I am homeless with home. This disaster with the virus only makes sure I don't get my services cut off. By the time this is over, I'll probably be killed by my country's economic crisis. But we're probably going to fall into chaos bigger and worse than []. In short I have nothing to lose....)<br>(1-3844) | (Code Rank: 1)<br>I feel out of control and a lot of things seem to set me off. It's harder because I rarely get to leave the house so I get no me time unless it's to go grocery shopping. I also find it harder to text about my problems with my friends then to talk in person.<br>(1-280) | (Code Rank: 1)<br>I lost my wife to COVID, it's been terrible for us all during lock down as our daughter is not allowed to return to the house.<br>(2-927) |

|  |  |  |  |  |  |  |
| --- | --- | --- | --- | --- | --- | --- |
| Respondents with Code Applied<br>(n; %:95% CI) | Exemplary quote, in respondent's language (with respondent number) |  |  |  |  |  |
|  | Africa | Asia | Europe | Latin America | Northern America | Oceania |
| **2. Stressed**<br>(3705; 54.0%: 52.8%, 55.2%) | (Code Rank: 2)<br>It is a new negative experience. When your social bond is affected. You do not know when this will end.<br>(2-912) | (Code Rank: 2)<br>I am sometimes depressed that the world as I knew it, with things that I enjoyed doing, has come to an end. I am very afraid of the symptoms of the disease and worried that I will leave my family unprotected if something happens to me. I am afraid for my elderly friends.<br>(1-1300) | (Code Rank: 2)<br>I am a key worker and the urge to barricade myself inside my house to keep safe is strong but I have to go to work, my anxiety is off the scale.<br>(1-2195) | (Code Rank: 2)<br>Por que mi esposo se ah visto afectado mentalmente al punto de du presion arterial la tiene alterada, tiene s'ntomas de ansiedad que lo han afectado mucho a causa de la citiacion igual a mi. Tambien por que nuestros negocios no estan generando ingresos hace 2 meses que lleo el birus a panama...<br>(Because my husband has been mentally affected to the point of blood pressure, he has it altered, he has symptoms of anxiety that have affected him a lot because of [the situation] like me. Also because our businesses are not generating income [since] 2 months ago that the virus arrived in [ ] )<br>(2-2468) | (Code Rank: 2)<br>It's pretty much all I can think about. I worry about my teen daughter having to lose both her parents. I feel helpless. (2-109) | (Code Rank: 2)<br>Severe mental stress. Loss of livelihood and connection with other humans. Loss of trust in media and government. Loss of trust in this world. Very worrying time ahead.<br>(2-25) |
| Respondents with Code Applied<br>(n; %:95% CI) | Exemplary quote, in respondent's language (with respondent number) |  |  |  |  |  |
|  | Africa | Asia | Europe | Latin America | Northern America | Oceania |
| **3. Changes in work**<br>(2638; 38.5%: 37.3%, 39.6%) | (Code Rank: 3)<br>Comme je travaille dans un hôpital j'ai peur d'être un vecteur de transmission pour ma famille. (As I work in a hospital I am afraid of being a vector of transmission for my family.) (1-3389) | (Code Rank: 3)<br>I had lot of work stress, because working from home is much difficult than working from office. Sometimes I don't leave my chair for hours and lots of calls to attend to increased the stress. (1-1146) | (Code Rank: 3)<br>It's because I have to work also by home. I am a person who loves walking in nature. Staying at home with a lot of work to do with ought having contact with nature is very hard and affect negatively my mental health. (1-5) | (Code Rank: 3)<br>My work (teaching) should require a lot of feedback and communication with 90 students and we barely have the means (and we are forced to provide them), and students in many cases don't have internet or PC, it was an insane amount of work putting up a platform and a set of tools, rebuilding course strategies, work piles up, always something fails or people don't understand how to use the tools, we are working 24/7 always with work to do or a problem to solve or a boss messaging at any day or hour. (1-26) | (Code Rank: 3)<br>I am mentally not OK with working at home and teaching my kids. It is actually very stressful and makes my anxiety very high. (2-95) | (Code Rank: 4)<br>Work is a major stress as hard to do from a distance and we weren't really ready to manage it once lock down happened so were ' caught out. (2-1571) |
|  | Africa | Asia | Europe | Latin America | Northern America | Oceania |
| **4. Staying home/social distancing/lockdown**<br>(2075; 30.3%: 29.2%, 31.3%) | (Code Rank: 4)<br>Like a self employed person, my means of life are based on what I do on daily basis. Now am stuck at home with nothing to do.<br>(1-1338) | (Code Rank: 4)<br>Nothing to do with the virus honestly. more about living under the same roof with your parents. being told how to raise your own children. having different opinions and ideas from your parents is very stressful.<br>(1-1846) | (Code Rank: 4)<br>Estamos encerrados, queremos libertad.<br>(We are locked up, we want freedom.)<br>(1-4014) | (Code Rank: 4)<br>Al iniciar la cuarentena había problemas personales preexistentes, estar en cuarentena provoca que no pueda pensar en otra cosa mas que en esa situación, lo cual provoca ansiedad, estrés lo que provoca problemas con el sueño y así como la rutina alimenticia y de activación física.<br>(When starting the quarantine there were pre-existing personal problems, being in quarantine causes you to be able to think of nothing else but that situation, which causes anxiety, stress that causes problems with sleep and as well as eating and physical activation routine.)<br>(2-1868) | (Code Rank: 5)<br>I can't wrap my head around the lockdowns. Why are we quarantining healthy people? It is making me very angry. (2-479) | (Code Rank: 6)<br>I'm old at the end of my life and am locked away from life friends.<br>(2-591) |

|  |  |  |  |  |  |  |
| --- | --- | --- | --- | --- | --- | --- |
| Respondents with Code Applied<br>(n; %:95% CI) | Exemplary quote, in respondent's language (with respondent number) |  |  |  |  |  |
|  | Africa | Asia | Europe | Latin America | Northern America | Oceania |
| **5. Struggle to access to basic goods and services**<br>(1890; 27.6%: 26.5%, 28.6%) | *(Code Rank: 5)*<br>Je suis un agent de santé mon ministère m'a rien donné comme moyen de protection et j'ai plusieurs personnes sous ma responsabilité et si jamais je suis atteint qui prendra la charge de cette famille cela m'affecte.<br>(I am a health worker my ministry gave me nothing as a means of protection and I have several people under my responsibility and if I am ever affected, who will take care of this family? It affects me.)<br>(2-1460) | *(Code Rank: 6)*<br>I finish work in the hospital and the shops for foods are closed. How to take my bread and cheese, for example.<br>Because curfew.<br>(1-1332) | *(Code Rank: 6)*<br>Feel very stressed, caring for parent with terminal cancer, worried about not being able to access services for him, worried about family and loved ones making it safely through this awful pandemic. Worried all the time..<br>(1-1253) | *(Code Rank: 6)*<br>El estress aumento mucho en mi vida a cause de no poder proveer para mis hijos...Mi mas grande estress y por la comida de mis hijos y tener que pagar renta y luz.<br>(The stress increased a lot in my life because of not being able to provide for my children...My biggest stress is for the food of my children and having to pay rent and electricity.)<br>(2-2005) | *(Code Rank: 4)*<br>I am pregnant so I'm too scared to go out and online food delivery are packed/booked. It is challenging for me to find an available time slot for grocery delivery in my area.<br>(1-2460) | *(Code Rank: 3)*<br>I chose to cancel my daughters followup appointment at our local hospital because we already had patients there with the virus....I was just scared and didn't even want to risk us from getting it.<br>(1-687) |
| Respondents with Code Applied<br>(n; %:95% CI) | Exemplary quote, in respondent's language (with respondent number) |  |  |  |  |  |
|  | Africa | Asia | Europe | Latin America | Northern America | Oceania |
| **6. Primarily unaffected/coping**<br>(1746; 25.5%: 24.4%, 26.5%) | (Code Rank: 6)<br>In the past months it was not that bad, I had stress when it started in [ ] and nobody knew how bad it would be. Now that we have only a few infections, I'm quite calm (2-610) | (Code Rank: 5)<br>I feel okay at home. I'm also gainfully employed (working from home), and have been maintaining good relationships with my family, friends and partner. (2-960) | (Code Rank: 5)<br>Aucune influence car je n'écoute pas les médias. (No influence because I don't listen to the media.) (1-3360) | (Code Rank: 5)<br>Al inicio de todo esto me la pasaba viendo noticias de Covid-19, me llegó a afectar tanto que no dormía bien en la noche por nervios. Después de unos días dejé de leer tantas noticias, ahora solo las veo 1-2 veces al día y ocupo mi mente con otras actividades. (At the beginning of all this I was watching Covid-19 news, it affected me so much that I did not sleep well at night because of nerves. After a few days I stopped reading so much news, now I only see it 1-2 times a day and I occupy my mind with other activities.) (2-2077) | (Code Rank: 4)<br>I don't feel too stressed about my situation. Of course there is a bit of fear of getting sick since I am high risk. I am taking good precaution, at least I think so. I am more mad that other people do not. (1-2614) | (Code Rank: 7)<br>I'm not very concerned as I realize that covid19 is only high risk for people with co-morbidity, elderly or immune sufficiency. Very low risk for others. I'm a bit worried about financial implications for myself. (2-812) |
|  | Africa | Asia | Europe | Latin America | Northern America | Oceania |
| **7. No freedoms/ activities/ plans**<br>(1553; 22.6%: 21.7%, 23.6%) | (Code Rank: 7)<br>I can't see my grandchildren or children. My family is my life.<br>(2-378) | (Code Rank: 7)<br>I now work from home and spend much more time with my husband. I don't go anymore to the gym so the "me time" is now gone. I also do not see my friends as often but we still communicate.<br>(1-551) | (Code Rank: 7)<br>Me he sentido muy estresada por tener que trabajar desde casa y volver a vivir con mis padres. Siento que no tengo motivacion para nada y eso me estresa, y el hecho de no poder salir y vivir una vida normal me hace agobiarme por sentir que no esty cumpliendo unas expectativas.<br>(I have felt very stressed from having to work from home and go back to living with my parents. I feel like I'm not motivated at all and that stresses me out, and the fact that I can't go out and live a normal life makes me overwhelmed by feeling like I'm not meeting expectations.) (2-2444) | (Code Rank: 7)<br>Me siento frustrada por no poder salir y por tener problemas intrafamiliares. (I am frustrated that I can't go out and that I have family problems.)<br>(2-2292) | (Code Rank: 8)<br>Job is gone...substitute teacher. Can't go to church. [My] mother is in a rest home, can't visit her. Can't see my daughter or granddaughter. Wish I could just quit worrying about dying or my at-risk husband dying unexpectedly, horribly, suffering, not being able to be together. I think this is like PTSD...living on the edge of not knowing. I'm sure there will be consequences of what we are witnessing. (1-2635) | (Code Rank: 5)<br>Unable to visit mother & aunt in aged care. Unable to visit other family members. Worry of contracting virus and dying prematurely.<br>(2-1357) |
|  | Africa | Asia | Europe | Latin America | Northern America | Oceania |
| **8. Cleaning supplies**<br>(1256; 18.3%: 17.4%, 19.2%) | (Code Rank: 10)<br>Manque de produits (bavettes,gel etc ). Si ça se trouve,très cher.<br>(Lack of products (masks, gel, etc). If it is [available], [it's] very expensive.)<br>(2-1397) | (Code Rank: 9)<br>Most shops have been closed and the price has been increased than before.<br>(1-993) | (Code Rank: 8)<br>There wasn't a drop in the county of hand sanitizer. It was like gold dust.<br>(1-1271) | (Code Rank: 10)<br>No tener agua suficiente para aseo de manos y superficies. Ta angustias no poder adquirir alcohol o gel antibacterial. Y sobre todo el confinamiento o cuarentena. Te aburres. Asi hagas muchas cosas. No estamos acostumbrados a estar 24 horas en casa todos los día. (Not having enough water to clean hands and surfaces. It's anguishing not being able to buy alcohol or antibacterial gel. And especially the confinement or quarantine. You get bored So do many things. We are not used to being at home 24 hours every day.)<br>(1-4164) | (Code Rank: 6)<br>When we do go to the grocery store we have to suit up like we're going into some kind of horrible situation. And then the shelves are empty. I cannot find anything to disinfect with because people have hoarded it. I could go on and on about the ways it has affected me but mostly it has made me very sad.<br>(1-2628) | (Code Rank: 8)<br>When the country was in panic mode - all the hand sanitiser, soap and toilet paper was bought - but the flow is getting better (although shelves are still empty sometimes)<br>(2-1586) |
| **9. Shortages**<br>(1042; 15.2%: 14.3%, 16.0%) | (Code Rank: 21)<br>Rupture de stock puis hausse des prix apres reapparition des articles.<br>(Out of stock then prices increase after reappearance of items) | (Code Rank: 13)<br>We live in a village in [ ]. There are no shops around. As a result, it is difficult to get essential commodities such as rice, pulse oil and other products. | (Code Rank: 9)<br>La gente terrorizzata inizialmente ha depredato i supermercati che non sono riusciti a rimpiazzare i prodotti mancanti.<br>(Terrified people initially looted supermarkets which | (Code Rank: 15)<br>Debido a las compras de panico, mucjas cosas se agotaron o subieron de precio. Ejemplo.el.huevo. (Due to panic purchases, many things sold out or went up in price, for example, eggs) | (Code Rank: 9)<br>It's been very difficult to get necessities in the local stores because what little stock they have coming in disappears almost immediately.<br>(2-3) | (Code Rank: 9)<br>No hand sanitizer anywhere for weeks, no soap, ended up using body wash at ten dollars a bottle!<br>(2-601) |

|  |  |  |  |  |  |  |
| --- | --- | --- | --- | --- | --- | --- |
| Respondents with Code Applied<br>(n; %:95% CI) | Exemplary quote, in respondent's language (with respondent number) |  |  |  |  |  |
|  | Africa | Asia | Europe | Latin America | Northern America | Oceania |
|  | (2-1581) | (1-182) | failed to replace the missing products)<br>(2-1737) | (2-2211) |  |  |
| **10. Unemployment**<br>(1039; 15.1%: 14.3%, 16.0%) | (Code Rank: 8)<br>No jobs and money and NO FREEDOM.<br>(2-871) | (Code Rank: 8)<br>We had to shut down our restaurant<br>(1-1751) | (Code Rank: 10)<br>Je n'arrive pas à trouver un emploi.<br>(I can't find a job)<br>(2-1443) | (Code Rank: 8)<br>Nessun lavoro, nessun reddito per comprare cibo.<br>(no work, no income to buy food)<br>(1-3439) | (Code Rank: 11)<br>My job is "on hold" until this is over.<br>(1-333) | (Code Rank: 11)<br>I have 3 young adult sons living here - 2 lost their jobs due to coronavirus. We can't spend time doing the things we normally do with people we normally see. Everyone is stressed.<br>(1-40) |

Table 6 presents the results of qualitative code clustering to accomplish dimension reduction and to aggregate similar codes together. The original 80 qualitative codes (parent, child, and grandchild codes) were reduced to 26 clusters with of the most representative variable and total proportion of variation explained by each cluster identified. Overall, the proportion of variation explained by clustering was 49%, with dispersion of the primary dominant clusters across the domains and levels of the medical ecological model. For example, two clusters in the “abiotic” domain (“Struggle to access cleaning supplies” primarily functioning at the household level and “Change in work to remote” primarily functioning at the individual level), two clusters in the “biological” domain (“Worried while pregnant” and “Worse mental health” both primarily functioning at the individual domain), and three clusters in the “sociocultural” domain at the community level (“Government preparedness,” “Need to improve current systems,” and “World will not be the same/ loss of life”) all reflect the multidimensional, multileveled nature of lived experience during COVID-19. Further, most clusters contain other qualitative codes in addition to the most representative code that reflect other levels and domains, illustrating the interface – in reality – among different types of dimensions of lived experiences. Respondents frequently indicated this intersection of distinct, but connected, phenomena in describing their experience; for example:

> It causes me anxiety because a lot of people do not follow the recommended measures… The measures [the government] introduced were strict but came late. The way they did it caused people outrage so a lot of people ignore them out of spite for the government and mistrust… I feel scared to go out because I know a lot of people don’t care, and a lot of people cover up their symptoms… I feel anxious to work, I cannot do my best at work. Also, I work over the internet so I can’t work as much because the internet connection is very unstable now… I don’t go out at all… I feel very stressed. (Central Europe woman in her 30s; #1-2034)

**Table 6:** Qualitative Code Cluster with Most Representative Code, Proportion of Variation Explained, and Medical Ecological Levels and Domains.

|  |  |  |  |  |  |
| --- | --- | --- | --- | --- | --- |
| Variables Included in Cluster | Most Representative Variable (Cluster ID) | Medical Ecological Level | Medical Ecological Domain | Total Proportion of Variation Explained | Exemplary Quote |
| Struggle to access cleaning supplies (Code 6.e)<br>Supply shortages (Code 6.e.1)<br>Struggle to basic goods and services (Code 6)<br>Price gouging of supplies (Code 6.e.3) | Struggle to access cleaning supplies (Code 6.e) | Household | Abiotic | 0.033 | There was a shortage of masks & sanitisers due to bulk buying as the virus entered our country. After really long wait, purchased sanitiser from an online grocery store at three times of the real price. (1-779) |
| Worried while pregnant (Code 22.b.3)<br>Pregnancy (Code 22.b)<br>Health Effects of newborns (Code 22.b.4) | Worried while pregnant (Code 22.b.3) | Individual | Biological | 0.025 | Being pregnant, it's a constant thought, there is limited research on how COVID19 affects newborns and pregnant women. Affects more of my mental health. [Stress] increased due to being worried about catching the virus being pregnant. (1-2473) |
| Government preparedness (Code 12)<br>Government unprepared (Code 12.a)<br>No freedoms/activities/plans (Code 2) | Government preparedness (Code 12) | Community | Sociocultural | 0.023 | No implementaron politicas de prevencion. No equiparon a los trabajadores de la salud para que estos se protegieran. No hicieron nada para un mejor control y propagación de la pandemia. Expusieron ynsiguen exponiendo a personas al contagion (They did not implement prevention policies. They did not equip health workers to protect themselves. They did nothing to better control and spread the pandemic. They exposed and continue to expose people to contagion) (2-2037) |
| Variables Included in Cluster | Most Representative Variable (Cluster ID) | Medical Ecological Level | Medical Ecological Domain | Total Proportion of Variation Explained | Exemplary Quote |
| Worse Mental Health (Code 8)<br>Stressed (Code 8a) | Worse Mental Health (Code 8) | Individual | Biological | 0.023 | Une famille qui doit gerer des enfants, le stress lie a la perte de revenu pour le menage; le risque de violence conjugale et face aux enfants, beaucoup de facteurs qui favorisent un glissement psychologique... (A family that has to manage children, the stress linked to the loss of income for the household; the risk of spousal and child violence, many factors that promote a psychological slide ...) (1-3384) |
| Change in work to remote (Code 7.b)<br>Difficult to work remotely (Code 7.b.2)<br>Facilitated through technology (Code 7.b.1)<br>Changes in exercise (Code 2.a) | Change in work to remote (Code 7.b) | Individual | Abiotic | 0.022 | Le fait de travailler à la maison mettait pas mal de contrainte. Techniquement, il était possible de travailler mais il n'y a plus de contact physique avec les collègues ce qui gêne l'avancement des projets, plus que ce que j'avais pensé. En voyant des actions qui n'avançaient pas et en addition du manque de sport, je pense que c'est la combinaison de ces deux choses qui a pesé sur ma santé mentale. (Working at home was a lot of stress. Technically, it was possible to work, but there is no longer any physical contact with colleagues, which hinders the progress of projects, more than I had expected. Seeing actions that were not progressing and in addition to the lack of sport, I think it was the combination of these two things that took a toll on my sanity.) (2-1482) |
| Changes in work/ employment (Code 7)<br>Unemployment (Code 7.c)<br>Reduced pay or work hours (Code 7.f)<br>Less business at work (Code 7.a)<br>Quit for safety reasons (Code 7.e) | Changes in work/ employment (Code 7) | Individual | Sociocultural | 0.022 | Estoy en casa encerrada, no tengo trabajo... Trabajo por comision, si no vendo no hay ingresos... Que no estoy trabajando, pero el gobierno no ha exigido que los banco y demas instituciones crediticias sean benevolentes con los deudores. Si teniendo que pagar alquiler, prestamos, servicios. (I'm locked up at home, I don't have a job... I work on commission, if I don't sell there is no income... I'm not working, but the government has not demanded that banks and other credit institutions be benevolent with debtors. If having to pay rent, loans, services.) (2-2172) |
| Need to improve current systems/learn (Code 13)<br>See less environmental degradation (Code 13.a) | Need to improve current systems/learn (Code 13) | Community | Sociocultural | 0.021 | Spero che questa situazione ci insegni a cambiare tante nostre cattive abitudini, a rispettare di più gli altri e la natura e non avere il solo denaro come scopo nella vita, esistono tantissimi altri valori molto più importanti per condurre una vita sana e di principi. (I hope this situation teaches us to change |
| Variables Included in Cluster | Most Representative Variable (Cluster ID) | Medical Ecological Level | Medical Ecological Domain | Total Proportion of Variation Explained | Exemplary Quote |
| Not let economy be over health /<br>Need more investment in health<br>(Code 13.b) |  |  |  |  | many of our bad habits, to respect others and nature more and not to have only money as a purpose in life, there are many other values that are much more important for leading a healthy and principled life.) (1-3664) |
| Struggle to access food (Code 6.a)<br>Food shortages (Code 6.a.1)<br>Not enough income to buy goods and services (Code 6.d) | Struggle to access food (Code 6.a) | Household | Sociocultural | 0.021 | I have no income and no way to pay my bills until the world reopens. Still need to eat. I hate living off the charity of others... We live on a small island that depends on food to be brought in. We have not had a boat in months, prices are increasing and I have no money. (2-645) |
| Loss of life (Code 14.a)<br>World will not be the same (Code 14)<br>Conspiracy Theory (Code 19) | World will not be the same/ Loss of life (Code 14.a) | Community | Sociocultural | 0.02 | Lo que me entristece es esos países donde hay tanto contagio y muertes y el susto de que se repita aquí. Es doloroso ver hijos con padres muertos, padres con hijos muertos, y esos enfermos sin poder recibir tratamiento. (What saddens me is those countries where there is so much contagion and deaths and the fright that it will be repeated here. It is painful to see children with dead parents, parents with dead children, and those sick without being able to receive treatment.) (2-2296) |
| Primarily unaffected/coping (Code 9)<br>Following prevention guidelines (Code 9.c)<br>Faith (Code 9.a) | Primarily unaffected/coping (Code 9) | Individual | Sociocultural | 0.019 | No impact. I have faith and go for walks in fresh air staying 2 meters away from people not in my bubble... Can't go walking or coffee with friends. Can't see my grandchildren. Hoping to go to level 2 asap so can do these things...{The} government has let people down in keeping people away from seeing dying relatives. So cruel. (2-939) |
| Societal disruption - differential impacts across countries (Code 10.a)<br>Societal disruption (Code 10)<br>Government prepared (Code 12.b) | Societal disruption - differential impacts across countries (Code 10.a) | Community | Sociocultural | 0.018 | I don't have too much worry in my own country because I think we have handled it well, but I worry about countries like [ ] who are struggling to follow directives and putting others at risk. (2-186) |
| Worried about loved ones getting sick (Code 8.a.4)<br>Worried about getting sick (Code 8.a.6) | Worried about loved ones getting sick (Code 8.a.4) | Individual | Biological | 0.018 | Lo stress mi ha lasciato profondamente preoccupato per la salute di molte persone, la possibile morte di persone vicine a me e la mia stessa morte...Sono in quarantena. La mia vita sociale è limitata, non posso entrare all'università e non posso lavorare fuori. Voglio solo essere più forte per gestire tutto. (The stress has left me deeply concerned about the health of many people, the possible death of people close to me and my own death ... I |
| Variables Included in Cluster | Most Representative Variable (Cluster ID) | Medical Ecological Level | Medical Ecological Domain | Total Proportion of Variation Explained | Exemplary Quote |
| Preexisting medical conditions and/ or elderly that increases vulnerability (Code 8.a.10)<br>Worried about losing loved ones (Code 8.a.5)<br>Disturbed sleep (Code 8.b) |  |  |  |  | am in quarantine. My social life is limited, I cannot enter university and I cannot work outside. I just want to be stronger to handle everything.) (1-3412) |
| Worries or concerns about childcare (Code 22.a)<br>Maternal and child health worries and concerns (Code 22) | Worries or concerns about childcare (Code 22.a) | Household | Sociocultural | 0.018 | Au debut tous les magasins y compris des vivres et le marché étaient fermés. Plus tard la situation s'est assouplie et on pouvait accéder aux magasins de vivres durant la moitié de la journée. Plus tard un accès limité au marché devenait aussi possible et l'accès au magasin était plus élargie. On est au dernier trimestre de grossesse de ma femme au début du confinement. Durant le confinement nous avons pu s'organiser clandestinement avec d'autres pour pouvoir acheter l'essentiel pour le futur bébé en cas où le confinement soit encore si stricte lors de l'accouchement (éventuellement prématuré). (At the beginning all stores including food and the market were closed. Later, the situation eased up and we could access the food stores for half the day. Later, limited access to the market also became possible and access to the store was widened. We are in the last trimester of my wife's pregnancy at the start of lockdown. During the lockdown, we were able to organize clandestinely with others to be able to buy the essentials for the future baby in the event that the confinement is still so strict during childbirth (possibly premature). (2-1532) |
| Panic buying of supplies (Code 6.e.2)<br>Panic buying of food (Code 6.a.2)<br>Pre-existing mental health condition exacerbated (Code 8.f) | Panic buying of supplies (Code 6.e.2) | Individual | Sociocultural | 0.017 | Because the PIG HOG panic buyers got the lot. We nearly starved the first 2 weeks! (2-307) |
| Uncertainty of loss of life (Code 15.b)<br>Uncertain Future (Code 15) | Uncertainty of loss of life (Code 15.b) | Individual | Sociocultural | 0.017 | La falta de control de la situación genera mucho estrés ... La burocracia y la ineptitud afecta grandemente al país, a sus habitantes y al proceso de control de contagio...Lo que más preocupa es la despreocupación de tantas personas en la calle y lugares públicos que no se protegen y ponen en riesgo |
| Variables Included in Cluster | Most Representative Variable (Cluster ID) | Medical Ecological Level | Medical Ecological Domain | Total Proportion of Variation Explained | Exemplary Quote |
| Hope for an end to the pandemic (Code 17)<br>Origin of pandemic uncertain (Code 18) |  |  |  |  | la salud y la vida de los demás. La interacción familiar y social, la cotidianidad y los hábitos de vida, todo se ha trastocado. Cuando la cosas no están bajo nuestro control y la salud y la vida se ven amenazadas por un virus tan letal genera mucha incertidumbre, tristeza y impotencia. (The lack of control of the situation generates a lot of stress ... The bureaucracy and ineptitude greatly affect the country, its inhabitants and the contagion control process ... What worries the most is the carelessness of so many people on the street and public places that are not protected and put the health and lives of others at risk. Family and social interaction, daily life and habits of life, everything has been upset. When things are not under our control and health and life are threatened by such a lethal virus, it generates a lot of uncertainty, sadness and helplessness.) (2-2507) |
| Hard taking care of family (Code 1.a)<br>Staying home/social distancing/ quarantine (Code 1)<br>Social disruption - changes in education (Code 10.b) | Hard taking care of family (Code 1.a) | Household | Sociocultural | 0.016 | Me preocupa algo la situación, pero una cantidad normal y asumible. El estar confinados, especialmente al tener niñas pequeñas, puede causar algo de estrés, pero creo que también manejable, excepto en momentos puntuales... Al tener que trabajar, atender a los niños, la casa... organizarlo todo (lo cual es una tarea que suele recaer sobre las mujeres)... he tenido momentos de estrés, sobre todo por la atención constante que reclaman las niñas, cosa natural debido a su corta edad - 4 años y 6 meses. (I am somewhat concerned about the situation, but a normal and manageable amount. Being confined, especially when having little girls, can cause some stress, but I think it is also manageable, except in specific moments ... When having to work, take care of the children, the house ... organize everything (which is a chore which usually falls on women) ... I have had moments of stress, especially due to the constant attention that girls demand, a natural thing due to their young age - 4 years and 6 months.) (1-4056) |
| Stressed about work (Code 8.2.a)<br>Stressed about finances (Code 8.1.a)<br>Stressed - far from loved ones (Code 8.7.a) | Stressed about work (Code 8.2.a) | Individual | Sociocultural | 0.015 | Non posso lavorare fuori come hostess quindi non ricevo più un reddito... Non sono riuscita a comprare molte cose non avendo soldi e mezzi per andare al supermercato. (I cannot work outside as a hostess so I no longer receive an income... I have not been able to buy many things, not having the money and means to go to the supermarket.) (1-3748) |
| Variables Included in Cluster | Most Representative Variable (Cluster ID) | Medical Ecological Level | Medical Ecological Domain | Total Proportion of Variation Explained | Exemplary Quote |
| Struggle to Access Non-COVID-19 medical services (Code 6.b)<br>Loss of travel (code 5)<br>Struggle to access COVID-19 medical equipment, PPE, services, and personnel (Code 6.c) | Struggle to Access Non-COVID-19 medical services (Code 6.b) | Individual | Health care | 0.015 | Je devrais me rendre dans un centre de pasteur pour une consultation gastro-enterologique mais la pandémie est venu m'empêcher maintenant je continue à souffrir. (I should go to a center for a gastroenterologic consultation but the pandemic prevents me, now I continue to suffer.) (2-1460) |
| Stressed because people are not following guidelines (Code 8.a.12)<br>Essential worker (Code 7.d)<br>Using PPE (Code 3) | Stressed because people are not following guidelines (Code 8.a.12) | Individual | Sociocultural | 0.014 | I am an "essential worker" and have to work with customers everyday. Not a lot of people are taking it seriously. Work has changed dramatically. Haven't seen family in weeks. Protective shield supplied by work, only a few are wearing them. Dr. Appointments changed to telephone. Surgery cancelled as it is not life threatening. Due to anxiety, skipped work some days. No wipes, toilet paper, or paper towel. But able to get other items, a lot is sold out. (1-468) |
| Bored (Code 8.h)<br>Depressed (Code 8.d) | Bored (Code 8.h) | Individual | Sociocultural | 0.013 | I'm feeling more anxious and sad, more bored and it's like being in a cage, and like the world is about to end. (1-3466) |
| Variables Included in Cluster | Most Representative Variable (Cluster ID) | Medical Ecological Level | Medical Ecological Domain | Total Proportion of Variation Explained | Exemplary Quote |
| Economic decline (Code 16)<br>Frustration (Code 8.e) | Economic decline (Code 16) | Community | Sociocultural | 0.013 | The [ ] government is working hard but I believe the lockdown measures if not relaxed soon will cause socio-economic issues far worse than Covid 19. (1-2282) |
| Limited coping mechanisms (Code 8.i)<br>Isolation / Lack of social support (Code 4) | Limited coping mechanisms (Code 8.i) | Individual | Sociocultural | 0.013 | En mi caso la rutina que tenía ha sido trastocada. El cambio al plan de continuidad de las universidades, las nuevas exigencias, la incertidumbre... eso provoca ansiedad y estrés...Acostumbrarme demasiado a estar encerrada en casa y luego tener agorafobia. Universidad online, estudiar en casa en vez de en una sala de lectura o biblioteca. No poder distraerme con amigos o socializar. CAMBIO DE RUTINA TOTAL. ansiedad por la incertidumbre, sobre como gestionan las cosas en la universidad (aplazándolo todo y acumulándose luego, mayor dificultad en los exámenes, etc.) (In my case the routine I had has been upset. The change to the university continuity plan, the new demands, the uncertainty ... that causes anxiety and stress ... Getting too used to being locked up at home and then having agoraphobia. Online university, study at home instead of in a reading room or library. Not being able to distract myself with friends or socialize. TOTAL ROUTINE CHANGE. anxiety about uncertainty, about how things are managed at the university (putting everything off and accumulating later, greater difficulty in exams, etc.) (1-3988) |
| Variables Included in Cluster | Most Representative Variable (Cluster ID) | Medical Ecological Level | Medical Ecological Domain | Total Proportion of Variation Explained | Exemplary Quote |
| Sometimes stressed (Code 8.c)<br>Outbreak re-occurrence/ endless (Code 15.a) | Sometimes stressed (Code 8.c) | Individual | Sociocultural | 0.013 | Moi personnellement le fait de rester à la maison crée un grand vide pour. Je ne suis plus avec mes amis, mes camarades. En tout la solitude me stresse beaucoup. Aussi le fait que chez nous les gens ne respecte pas les mesures barrières m'ennerve et me rends impuissant craignant un rebond de la maladie. (Me personally the fact of staying at home creates a great void. I am no longer with my friends, my comrades. In all, loneliness stresses me a lot. Also the fact that with us people do not respect the barrier measures irritates me and makes me powerless fearing a rebound of the disease.) (2-1454) |
| Stigma/ Bullying (Code 23)<br>Misinformation (Code 11) | Stigma/ Bullying (Code 23) | Individual | Sociocultural | 0.013 | Discrimination in public when wearing nursing scrubs. Educating others online who spread misinformation has caused unnecessary stress. (1-2788) |
| Stressed about government action (Code 8.a.9)<br>Need collaborative action (Code 12.c) | Stressed about government action (Code 8.a.9) | Individual | Sociocultural | 0.013 | Sono ansioso e pessimista (e il panorama politico in {*} non aiuta molto). Il governo {*} ha una mentalità zero, nessuna politica sociale e ha negato la malattia sin dall'inizio (anche con {*} infetto da covid-19)... Non ho entrate, non ci sono previsioni per un ritorno all'economia e il governo {*} sta rapidamente crollando. ..È una pandemia grave e rimarrà nella società per gli anni a venire. Dobbiamo reinventarci adesso, o non saremo in grado di far avanzare la società. (I'm anxious and pessimistic (and the political landscape doesn't help much)... The government has a zero mindset, no social policy and has denied the disease from the start (even when covid-19 infected {the leader}) ... I have no income, there is no forecast for a return to the economy and the government is rapidly collapsing...It is a serious pandemic and will remain in society for years to come. We have to reinvent ourselves now, or we won't be able to move society forward) (1-3501) |
| Variables Included in Cluster | Most Representative Variable (Cluster ID) | Medical Ecological Level | Medical Ecological Domain | Total Proportion of Variation Explained | Exemplary Quote |
| Stressed about news (Code 8.a.8)<br>Stress worsening the immune system (Code 8.a.11) | Stressed about news (Code 8.a.8) | Individual | Sociocultural | 0.013 | Produce estress al encierro, al miedo a contagiarse, a la desinformacion, notocias falsas, informacion contradictoria en todos los medios y produce disminución al sistema inmunológico... muy contradictorias las noticias ademas les interesa mas la nota roja que labrealidad de la mayoria. demasiadas notivias falsas y bots politicos que nadie ha podido controlar. Hay que saber buscar los espacios con informacion veraz. (It produces stress to confinement, fear of infection, misinformation, false news, contradictory information in all media and produces a decrease in the immune system ... the news is very contradictory, and they are more interested in sensationalism than in the majority of workers. Too many fake news and political bots that nobody has been able to control. You have to know how to find the spaces with truthful information.) (2-2143) |

In this instance, the respondent links their anxiety (*Worse Mental Health (Code 8)*) to lack of adherence to prevention measures (*Stressed because people are not following guidelines (Code 8.a.12)*), brought about by lack of timely government preparation (*Government preparedness (Code 12*), compounded by an inability to work effectively from home (*Change in work to remote (Code 7.b*). In so doing, this respondent illustrates the simultaneous experience of levels and domains of the medical ecological model (e.g., anxiety (biological and individual), government preparedness (community and sociocultural), technology functionality (abiotic and household)) in relaying her experience. Similarly, a South American man in his 20s conveys a similar sense of complexity in lived experience that reflects the dynamic intersection of numerous qualitative codes and levels/ domains of the medical ecological model:

> *Me he sentido triste y preocupado pues mi situación económica va a empeorar debido al confinamiento…enfermar de algo mas y no poder recibir atención médica…tengo un negocio propio que en cuestión de días desaparecerá pues no tengo dinero para mantenerlo…mi negocio va a desaparecer pues debo pagar arriendo y no tengo dinero…Me he sentido mal, estoy muy preocupado porque el dinero para comer se agota. Pienso que los gobiernos deberían dar ayudas a todas las personas para la alimentación pues sin trabajar es dificil obtener dinero y sobrevivir.* (I have felt sad and worried because my economic situation is going to worsen due to the shutdown … to get sick from something else and not be able to receive medical attention … I have my own business that will disappear in a matter of days because I do not have money to support it…my business is going to disappear because I have to pay rent and I have no money… I have felt bad, I am very worried because the money to eat is running out. I think that governments should give aid to all people for food because without working it is difficult to obtain money and survive.) (South America man in his 20s; #1-3895)

### Multivariate, Multilevel Analysis

Mentioned previously, only those quantitative variables with <10% missing data and <5 VIF were included in the multivariate models. Table 7 shows heat maps of variable coefficients that were significantly (p<.05) associated with the Impact Score and its four component scores, by six global regions. Variables from all three levels of the ecological model remained significant in the multilevel, multivariate analysis.

**Table 7a:**
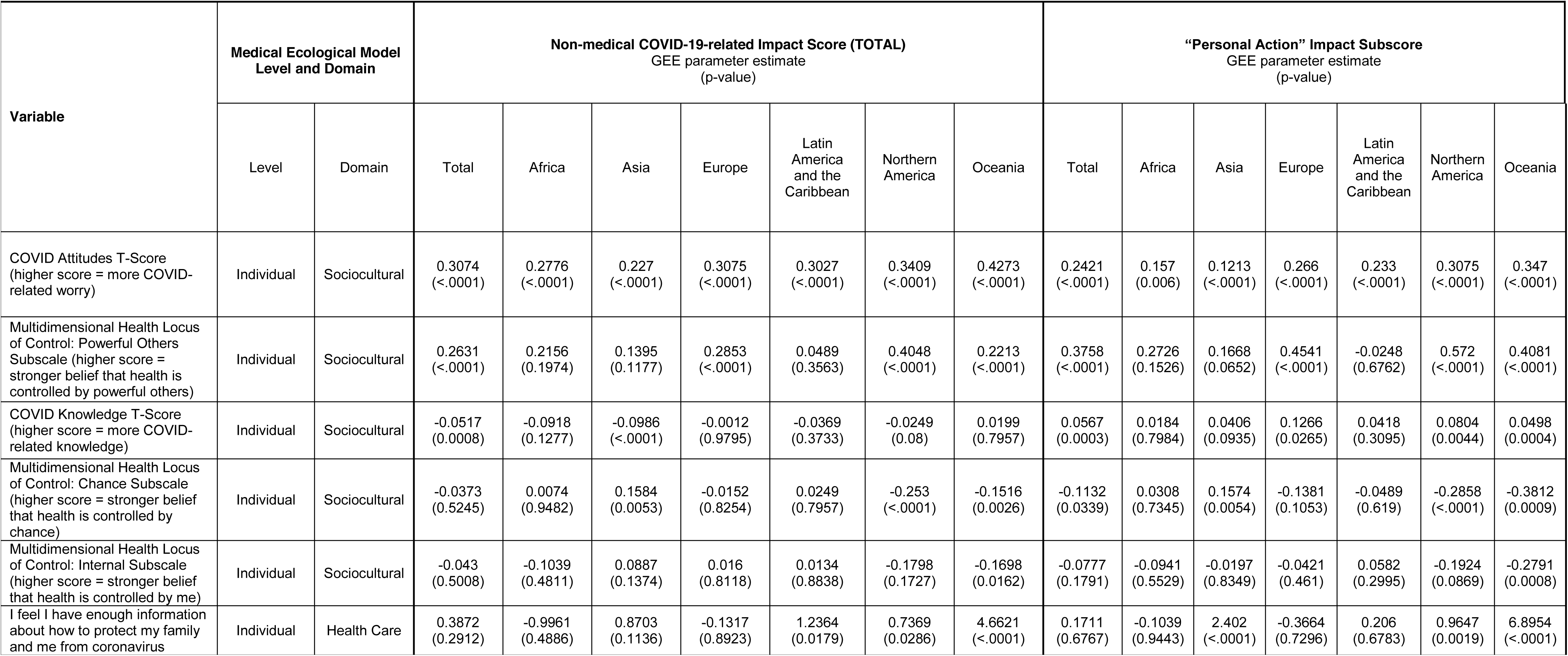
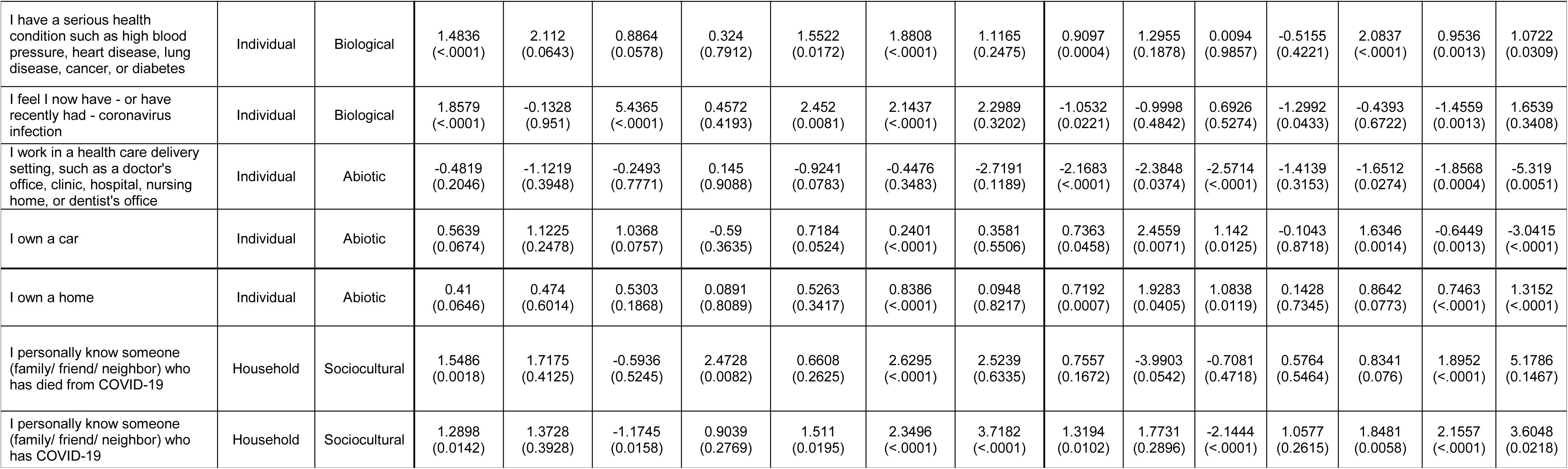
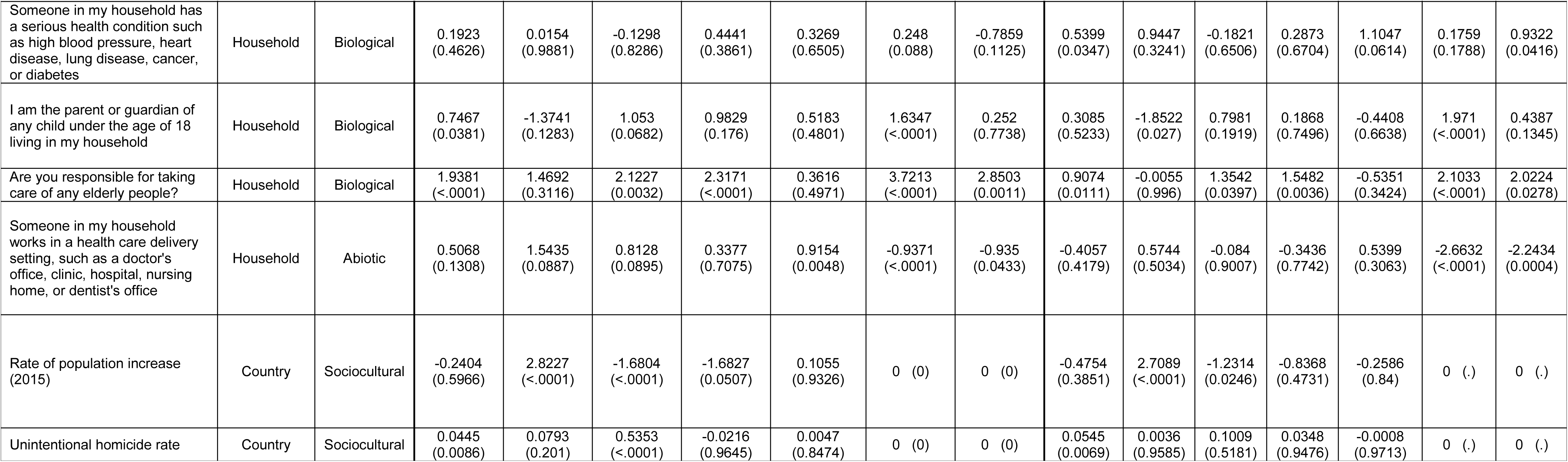
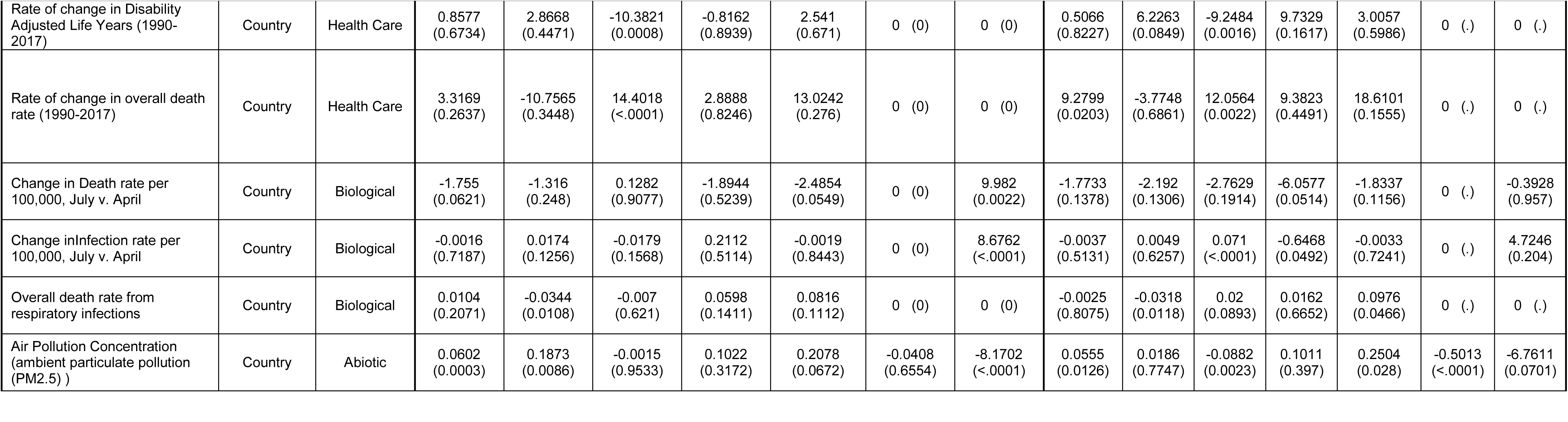
Probability of association (p-value of GEE coefficient) between Medical Ecological Variables with non-medical COVID-19-related Impact Score and Four Subscores.

|  |  |  |  |  |  |  |  |  |  |  |  |  |  |  |  |  |
| --- | --- | --- | --- | --- | --- | --- | --- | --- | --- | --- | --- | --- | --- | --- | --- | --- |
| Variable | Medical Ecological Model Level and Domain |  | Non-medical COVID-19-related Impact Score (TOTAL)<br>GEE parameter estimate<br>(p-value) |  |  |  |  |  |  | “Personal Action” Impact Subscore<br>GEE parameter estimate<br>(p-value) |  |  |  |  |  |  |
|  | Level | Domain | Total | Africa | Asia | Europe | Latin America and the Caribbean | Northern America | Oceania | Total | Africa | Asia | Europe | Latin America and the Caribbean | Northern America | Oceania |
| COVID Attitudes T-Score<br>(higher score = more COVID-related worry) | Individual | Sociocultural | 0.3074<br>(<.0001) | 0.2776<br>(<.0001) | 0.227<br>(<.0001) | 0.3075<br>(<.0001) | 0.3027<br>(<.0001) | 0.3409<br>(<.0001) | 0.4273<br>(<.0001) | 0.2421<br>(<.0001) | 0.157<br>(0.006) | 0.1213<br>(<.0001) | 0.266<br>(<.0001) | 0.233<br>(<.0001) | 0.3075<br>(<.0001) | 0.347<br>(<.0001) |
| Multidimensional Health Locus of Control: Powerful Others Subscale (higher score = stronger belief that health is controlled by powerful others) | Individual | Sociocultural | 0.2631<br>(<.0001) | 0.2156<br>(0.1974) | 0.1395<br>(0.1177) | 0.2853<br>(<.0001) | 0.0489<br>(0.3563) | 0.4048<br>(<.0001) | 0.2213<br>(<.0001) | 0.3758<br>(<.0001) | 0.2726<br>(0.1526) | 0.1668<br>(0.0652) | 0.4541<br>(<.0001) | -0.0248<br>(0.6762) | 0.572<br>(<.0001) | 0.4081<br>(<.0001) |
| COVID Knowledge T-Score<br>(higher score = more COVID-related knowledge) | Individual | Sociocultural | -0.0517<br>(0.0008) | -0.0918<br>(0.1277) | -0.0986<br>(<.0001) | -0.0012<br>(0.9795) | -0.0369<br>(0.3733) | -0.0249<br>(0.08) | 0.0199<br>(0.7957) | 0.0567<br>(0.0003) | 0.0184<br>(0.7984) | 0.0406<br>(0.0935) | 0.1266<br>(0.0265) | 0.0418<br>(0.3095) | 0.0804<br>(0.0044) | 0.0498<br>(0.0004) |
| Multidimensional Health Locus of Control: Chance Subscale (higher score = stronger belief that health is controlled by chance) | Individual | Sociocultural | -0.0373<br>(0.5245) | 0.0074<br>(0.9482) | 0.1584<br>(0.0053) | -0.0152<br>(0.8254) | 0.0249<br>(0.7957) | -0.253<br>(<.0001) | -0.1516<br>(0.0026) | -0.1132<br>(0.0339) | 0.0308<br>(0.7345) | 0.1574<br>(0.0054) | -0.1381<br>(0.1053) | -0.0489<br>(0.619) | -0.2858<br>(<.0001) | -0.3812<br>(0.0009) |
| Multidimensional Health Locus of Control: Internal Subscale (higher score = stronger belief that health is controlled by me) | Individual | Sociocultural | -0.043<br>(0.5008) | -0.1039<br>(0.4811) | 0.0887<br>(0.1374) | 0.016<br>(0.8118) | 0.0134<br>(0.8838) | -0.1798<br>(0.1727) | -0.1698<br>(0.0162) | -0.0777<br>(0.1791) | -0.0941<br>(0.5529) | -0.0197<br>(0.8349) | -0.0421<br>(0.461) | 0.0582<br>(0.2995) | -0.1924<br>(0.0869) | -0.2791<br>(0.0008) |
| I feel I have enough information about how to protect my family and me from coronavirus | Individual | Health Care | 0.3872<br>(0.2912) | -0.9961<br>(0.4886) | 0.8703<br>(0.1136) | -0.1317<br>(0.8923) | 1.2364<br>(0.0179) | 0.7369<br>(0.0286) | 4.6621<br>(<.0001) | 0.1711<br>(0.6767) | -0.1039<br>(0.9443) | 2.402<br>(<.0001) | -0.3664<br>(0.7296) | 0.206<br>(0.6783) | 0.9647<br>(0.0019) | 6.8954<br>(<.0001) |
| Variable | Medical Ecological Model Level and Domain |  | Non-medical COVID-19-related Impact Score (TOTAL)<br>GEE parameter estimate<br>(p-value) |  |  |  |  |  |  | “Personal Action” Impact Subscore<br>GEE parameter estimate<br>(p-value) |  |  |  |  |  |  |
|  | Level | Domain | Total | Africa | Asia | Europe | Latin America and the Caribbean | Northern America | Oceania | Total | Africa | Asia | Europe | Latin America and the Caribbean | Northern America | Oceania |
| I have a serious health condition such as high blood pressure, heart disease, lung disease, cancer, or diabetes | Individual | Biological | 1.4836<br>(<.0001) | 2.112<br>(0.0643) | 0.8864<br>(0.0578) | 0.324<br>(0.7912) | 1.5522<br>(0.0172) | 1.8808<br>(<.0001) | 1.1165<br>(0.2475) | 0.9097<br>(0.0004) | 1.2955<br>(0.1878) | 0.0094<br>(0.9857) | -0.5155<br>(0.4221) | 2.0837<br>(<.0001) | 0.9536<br>(0.0013) | 1.0722<br>(0.0309) |
| I feel I now have - or have recently had - coronavirus infection | Individual | Biological | 1.8579<br>(<.0001) | -0.1328<br>(0.951) | 5.4365<br>(<.0001) | 0.4572<br>(0.4193) | 2.452<br>(0.0081) | 2.1437<br>(<.0001) | 2.2989<br>(0.3202) | -1.0532<br>(0.0221) | -0.9998<br>(0.4842) | 0.6926<br>(0.5274) | -1.2992<br>(0.0433) | -0.4393<br>(0.6722) | -1.4559<br>(0.0013) | 1.6539<br>(0.3408) |
| I work in a health care delivery setting, such as a doctor's office, clinic, hospital, nursing home, or dentist's office | Individual | Abiotic | -0.4819<br>(0.2046) | -1.1219<br>(0.3948) | -0.2493<br>(0.7771) | 0.145<br>(0.9088) | -0.9241<br>(0.0783) | -0.4476<br>(0.3483) | -2.7191<br>(0.1189) | -2.1683<br>(<.0001) | -2.3848<br>(0.0374) | -2.5714<br>(<.0001) | -1.4139<br>(0.3153) | -1.6512<br>(0.0274) | -1.8568<br>(0.0004) | -5.319<br>(0.0051) |
| I own a car | Individual | Abiotic | 0.5639<br>(0.0674) | 1.1225<br>(0.2478) | 1.0368<br>(0.0757) | -0.59<br>(0.3635) | 0.7184<br>(0.0524) | 0.2401<br>(<.0001) | 0.3581<br>(0.5506) | 0.7363<br>(0.0458) | 2.4559<br>(0.0071) | 1.142<br>(0.0125) | -0.1043<br>(0.8718) | 1.6346<br>(0.0014) | -0.6449<br>(0.0013) | -3.0415<br>(<.0001) |
| I own a home | Individual | Abiotic | 0.41<br>(0.0646) | 0.474<br>(0.6014) | 0.5303<br>(0.1868) | 0.0891<br>(0.8089) | 0.5263<br>(0.3417) | 0.8386<br>(<.0001) | 0.0948<br>(0.8217) | 0.7192<br>(0.0007) | 1.9283<br>(0.0405) | 1.0838<br>(0.0119) | 0.1428<br>(0.7345) | 0.8642<br>(0.0773) | 0.7463<br>(<.0001) | 1.3152<br>(<.0001) |
| I personally know someone (family/ friend/ neighbor) who has died from COVID-19 | Household | Sociocultural | 1.5486<br>(0.0018) | 1.7175<br>(0.4125) | -0.5936<br>(0.5245) | 2.4728<br>(0.0082) | 0.6608<br>(0.2625) | 2.6295<br>(<.0001) | 2.5239<br>(0.6335) | 0.7557<br>(0.1672) | -3.9903<br>(0.0542) | -0.7081<br>(0.4718) | 0.5764<br>(0.5464) | 0.8341<br>(0.076) | 1.8952<br>(<.0001) | 5.1786<br>(0.1467) |
| I personally know someone (family/ friend/ neighbor) who has COVID-19 | Household | Sociocultural | 1.2898<br>(0.0142) | 1.3728<br>(0.3928) | -1.1745<br>(0.0158) | 0.9039<br>(0.2769) | 1.511<br>(0.0195) | 2.3496<br>(<.0001) | 3.7182<br>(<.0001) | 1.3194<br>(0.0102) | 1.7731<br>(0.2896) | -2.1444<br>(<.0001) | 1.0577<br>(0.2615) | 1.8481<br>(0.0058) | 2.1557<br>(<.0001) | 3.6048<br>(0.0218) |

| Variable | Medical Ecological Model Level and Domain |  | Non-medical COVID-19-related Impact Score (TOTAL)<br>GEE parameter estimate<br>(p-value) |  |  |  |  |  |  | “Personal Action” Impact Subscore<br>GEE parameter estimate<br>(p-value) |  |  |  |  |  |  |
| --- | --- | --- | --- | --- | --- | --- | --- | --- | --- | --- | --- | --- | --- | --- | --- | --- |
|  | Level | Domain | Total | Africa | Asia | Europe | Latin America and the Caribbean | Northern America | Oceania | Total | Africa | Asia | Europe | Latin America and the Caribbean | Northern America | Oceania |
| Someone in my household has a serious health condition such as high blood pressure, heart disease, lung disease, cancer, or diabetes | Household | Biological | 0.1923<br>(0.4626) | 0.0154<br>(0.9881) | -0.1298<br>(0.8286) | 0.4441<br>(0.3861) | 0.3269<br>(0.6505) | 0.248<br>(0.088) | -0.7859<br>(0.1125) | 0.5399<br>(0.0347) | 0.9447<br>(0.3241) | -0.1821<br>(0.6506) | 0.2873<br>(0.6704) | 1.1047<br>(0.0614) | 0.1759<br>(0.1788) | 0.9322<br>(0.0416) |
| I am the parent or guardian of any child under the age of 18 living in my household | Household | Biological | 0.7467<br>(0.0381) | -1.3741<br>(0.1283) | 1.053<br>(0.0682) | 0.9829<br>(0.176) | 0.5183<br>(0.4801) | 1.6347<br>(<.0001) | 0.252<br>(0.7738) | 0.3085<br>(0.5233) | -1.8522<br>(0.027) | 0.7981<br>(0.1919) | 0.1868<br>(0.7496) | -0.4408<br>(0.6638) | 1.971<br>(<.0001) | 0.4387<br>(0.1345) |
| Are you responsible for taking care of any elderly people? | Household | Biological | 1.9381<br>(<.0001) | 1.4692<br>(0.3116) | 2.1227<br>(0.0032) | 2.3171<br>(<.0001) | 0.3616<br>(0.4971) | 3.7213<br>(<.0001) | 2.8503<br>(0.0011) | 0.9074<br>(0.0111) | -0.0055<br>(0.996) | 1.3542<br>(0.0397) | 1.5482<br>(0.0036) | -0.5351<br>(0.3424) | 2.1033<br>(<.0001) | 2.0224<br>(0.0278) |
| Someone in my household works in a health care delivery setting, such as a doctor's office, clinic, hospital, nursing home, or dentist's office | Household | Abiotic | 0.5068<br>(0.1308) | 1.5435<br>(0.0887) | 0.8128<br>(0.0895) | 0.3377<br>(0.7075) | 0.9154<br>(0.0048) | -0.9371<br>(<.0001) | -0.935<br>(0.0433) | -0.4057<br>(0.4179) | 0.5744<br>(0.5034) | -0.084<br>(0.9007) | -0.3436<br>(0.7742) | 0.5399<br>(0.3063) | -2.6632<br>(<.0001) | -2.2434<br>(0.0004) |
| Rate of population increase (2015) | Country | Sociocultural | -0.2404<br>(0.5966) | 2.8227<br>(<.0001) | -1.6804<br>(<.0001) | -1.6827<br>(0.0507) | 0.1055<br>(0.9326) | 0 (0) | 0 (0) | -0.4754<br>(0.3851) | 2.7089<br>(<.0001) | -1.2314<br>(0.0246) | -0.8368<br>(0.4731) | -0.2586<br>(0.84) | 0 (.) | 0 (.) |
| Unintentional homicide rate | Country | Sociocultural | 0.0445<br>(0.0086) | 0.0793<br>(0.201) | 0.5353<br>(<.0001) | -0.0216<br>(0.9645) | 0.0047<br>(0.8474) | 0 (0) | 0 (0) | 0.0545<br>(0.0069) | 0.0036<br>(0.9585) | 0.1009<br>(0.5181) | 0.0348<br>(0.9476) | -0.0008<br>(0.9713) | 0 (.) | 0 (.) |
| Variable | Medical Ecological Model Level and Domain |  | Non-medical COVID-19-related Impact Score (TOTAL)<br>GEE parameter estimate (p-value) |  |  |  |  |  |  | “Personal Action” Impact Subscore<br>GEE parameter estimate (p-value) |  |  |  |  |  |  |
|  | Level | Domain | Total | Africa | Asia | Europe | Latin America and the Caribbean | Northern America | Oceania | Total | Africa | Asia | Europe | Latin America and the Caribbean | Northern America | Oceania |
| Rate of change in Disability Adjusted Life Years (1990-2017) | Country | Health Care | 0.8577<br>(0.6734) | 2.8668<br>(0.4471) | -10.3821<br>(0.0008) | -0.8162<br>(0.8939) | 2.541<br>(0.671) | 0 (0) | 0 (0) | 0.5066<br>(0.8227) | 6.2263<br>(0.0849) | -9.2484<br>(0.0016) | 9.7329<br>(0.1617) | 3.0057<br>(0.5986) | 0 (.) | 0 (.) |
| Rate of change in overall death rate (1990-2017) | Country | Health Care | 3.3169<br>(0.2637) | -10.7565<br>(0.3448) | 14.4018<br>(<.0001) | 2.8888<br>(0.8246) | 13.0242<br>(0.276) | 0 (0) | 0 (0) | 9.2799<br>(0.0203) | -3.7748<br>(0.6861) | 12.0564<br>(0.0022) | 9.3823<br>(0.4491) | 18.6101<br>(0.1555) | 0 (.) | 0 (.) |
| Change in Death rate per 100,000, July v. April | Country | Biological | -1.755<br>(0.0621) | -1.316<br>(0.248) | 0.1282<br>(0.9077) | -1.8944<br>(0.5239) | -2.4854<br>(0.0549) | 0 (0) | 9.982<br>(0.0022) | -1.7733<br>(0.1378) | -2.192<br>(0.1306) | -2.7629<br>(0.1914) | -6.0577<br>(0.0514) | -1.8337<br>(0.1156) | 0 (.) | -0.3928<br>(0.957) |
| Change in Infection rate per 100,000, July v. April | Country | Biological | -0.0016<br>(0.7187) | 0.0174<br>(0.1256) | -0.0179<br>(0.1568) | 0.2112<br>(0.5114) | -0.0019<br>(0.8443) | 0 (0) | 8.6762<br>(<.0001) | -0.0037<br>(0.5131) | 0.0049<br>(0.6257) | 0.071<br>(<.0001) | -0.6468<br>(0.0492) | -0.0033<br>(0.7241) | 0 (.) | 4.7246<br>(0.204) |
| Overall death rate from respiratory infections | Country | Biological | 0.0104<br>(0.2071) | -0.0344<br>(0.0108) | -0.007<br>(0.621) | 0.0598<br>(0.1411) | 0.0816<br>(0.1112) | 0 (0) | 0 (0) | -0.0025<br>(0.8075) | -0.0318<br>(0.0118) | 0.02<br>(0.0893) | 0.0162<br>(0.6652) | 0.0976<br>(0.0466) | 0 (.) | 0 (.) |
| Air Pollution Concentration (ambient particulate pollution (PM2.5) ) | Country | Abiotic | 0.0602<br>(0.0003) | 0.1873<br>(0.0086) | -0.0015<br>(0.9533) | 0.1022<br>(0.3172) | 0.2078<br>(0.0672) | -0.0408<br>(0.6554) | -8.1702<br>(<.0001) | 0.0555<br>(0.0126) | 0.0186<br>(0.7747) | -0.0882<br>(0.0023) | 0.1011<br>(0.397) | 0.2504<br>(0.028) | -0.5013<br>(<.0001) | -6.7611<br>(0.0701) |

**Table 7b:** Probability of association (p-value of GEE coefficient) between Medical Ecological Variables with non-medical COVID-19-related Impact Score and Four Subscores.

|  |  |  |  |  |  |  |  |  |  |  |  |  |  |  |  |  |  |  |  |  |  |
| --- | --- | --- | --- | --- | --- | --- | --- | --- | --- | --- | --- | --- | --- | --- | --- | --- | --- | --- | --- | --- | --- |
|  | “Supplies” Impact Subscore<br>GEE parameter estimate<br>(p-value) |  |  |  |  |  |  | “Cancellations” Impact Subscore<br>GEE parameter estimate<br>(p-value) |  |  |  |  |  |  | “Livelihood” Impact Subscore<br>GEE parameter estimate<br>(p-value) |  |  |  |  |  |  |
| Variable | Total | Africa | Asia | Europe | Latin America and the Caribbean | Northern America | Oceania | Total | Africa | Asia | Europe | Latin America and the Caribbean | Northern America | Oceania | Total | Africa | Asia | Europe | Latin America and the Caribbean | Northern America | Oceania |
| COVID Attitudes T-Score (higher score = more COVID-related worry) | 0.1143<br>(<.0001) | 0.206<br>(<.0001) | 0.1148<br>(0.0003) | 0.0772<br>(0.0006) | 0.0804<br>(0.0004) | 0.1266<br>(<.0001) | 0.1946<br>(<.0001) | 0.1952<br>(<.0001) | 0.1348<br>(0.0079) | 0.1735<br>(<.0001) | 0.2088<br>(<.0001) | 0.1789<br>(<.0001) | 0.2085<br>(<.0001) | 0.2493<br>(<.0001) | 0.2282<br>(<.0001) | 0.2305<br>(<.0001) | 0.1902<br>(<.0001) | 0.241<br>(<.0001) | 0.2801<br>(<.0001) | 0.1867<br>(<.0001) | 0.273<br>(<.0001) |
| Multidimensional Health Locus of Control: Powerful Others Subscale (higher score = stronger belief that health is controlled by powerful others) | -0.0576<br>(0.0502) | -0.1042<br>(0.4926) | -0.0119<br>(0.8371) | -0.075<br>(0.0642) | 0.0456<br>(0.5501) | -0.1089<br>(0.0142) | -0.015<br>(0.8944) | 0.3189<br>(<.0001) | 0.3012<br>(0.0108) | 0.2116<br>(0.0766) | 0.3079<br>(<.0001) | 0.1179<br>(0.0316) | 0.522<br>(<.0001) | 0.2713<br>(<.0001) | -0.1048<br>(0.0011) | 0.0063<br>(0.9623) | -0.0226<br>(0.6953) | -0.1451<br>(0.0657) | -0.0076<br>(0.8925) | -0.16<br>(<.0001) | -0.2601<br>(0.0002) |
| COVID Knowledge T-Score (higher score = more COVID-related knowledge) | -0.03<br>(0.195) | -0.131<br>(0.0481) | -0.0537<br>(0.1111) | -0.0213<br>(0.5774) | -0.0937<br>(0.0023) | 0.0007<br>(0.9687) | 0.1612<br>(0.0167) | -0.0347<br>(0.0157) | -0.0335<br>(0.5837) | -0.0745<br>(0.0001) | 0.013<br>(0.7735) | -0.0446<br>(0.2442) | -0.0097<br>(<.0001) | 0.0349<br>(0.7395) | -0.1819<br>(<.0001) | -0.167<br>(0.0839) | -0.2288<br>(<.0001) | -0.1787<br>(<.0001) | -0.0705<br>(0.0885) | -0.1901<br>(<.0001) | -0.1963<br>(<.0001) |
| Multidimensional Health Locus of Control: Chance Subscale (higher score = stronger belief that health is controlled by chance) | 0.073<br>(0.1008) | 0.1273<br>(0.2668) | 0.036<br>(0.4889) | 0.1364<br>(0.0008) | 0.1589<br>(0.1155) | -0.0849<br>(<.0001) | -0.0013<br>(0.9875) | -0.0576<br>(0.1833) | -0.144<br>(0.1993) | 0.0327<br>(0.714) | -0.0188<br>(0.8454) | -0.0433<br>(0.6377) | -0.1627<br>(<.0001) | 0.0104<br>(0.9422) | 0.0525<br>(0.1197) | 0.0183<br>(0.8668) | 0.1596<br>(0.0001) | 0.0592<br>(0.2421) | -0.0145<br>(0.7635) | -0.0491<br>(0.0013) | 0.1592<br>(<.0001) |
|  | “Supplies” Impact Subscore<br>GEE parameter estimate<br>(p-value) |  |  |  |  |  |  | “Cancellations” Impact Subscore<br>GEE parameter estimate<br>(p-value) |  |  |  |  |  |  | “Livelihood” Impact Subscore<br>GEE parameter estimate<br>(p-value) |  |  |  |  |  |  |
| Variable | Total | Africa | Asia | Europe | Latin America and the Caribbean | Northern America | Oceania | Total | Africa | Asia | Europe | Latin America and the Caribbean | Northern America | Oceania | Total | Africa | Asia | Europe | Latin America and the Caribbean | Northern America | Oceania |
| Multidimensional Health Locus of Control: Internal Subscale (higher score = stronger belief that health is controlled by me) | -0.1048<br>(0.006) | -0.3352<br>(0.0647) | 0.1114<br>(0.0099) | -0.0355<br>(0.5841) | -0.0448<br>(0.6116) | -0.2053<br>(<.0001) | -0.3422<br>(<.0001) | 0.0462<br>(0.4074) | 0.0542<br>(0.7547) | 0.0969<br>(0.2636) | 0.1275<br>(0.0717) | -0.0255<br>(0.7305) | -0.0344<br>(0.7632) | 0.1163<br>(0.2637) | 0.0419<br>(0.4014) | -0.0091<br>(0.9664) | 0.0563<br>(0.6718) | 0.0263<br>(0.7014) | 0.0102<br>(0.9165) | 0.0141<br>(0.8882) | 0.2901<br>(<.0001) |
| I feel I have enough information about how to protect my family and me from coronavirus | 0.4502<br>(0.3335) | -1.0587<br>(0.6067) | -1.7233<br>(0.007) | -0.1175<br>(0.844) | 2.2345<br>(0.009) | -0.6842<br>(0.0014) | 0.1334<br>(0.9547) | 0.0709<br>(0.8553) | -0.673<br>(0.6109) | -0.094<br>(0.9023) | 0.8448<br>(0.5065) | -0.213<br>(0.6369) | 0.303<br>(0.008) | 4.9528<br>(0.0538) | 0.4404<br>(0.3345) | -0.8563<br>(0.6933) | 0.4894<br>(0.301) | -0.4052<br>(0.5571) | 0.833<br>(0.1947) | 1.5883<br>(0.0032) | -1.8201<br>(<.0001) |
| I have a serious health condition such as high blood pressure, heart disease, lung disease, cancer, or diabetes | 1.4391<br>(0.0066) | 1.5806<br>(0.1352) | 0.769<br>(0.331) | 1.811<br>(0.1521) | 0.0839<br>(0.921) | 2.5802<br>(<.0001) | 0.5873<br>(0.6908) | 0.8705<br>(0.0195) | 0.5832<br>(0.7406) | 1.5956<br>(<.0001) | 0.0381<br>(0.9774) | 1.4345<br>(0.0191) | 0.2599<br>(0.085) | 1.4573<br>(0.0327) | 0.4414<br>(0.2174) | 1.3915<br>(0.4175) | 0.246<br>(0.7181) | -0.26<br>(0.6682) | -0.4611<br>(0.5456) | 1.2774<br>(0.0002) | -0.1462<br>(0.833) |
| I feel I now have - or have recently had - coronavirus infection | 4.4427<br>(<.0001) | 3.4812<br>(0.2791) | 10.5516<br>(<.0001) | 2.7578<br>(<.0001) | 3.8103<br>(<.0001) | 4.031<br>(<.0001) | 2.2491<br>(0.4449) | -0.9846<br>(0.0189) | -2.5932<br>(0.2037) | -1.4699<br>(0.1943) | -1.5426<br>(0.0059) | 0.4657<br>(0.5933) | 0.215<br>(0.839) | -2.1394<br>(0.0002) | 3.5883<br>(<.0001) | -0.1934<br>(0.8977) | 5.9048<br>(<.0001) | 2.2211<br>(0.0036) | 3.6923<br>(0.0042) | 4.2874<br>(<.0001) | 3.3211<br>(0.1211) |
|  | “Supplies” Impact Subscore<br>GEE parameter estimate<br>(p-value) |  |  |  |  |  |  | “Cancellations” Impact Subscore<br>GEE parameter estimate<br>(p-value) |  |  |  |  |  |  | “Livelihood” Impact Subscore<br>GEE parameter estimate<br>(p-value) |  |  |  |  |  |  |
| Variable | Total | Africa | Asia | Europe | Latin America and the Caribbean | Northern America | Oceania | Total | Africa | Asia | Europe | Latin America and the Caribbean | Northern America | Oceania | Total | Africa | Asia | Europe | Latin America and the Caribbean | Northern America | Oceania |
| I work in a health care delivery setting, such as a doctor's office, clinic, hospital, nursing home, or dentist's office | 1.3333<br>(0.0208) | 1.5701<br>(0.1574) | 1.5723<br>(0.107) | 1.7564<br>(0.2851) | -0.4769<br>(0.5578) | 2.2686<br>(<.0001) | 1.2704<br>(0.4952) | -0.3828<br>(0.3322) | -0.6097<br>(0.6073) | -0.8141<br>(0.0293) | 0.5316<br>(0.6507) | 0.8627<br>(0.2461) | -1.1611<br>(0.0062) | -0.9198<br>(<.0001) | 0.9422<br>(0.1133) | -1.0447<br>(0.5615) | 2.5186<br>(0.0104) | -0.6552<br>(0.5355) | 0.1426<br>(0.7949) | 0.5954<br>(0.0084) | 0.345<br>(0.3187) |
| I own a car | -0.2169<br>(0.5223) | -0.9467<br>(0.2904) | 0.013<br>(0.9821) | -0.3707<br>(0.4544) | -1.4911<br>(0.0515) | 1.5331<br>(<.0001) | 0.3955<br>(0.6892) | 0.7638<br>(0.0041) | 2.027<br>(0.0465) | 1.5699<br>(<.0001) | -0.412<br>(0.3867) | 1.2551<br>(0.019) | 0.0149<br>(0.9467) | 1.9248<br>(0.296) | -0.2908<br>(0.4659) | -1.6337<br>(0.1222) | -0.7339<br>(0.3848) | -0.44<br>(0.6417) | -0.2473<br>(0.7618) | -0.4394<br>(0.1036) | 3.8187<br>(<.0001) |
| I own a home | -0.3492<br>(0.2583) | -1.4041<br>(0.2665) | -0.9162<br>(0.2366) | -0.1691<br>(0.6778) | -0.6834<br>(0.1295) | 0.746<br>(<.0001) | -1.6355<br>(0.0013) | 1.1551<br>(<.0001) | 1.2349<br>(0.0902) | 1.3811<br>(0.0031) | 0.741<br>(0.1749) | 1.0289<br>(0.092) | 1.2516<br>(<.0001) | 1.2436<br>(0.4595) | -0.7387<br>(0.0063) | -1.4538<br>(0.2182) | -0.7626<br>(0.0844) | -0.6444<br>(0.2038) | -0.223<br>(0.7408) | -0.8871<br>(0.0597) | -1.7745<br>(0.001) |
| I personally know someone (family/ friend/ neighbor) who has died from COVID-19 | 0.3821<br>(0.4351) | 5.333<br>(0.0063) | -0.7834<br>(0.3422) | 1.8288<br>(0.0741) | -1.0337<br>(0.2598) | 1.1872<br>(0.1167) | -1.4704<br>(0.6494) | 0.5183<br>(0.3629) | -0.7841<br>(0.6669) | -2.2498<br>(0.0068) | 1.2533<br>(0.3147) | 1.1487<br>(0.1262) | 1.4659<br>(<.0001) | -2.5501<br>(0.6224) | 2.3915<br>(0.0005) | 7.0329<br>(0.0974) | 2.31<br>(0.0873) | 2.9369<br>(0.1491) | 0.3007<br>(0.7709) | 2.4648<br>(<.0001) | 5.0135<br>(0.014) |
| I personally know someone (family/ friend/ neighbor) who has COVID-19 | 0.5503<br>(0.2216) | -0.3726<br>(0.8277) | 0.1572<br>(0.8359) | 0.2099<br>(0.7298) | 0.8027<br>(0.3586) | 1.2595<br>(0.0961) | -0.0524<br>(0.9608) | 0.7714<br>(0.1659) | 2.1108<br>(0.1904) | -0.954<br>(0.2608) | 0.357<br>(0.6414) | 0.232<br>(0.714) | 2.12<br>(<.0001) | 4.4102<br>(<.0001) | 0.7261<br>(0.0027) | -0.2782<br>(0.8959) | 0.9719<br>(0.0601) | 0.652<br>(0.2077) | 1.2385<br>(0.0125) | 0.3671<br>(0.1365) | 0.0098<br>(0.9917) |
|  | “Supplies” Impact Subscore<br>GEE parameter estimate<br>(p-value) |  |  |  |  |  |  | “Cancellations” Impact Subscore<br>GEE parameter estimate<br>(p-value) |  |  |  |  |  |  | “Livelihood” Impact Subscore<br>GEE parameter estimate<br>(p-value) |  |  |  |  |  |  |
| Variable | Total | Africa | Asia | Europe | Latin America and the Caribbean | Northern America | Oceania | Total | Africa | Asia | Europe | Latin America and the Caribbean | Northern America | Oceania | Total | Africa | Asia | Europe | Latin America and the Caribbean | Northern America | Oceania |
| Someone in my household has a serious health condition such as high blood pressure, heart disease, lung disease, cancer, or diabetes | 0.2296<br>(0.5483) | -2.3041<br>(0.0469) | 0.2203<br>(0.7091) | 0.3783<br>(0.5228) | 0.3643<br>(0.5862) | 1.2455<br>(<.0001) | -2.6583<br>(<.0001) | -0.2411<br>(0.5005) | 0.4838<br>(0.6197) | 0.2925<br>(0.5878) | 0.1028<br>(0.8857) | -0.6351<br>(0.517) | -0.7659<br>(0.0024) | -0.9941<br>(0.1403) | -0.2381<br>(0.3536) | -0.0593<br>(0.967) | -0.6414<br>(0.1405) | 0.1872<br>(0.6998) | -0.3697<br>(0.4939) | -0.1159<br>(0.5275) | 0.4728<br>(0.0577) |
| I am the parent or guardian of any child under the age of 18 living in my household | 0.6043<br>(0.0051) | -1.125<br>(0.3298) | 0.6975<br>(0.1487) | 0.8602<br>(0.1869) | 0.5432<br>(0.2416) | 0.7112<br>(<.0001) | 0.9105<br>(0.3597) | 0.2888<br>(0.3602) | -0.7249<br>(0.3815) | -0.4586<br>(0.402) | 0.672<br>(0.2518) | 1.0432<br>(0.0468) | 0.6682<br>(0.1701) | 0.291<br>(0.8144) | 1.0018<br>(0.003) | 0.8971<br>(0.2538) | 2.0707<br>(0.0002) | 0.9322<br>(0.2497) | 0.6254<br>(0.5145) | 0.6731<br>(<.0001) | -1.2694<br>(<.0001) |
| Are you responsible for taking care of any elderly people? | 1.1777<br>(<.0001) | 3.0077<br>(0.0058) | 0.3723<br>(0.3735) | 1.81<br>(0.0042) | 0.8099<br>(0.0944) | 1.248<br>(<.0001) | 1.3666<br>(0.0043) | 1.4318<br>(0.0004) | 0.2624<br>(0.8045) | 1.7746<br>(0.0002) | 1.0298<br>(0.2329) | 0.406<br>(0.4568) | 3.345<br>(<.0001) | 2.299<br>(0.0139) | 1.5875<br>(<.0001) | 1.6956<br>(0.1928) | 1.9388<br>(0.0103) | 1.5838<br>(0.0065) | 0.4903<br>(0.2175) | 3.1281<br>(<.0001) | 1.2117<br>(<.0001) |
| Someone in my household works in a health care delivery setting, such as a doctor's office, clinic, hospital, nursing home, | 0.8155<br>(0.0157) | 0.2046<br>(0.8559) | 1.0849<br>(0.0811) | 0.4981<br>(0.4489) | 0.317<br>(0.5949) | 1.1433<br>(<.0001) | 4.3687<br>(<.0001) | 0.6583<br>(0.1007) | 1.8486<br>(0.2184) | 1.1275<br>(0.0151) | -0.1991<br>(0.8404) | 1.4263<br>(0.0274) | -0.8738<br>(0.0906) | -2.64<br>(0.0678) | 0.7337<br>(0.0431) | 1.2612<br>(0.3801) | 0.1824<br>(0.8296) | 1.5229<br>(0.0546) | 0.2132<br>(0.6567) | 1.5654<br>(0.0401) | -2.2322<br>(<.0001) |
|  | “Supplies” Impact Subscore<br>GEE parameter estimate<br>(p-value) |  |  |  |  |  |  | “Cancellations” Impact Subscore<br>GEE parameter estimate<br>(p-value) |  |  |  |  |  |  | “Livelihood” Impact Subscore<br>GEE parameter estimate<br>(p-value) |  |  |  |  |  |  |
| Variable | Total | Africa | Asia | Europe | Latin America and the Caribbean | Northern America | Oceania | Total | Africa | Asia | Europe | Latin America and the Caribbean | Northern America | Oceania | Total | Africa | Asia | Europe | Latin America and the Caribbean | Northern America | Oceania |
| or dentist's office |  |  |  |  |  |  |  |  |  |  |  |  |  |  |  |  |  |  |  |  |  |
| Rate of population increase (2015) | 0.3456<br>(0.4245) | 0.8279<br>(0.4484) | -1.1206<br>(0.0753) | -0.5685<br>(0.4168) | -0.5076<br>(0.6681) | 0 (.) | 0 (.) | -0.2689<br>(0.3882) | 3.5151<br>(<.0001) | -1.3664<br>(0.0111) | -0.8531<br>(0.4642) | 0.5194<br>(0.2789) | 0 (.) | 0 (.) | -0.1274<br>(0.6746) | -0.8444<br>(0.5871) | -0.2783<br>(0.6769) | -2.7381<br>(0.0262) | 0.8792<br>(0.0211) | 0 (.) | 0 (.) |
| Unintentional homicide rate | 0.0215<br>(0.2306) | -0.0283<br>(0.7791) | 0.8357<br>(0.0002) | -0.4746<br>(0.3472) | -0.0018<br>(0.9259) | 0 (.) | 0 (.) | 0.0041<br>(0.8191) | 0.04<br>(0.6095) | 0.2254<br>(0.1422) | -0.4236<br>(0.5226) | 0.0327<br>(0.0962) | 0 (.) | 0 (.) | 0.0499<br>(0.0216) | 0.2003<br>(0.0384) | 0.2254<br>(0.1938) | 0.8094<br>(0.0939) | 0.0119<br>(0.518) | 0 (.) | 0 (.) |
| Rate of change in Disability Adjusted Life Years (1990-2017) | 2.3498<br>(0.3132) | 0.4838<br>(0.934) | -4.4183<br>(0.4781) | -5.3275<br>(0.2354) | 2.3822<br>(0.6359) | 0 (.) | 0 (.) | -2.7119<br>(0.1112) | 1.1041<br>(0.7848) | -<br>14.6014<br>(0.0005) | -<br>11.4349<br>(0.1715) | 6.4974<br>(0.116) | 0 (.) | 0 (.) | -0.1212<br>(0.9498) | -4.4877<br>(0.4556) | -2.0172<br>(0.609) | -1.2222<br>(0.7994) | -8.2724<br>(0.0012) | 0 (.) | 0 (.) |
| Rate of change in overall death rate (1990-2017) | -7.6887<br>(0.0389) | -5.655<br>(0.744) | 4.8177<br>(0.1776) | 0.6914<br>(0.9458) | 10.0102<br>(0.3795) | 0 (.) | 0 (.) | 2.3449<br>(0.4148) | -<br>17.1357<br>(0.1761) | 14.4097<br>(0.0022) | 7.5162<br>(0.6964) | -1.1512<br>(0.8837) | 0 (.) | 0 (.) | 0.0798<br>(0.9823) | 4.0803<br>(0.7421) | 6.1975<br>(0.0801) | -<br>17.3171<br>(0.2071) | -4.5291<br>(0.4306) | 0 (.) | 0 (.) |
| Change in Death rate per 100,000, July v. April | -1.5724<br>(0.0701) | 0.7792<br>(0.6838) | 1.7847<br>(0.2875) | 3.0239<br>(0.2176) | -3.402<br>(0.0102) | 0 (.) | -5.1529<br>(0.3719) | -1.2802<br>(0.0409) | -1.8502<br>(0.2002) | 1.1748<br>(0.4384) | 0.1723<br>(0.9708) | 0.5027<br>(0.6415) | 0 (.) | 27.6149<br>(<.0001) | 1.3017<br>(0.0814) | 1.106<br>(0.6) | 1.9648<br>(0.1285) | 0.9026<br>(0.668) | -1.0673<br>(0.1611) | 0 (.) | 12.4094<br>(0.0779) |
|  | “Supplies” Impact Subscore<br>GEE parameter estimate<br>(p-value) |  |  |  |  |  |  | “Cancellations” Impact Subscore<br>GEE parameter estimate<br>(p-value) |  |  |  |  |  |  | “Livelihood” Impact Subscore<br>GEE parameter estimate<br>(p-value) |  |  |  |  |  |  |
| Variable | Total | Africa | Asia | Europe | Latin America and the Caribbean | Northern America | Oceania | Total | Africa | Asia | Europe | Latin America and the Caribbean | Northern America | Oceania | Total | Africa | Asia | Europe | Latin America and the Caribbean | Northern America | Oceania |
| Change inInfection rate per 100,000, July v. April | 0.0025<br>(0.6998) | 0.0208<br>(0.2375) | -0.0833<br>(0.0003) | 0.6055<br>(0.0157) | 0.0017<br>(0.8543) | 0 (.) | -0.6898<br>(0.8473) | -0.0105<br>(0.0028) | 0.0021<br>(0.8534) | -0.0398<br>(0.0459) | 0.8746<br>(0.0144) | 0.0076<br>(0.2993) | 0 (.) | 15.2005<br>(<.0001) | 0.0117<br>(0.0851) | 0.0249<br>(0.0835) | -0.0378<br>(0.0007) | 0.125<br>(0.6978) | -0.0128<br>(0.0021) | 0 (.) | 6.6212<br>(0.048) |
| Overall death rate from respiratory infections | 0.0206<br>(0.0603) | -0.0052<br>(0.833) | -0.0228<br>(0.3259) | 0.1213<br>(<.0001) | 0.0243<br>(0.5348) | 0 (.) | 0 (.) | 0.0092<br>(0.2548) | -0.0475<br>(0.0189) | 0.0127<br>(0.3007) | 0.0864<br>(0.1337) | 0.0451<br>(0.1228) | 0 (.) | 0 (.) | -0.0016<br>(0.8685) | 0.0094<br>(0.582) | -0.0261<br>(0.0537) | -0.0841<br>(0.0063) | 0.007<br>(0.739) | 0 (.) | 0 (.) |
| Air Pollution Concentration (ambient particulate pollution (PM2.5) ) | 0.0466<br>(0.1082) | 0.264<br>(0.0114) | 0.0786<br>(0.1611) | 0.1496<br>(0.1571) | 0.2394<br>(0.0956) | -0.0307<br>(0.2516) | 4.7871<br>(0.2215) | 0.0368<br>(0.0204) | 0.1498<br>(0.0116) | -0.0178<br>(0.6593) | -0.0007<br>(0.9969) | -0.07<br>(0.3946) | 0.1544<br>(0.0019) | -<br>15.0953<br>(<.0001) | 0.0125<br>(0.5511) | 0.1126<br>(0.2959) | 0.0475<br>(0.1629) | -0.0055<br>(0.9613) | -0.0366<br>(0.5772) | 0.3805<br>(0.0432) | -6.8828<br>(0.0291) |

#### Individual Level

The Attitudes toward COVID-19 Score (an index of COVID-related “worry”) was positively associated with the Impact Score in all regions (as the Attitudes Score increased (e.g., more COVID-related worry), the Impact Score and its component scores increased.) Similarly, the COVID-19-related Knowledge Score was negatively associated with the Impact Score, though not consistently in all regions (in particular in Africa and in Latin America and the Caribbean). Further, with the exception of the “Personal Action” Impact sub-score (which reflects mostly preventive actions such as procuring and wearing masks, and staying home), as COVID-related Knowledge declined, the personal Impact Score increased. As COVID-related knowledge increased, the Personal Action sub-score increased overall, in Europe, Northern America, and Oceania.

The Multidimensional Health Locus of Control’s subcomponents differed by region and Impact sub-score. “Powerful Others” locus of control was positively associated with the total Impact Score at the global level and in Europe, Northern America, and Oceania: as the participant’s belief strengthened that their health is under the control of powerful others, their Impact Score also increased. The “Powerful Others” locus of control was positively associated with the “Cancellation” Impact sub-score in the total sample and in all regions: as the belief that one’s health is under the control of others strengthened, the negative impact of cancellations and similarly postponements increased. The other components of the Multidimensional Health Locus of Control were inconsistently associated across regions and Impact sub-scores. With the exception of the “Supplies” Impact sub-score, the “Internal” locus of control subcomponent was not consistently associated with Impact, though as the sense the one’s health was under one’s own control declined, the negative impact associated with procuring supplies increased overall globally, and in Northern America and Oceania in particular.

While having enough information to protect against COVID-19 was not associated overall with Impact or any of the four sub-scores at the global level, it was associated with Impact in Latin America, Northern America, and (especially) in Oceania. Further, having enough information was inconsistently associated with Supplies Impact: in Latin America, having more information was associated with higher Supplies Impact, but in Northern America and Oceania, it was associated with lower Supplies Impact.

Participants who reported believing they had COVID-19 infection were significantly and strongly associated with overall Impact and with each of its sub-scores, in particularly with the Livelihood and Supplies sub-scores, and especially within Latin America and the Caribbean, Northern America, and Asia where having reported COVID-19 was positively associated with Impact. Similarly, respondents who reported that they had a chronic health condition consistently experienced greater overall Impact and at all sub-scores with the exception of Livelihood, where having a chronic condition was associated with Impact only in Northern America.

Working in a health care delivery setting was not associated with overall Impact in any region nor globally, but was consistently and inversely associated with Personal Action Impact in all regions except Europe: as working in a health care setting increased, Personal Action Impact decreased. This type of work setting was significantly associated with Livelihood Impact in North America and, especially, in Asia, though not in the other regions.

Access to material assets was associated with overall Impact only in Northern America, though (with the exception of Europe) was associated with Personal Action in all regions: those with ownership of a car or home reported higher Personal Action impact, and in Northern America reported higher Supplies Impact. Globally, ownership of a home was inversely associated with Livelihood Impact, with those not owning a home more likely to experience higher impact on their livelihood.

#### Household/ Family Level

With the consistent exception of Europe and Africa, knowing someone who has -- or who has had -- COVID-19 was associated with greater Impact overall and across subcomponents except the “Supplies” Impact sub-score. The association between knowing someone with COVID-19 and Impact is particularly strong overall in Northern America and Oceania regarding Personal Action and “Cancellation”. That said, knowing someone with COVID-19 was not associated with the Supplies Impact subscore, and for the Overall Impact Score and Personal Action sub-score, knowing someone with COVID-19 was inversely associated in Asia (as knowing someone with COVID-19 increased, Impact and Personal Action decreased). Additionally, knowing someone who had died from COVID-19 was similarly associated with Impact and its sub-scores, particularly in Northern America. Notably, knowing someone who had died from COVID-19 in Oceania was strongly associated with increased “Livelihood Impact,” and knowing someone who had died from COVID-19 was strongly associated with “Supplies” Impact in Africa.

Caring for elderly and children was associated with higher Impact Scores globally, and across most regions and Impact Sub-scores, with the exception of Latin America and the Caribbean where caring for elderly was not associated with any of the Impact Scores and where caring for children was associated only with the “Cancellation” Impact subscore. Caring for the elderly was particularly associated positively with the Personal Action and Livelihood Impact sub-scores in all regions except Latin America and the Caribbean and Africa: as caring for the elderly increased, the impact on Personal Action and Livelihood increased. Caring for children was inconsistently associated with Personal Action Impact in Africa and in Northern America: in Africa, those not caring for children were more likely to experience Personal Action Impact while in Northern America, those caring for children were more likely to experience Personal Action Impact.

Having another person in the household with a chronic illness was not associated with overall Impact globally or in any region specifically. In Africa and in Oceania, having another person in the household with a chronic condition was associated with lower Supplies Impact, while in Northern America it was associated with greater Supplies Impact.

Having another person in the household working in a health care delivery setting was inconsistently associated with Impact: in Latin America and the Caribbean, for instance, having a household member working in health care was associated with higher overall Impact, while in Northern America and in Oceania, it was associated with lower overall Impact. Also, in Northern America, those with household members working in health care had higher Livelihood Impact while in Oceania, those with household members working in health care had lower Livelihood Impact.

#### Country-level Variables

Country-level variables were associated with Impact and its sub-scores inconsistently across regions. Two variables were associated with overall Impact at the global level: “ambient particulate pollution and the unintentional homicide rate”, both of which were also positively associated with Personal Action, with Pollution also positively associated with Cancellation Impact and with Homicide also associated with Livelihood Impact. Pollution was consistently inversely associated with Impact in Oceania (less pollution and more impact).

Several demographic and health variables (rate of population increase, rate of change in disability-adjusted life years, and rate of change in death rate) at the country level were consistently related to Impact predominantly in Asia and secondarily in Africa. In Asia, Overall Impact, Personal Action Impact, and Cancellation Impact all increased with lower country-rates of population growth and of change in Disability Adjusted Life Years (DALYs). In Africa, these same areas of Impact were all positively associated with larger rates of population change. In addition, larger changes in the overall death rates globally were associated with higher Personal Action Impact.

COVID-19 related metrics were most consistently associated with Impact in Oceania, the only region where higher COVID-19-related death and infection rates over time were associated with higher overall Impact and Cancellation Impact. Further, Livelihood Impact was most strongly associated with higher COVID-19 infection rates over time only in Oceania, though in Asia and in Latin America and the Caribbean Livelihood Impact was inversely associated with higher infection rates over time (as the rate of infection increased from April to July, the Livelihood Impact decreased).

### Effect Size

The proportions of overall Impact Score effect size attributable to levels and domains of the medical ecological model are presented in Figures 3 and 4.

**Figure 3.**
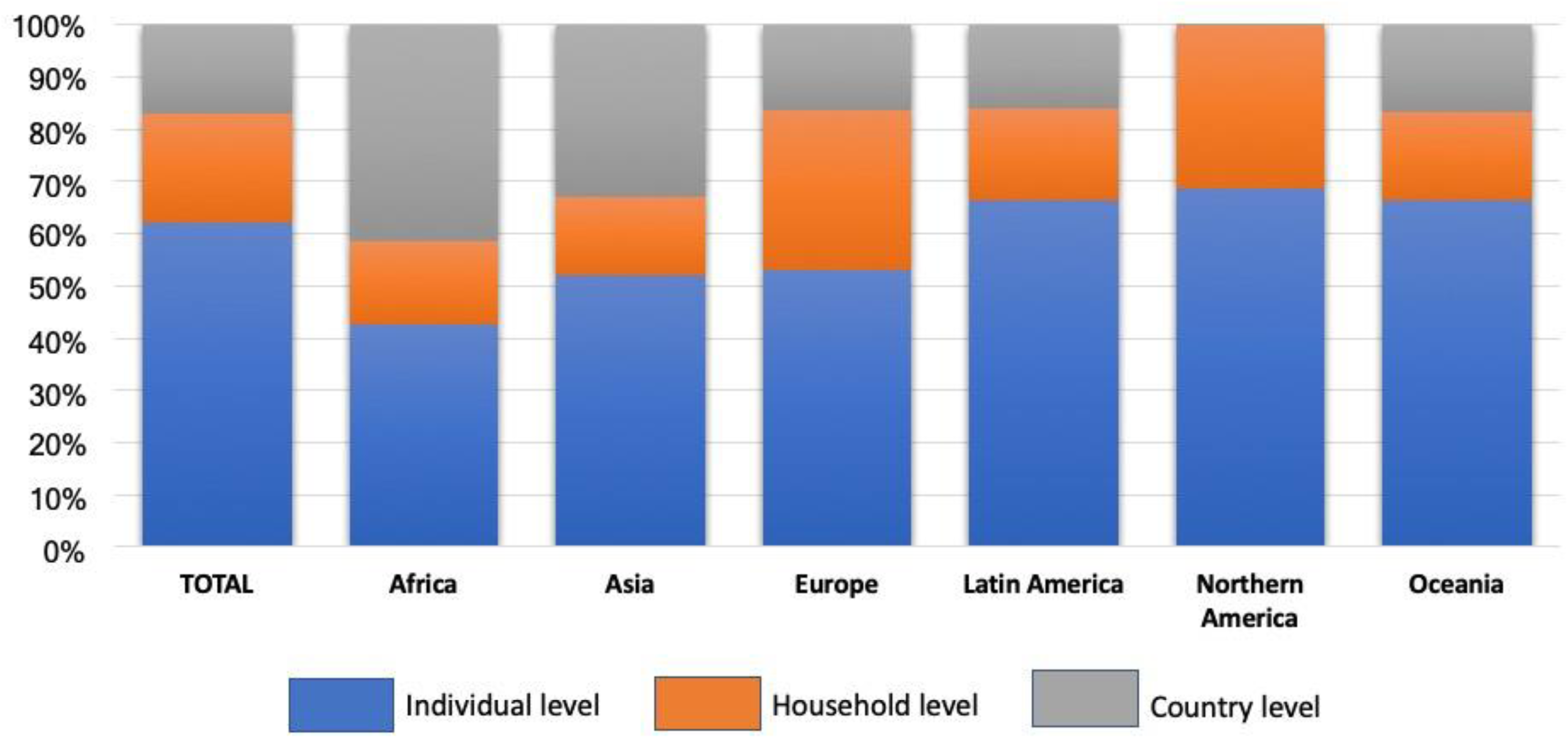
Variation in Total Impact Score, Attributable to Variables at Different Levels of the Medical Ecological Model.

**Figure 4.**
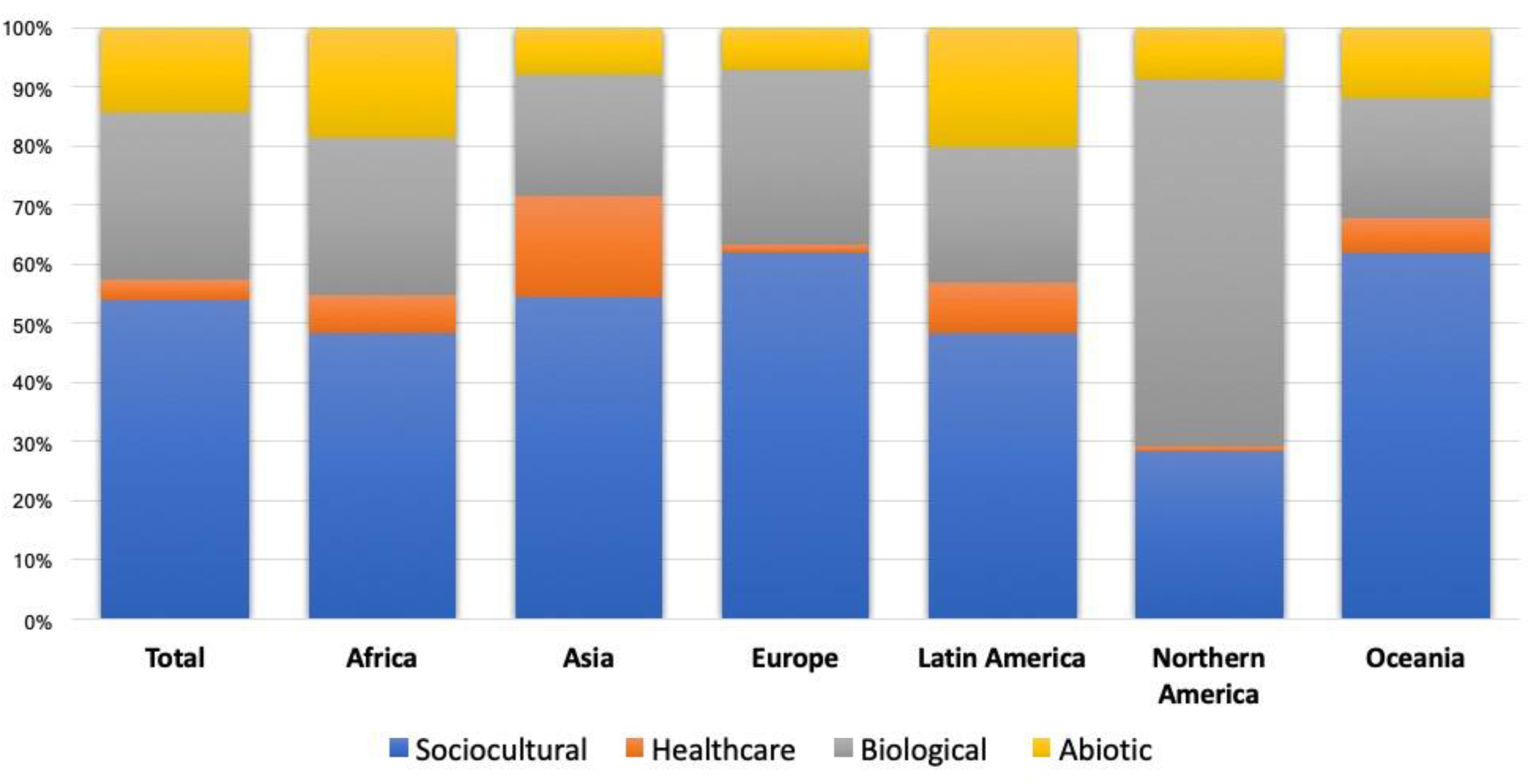
Variation in Total Impact Score, Attributable to Variables in Different Domains of the Medical Ecological Model.

#### Effect size attributable to level of the medical ecological model

Overall, 62% of the adjusted variation in Impact Score effect size (Figure 3) was attributable to Individual level factors such as COVID-19 related knowledge, worry, and locus of control. Additionally, variation in the Impact Score effect size attributable to household-level variables (21%) and to country-level variables, 17%. Northern America, Latin America, and Oceania had the highest proportion (69%, 66%, and 66%, respectively) of effect size attributable to individual-level variables, while Africa had the lowest (43%). Africa and Asia had the highest proportion of Impact Score variation in effect size attributable to country-level variables (41% and 33%, respectively).

#### Effect size attributable to domain

Substantially more variation was noted by region in proportions of Impact Score effect size attributable to different domains (Figure 4). Sociocultural variables accounted for 54% of Impact Score effect size variation, with 28% of variation explained by biological, 14% by abiotic/ environmental, and 4% explained by healthcare variables. The pattern of Impact Score effect size variation in Africa and Latin America similar, with almost half of the variation attributable to sociocultural variables, about 1/4 to biological variables, about 1/5 to abiotic variables, and low proportions of healthcare variables (<10%). Northern America’s patterning is unique in that much of the effect size variation was attributable to biological variables (>60%), while Asia had the highest proportion attributable to healthcare variables (17%). Oceania and Europe had the highest proportions of effect size variation attributable to sociocultural variables (>60%).

### Qualitative Multilevel Modeling

Table 8 presents a summary of the correlation between qualitative code clusters, Impact Score, and its subscores. Each score is uniquely correlated with qualitative experience, providing insight into dynamics and mechanisms of non-medical impact. (Note: Previously mentioned, all scores are Standardized T-scores, with an average of 50 and standard deviation of 10. Higher scores indicate greater intensity of the score/ subscore).

**Table 8:**
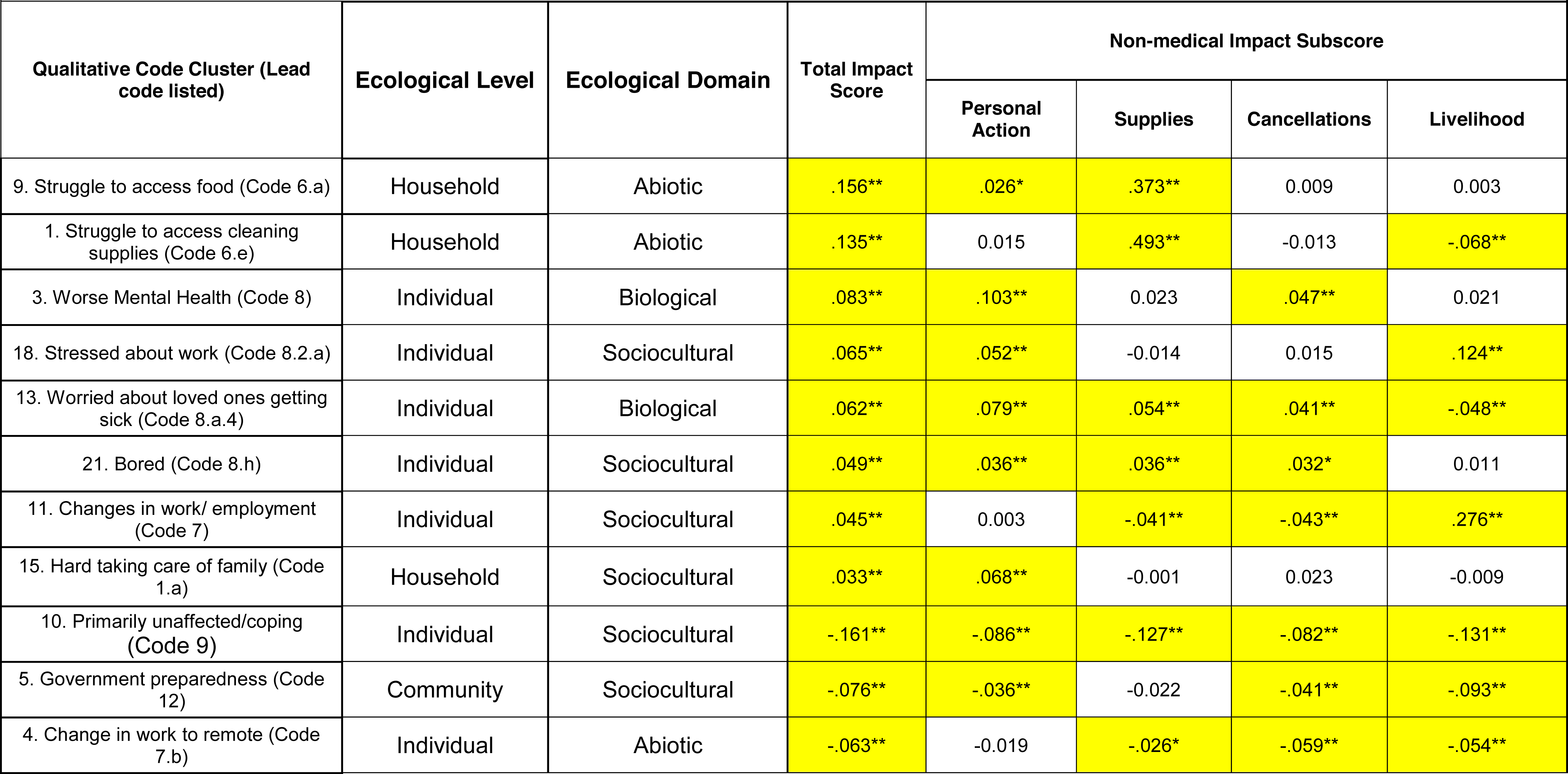
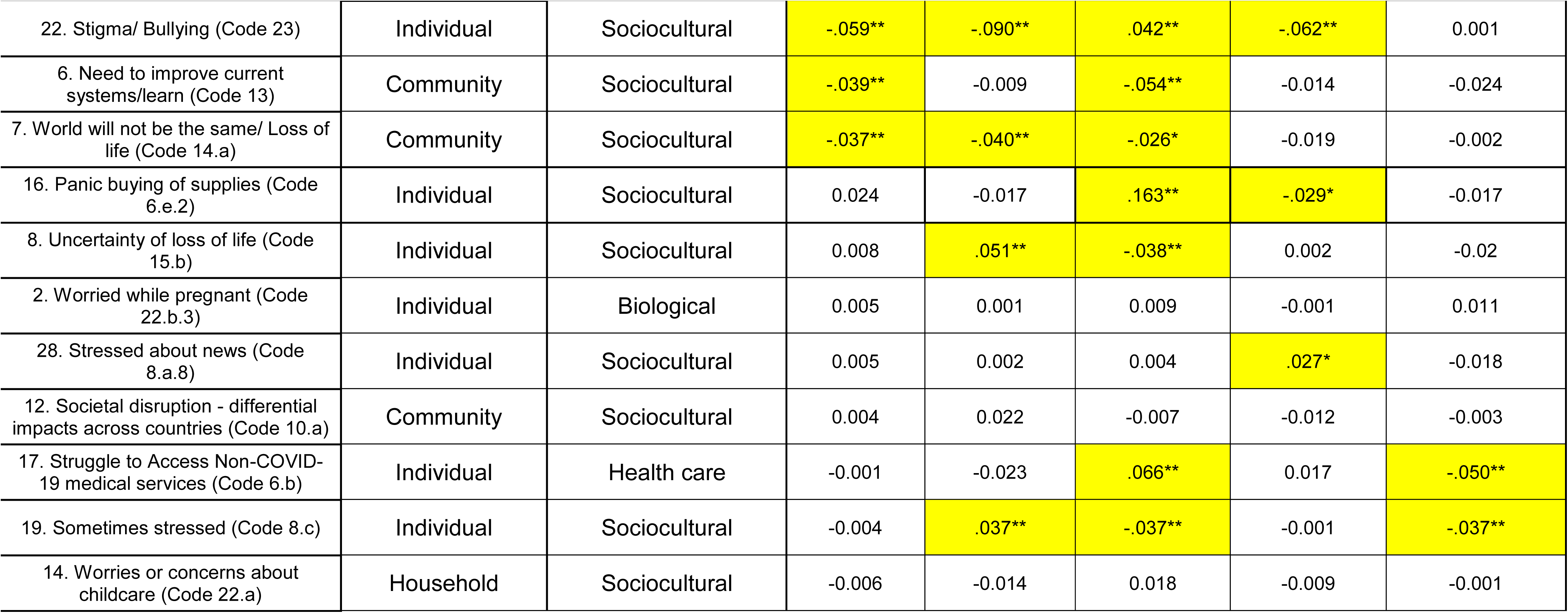
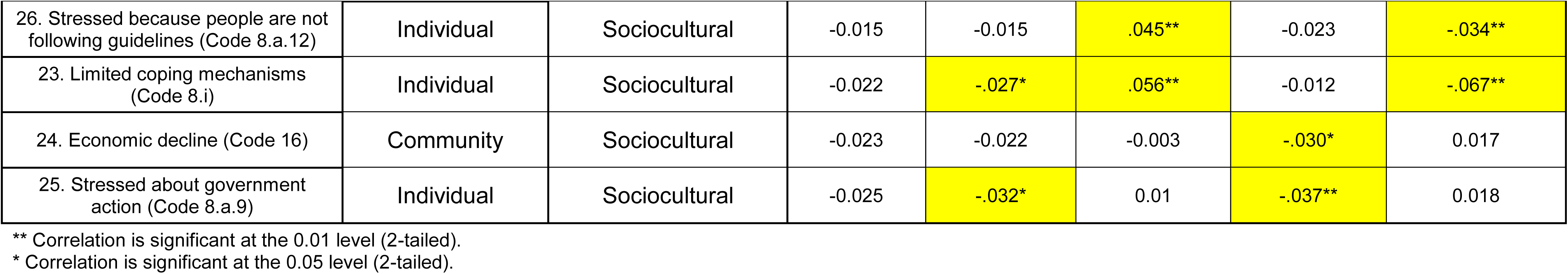
Spearman’s Correlation Coefficient between Qualitative Code Clusters Mapped to Ecological Model and COVID-19 Non-Medical Impact Score and Subscores (sorted by correlation coefficient with Total Impact Score)

| Qualitative Code Cluster (Lead code listed) | Ecological Level | Ecological Domain | Total Impact Score | Non-medical Impact Subscore |  |  |  |
| --- | --- | --- | --- | --- | --- | --- | --- |
|  |  |  |  | Personal Action | Supplies | Cancellations | Livelihood |
| 9. Struggle to access food (Code 6.a) | Household | Abiotic | .156** | .026* | .373** | 0.009 | 0.003 |
| 1. Struggle to access cleaning supplies (Code 6.e) | Household | Abiotic | .135** | 0.015 | .493** | -0.013 | -.068** |
| 3. Worse Mental Health (Code 8) | Individual | Biological | .083** | .103** | 0.023 | .047** | 0.021 |
| 18. Stressed about work (Code 8.2.a) | Individual | Sociocultural | .065** | .052** | -0.014 | 0.015 | .124** |
| 13. Worried about loved ones getting sick (Code 8.a.4) | Individual | Biological | .062** | .079** | .054** | .041** | -.048** |
| 21. Bored (Code 8.h) | Individual | Sociocultural | .049** | .036** | .036** | .032* | 0.011 |
| 11. Changes in work/ employment (Code 7) | Individual | Sociocultural | .045** | 0.003 | -.041** | -.043** | .276** |
| 15. Hard taking care of family (Code 1.a) | Household | Sociocultural | .033** | .068** | -0.001 | 0.023 | -0.009 |
| 10. Primarily unaffected/coping (Code 9) | Individual | Sociocultural | -.161** | -.086** | -.127** | -.082** | -.131** |
| 5. Government preparedness (Code 12) | Community | Sociocultural | -.076** | -.036** | -0.022 | -.041** | -.093** |
| 4. Change in work to remote (Code 7.b) | Individual | Abiotic | -.063** | -0.019 | -.026* | -.059** | -.054** |
|  |  |  |  | Personal Action | Supplies | Cancellations | Livelihood |
| 22. Stigma/ Bullying (Code 23) | Individual | Sociocultural | -.059** | -.090** | .042** | -.062** | 0.001 |
| 6. Need to improve current systems/learn (Code 13) | Community | Sociocultural | -.039** | -0.009 | -.054** | -0.014 | -0.024 |
| 7. World will not be the same/ Loss of life (Code 14.a) | Community | Sociocultural | -.037** | -.040** | -.026* | -0.019 | -0.002 |
| 16. Panic buying of supplies (Code 6.e.2) | Individual | Sociocultural | 0.024 | -0.017 | .163** | -.029* | -0.017 |
| 8. Uncertainty of loss of life (Code 15.b) | Individual | Sociocultural | 0.008 | .051** | -.038** | 0.002 | -0.02 |
| 2. Worried while pregnant (Code 22.b.3) | Individual | Biological | 0.005 | 0.001 | 0.009 | -0.001 | 0.011 |
| 28. Stressed about news (Code 8.a.8) | Individual | Sociocultural | 0.005 | 0.002 | 0.004 | .027* | -0.018 |
| 12. Societal disruption - differential impacts across countries (Code 10.a) | Community | Sociocultural | 0.004 | 0.022 | -0.007 | -0.012 | -0.003 |
| 17. Struggle to Access Non-COVID-19 medical services (Code 6.b) | Individual | Health care | -0.001 | -0.023 | .066** | 0.017 | -.050** |
| 19. Sometimes stressed (Code 8.c) | Individual | Sociocultural | -0.004 | .037** | -.037** | -0.001 | -.037** |
| 14. Worries or concerns about childcare (Code 22.a) | Household | Sociocultural | -0.006 | -0.014 | 0.018 | -0.009 | -0.001 |

|  |  |  |  |  |  |  |  |
| --- | --- | --- | --- | --- | --- | --- | --- |
| Qualitative Code Cluster (Lead code listed) | Ecological Level | Ecological Domain | Total Impact Score | Non-medical Impact Subscore |  |  |  |
|  |  |  |  | Personal Action | Supplies | Cancellations | Livelihood |
| 26. Stressed because people are not following guidelines (Code 8.a.12) | Individual | Sociocultural | -0.015 | -0.015 | .045** | -0.023 | -.034** |
| 23. Limited coping mechanisms (Code 8.i) | Individual | Sociocultural | -0.022 | -.027* | .056** | -0.012 | -.067** |
| 24. Economic decline (Code 16) | Community | Sociocultural | -0.023 | -0.022 | -0.003 | -.030* | 0.017 |
| 25. Stressed about government action (Code 8.a.9) | Individual | Sociocultural | -0.025 | -.032* | 0.01 | -.037** | 0.018 |

\*\* Correlation is significant at the 0.01 level (2-tailed).
\* Correlation is significant at the 0.05 level (2-tailed).

#### Qualitative Code correlation with Personal Action Impact SubScore

The “Personal Action” Subscore of COVID-19-related non-medical impact included volitional individual acts such as wearing a mask, obtaining extra prescriptions, or staying home. The strongest correlation of the Personal Action subscore was with *worse mental health (code 8);* in fact, Personal Action was the subscore with the highest correlation with this code among the subscores. Respondents with higher Personal Action subscores frequently mentioned a range of personal actions undertaken at the individual level and their entanglement with worsening mental health:

> *Tutto ciò che è stato consigliato….siamo in 5 persone in famiglia, ma io sono l’unica che esce ogni settimana…e tornando dalla spesa, pulisco tutte le cose acquistate, mi lavo le mani, sistemo tutto, pulisco le suole delle scarpe, le mascherine e poi mi faccio la doccia. sono molto più nervosa perchè comunque sono disoccupata e non posso uscire per cercare lavoro…. (uno dei tanti problemi che stanno vivendo in molti). Al momento vedo tutto negativo. Ero uscita da una situazione di depressione, ma questa malattia ha contribuito a farla riemergere…* (All that has been recommended …. we are 5 people in the family, but I am the only one who goes out every week … and returning from the shopping, I clean all the things bought, I wash my hands, I fix everything, I clean the soles of my shoes, the masks and then I take a shower. I am much more nervous because in any case I am unemployed and I cannot go out to look for a job …. (one of the many problems that many are experiencing). At the moment, I see everything negative. I had come out of a situation of depression, but this illness helped to make it re-emerge.) (Southern Europe woman in her 50s; Personal Action sub-score: 55.1; #2-1633)
>
> *Constant anxiety when going to buy groceries. Fear of touching things in public places…. Staying home and not being able to go out to work. Missed opportunities. Financial burden… Business has been slow and in financial hardship…. Non-availability of goods….* (South Asian man in his 30s; Personal Action sub-score 55.5; #1-880)

Additionally, respondents with high Personal Action sub-scores often attributed their mental health experience as nested within larger social factors at the community and country levels:

> *Tengo depresion, ya que el futuro es incierto y tenemos un gobierno dictatorial opresivo e inutil.* (I am depressed, as the future is uncertain and we have an oppressive and useless dictatorial government.) (Latin America woman in her 60s; Personal Action sub-score: 62.9; #2-2501)

Respondents with lower Personal Action sub-scores occasionally associated that their adherence to prevention recommendations and limiting news related to preventing worsening mental health:

> *We avoid large gatherings, wash hands, shower regularly, keep our house clean, and disinfect ourselves after going somewhere…I don’t spend too much time on facebook because I try to avoid reading sad information about corona virus. I feel sad when I find out the number of people dying because of the virus….It is a pandemic disease and there is no cure yet. Everyone should follow or obey the rules in order to be safe and protect the spread of the virus. Coronavirus is serious and everyone must take it seriously!* (Woman in her 30s, living in Pacific Islands; Personal Action sub-score 40.5; #2-1106)

Indeed, the *Primarily unaffected/coping (Code 9)* code cluster – which includes adherence to prevention recommendations – is strongly (and inversely) correlated with Personal Action sub-score, shown by this respondent (with one of the lowest Personal Action sub-scores):

> *We only go outside the recommended amount, only one person does a shop, don’t gather or visit groups of people, stay 2m away…I am not too stressed regarding the virus due to the precautions we have taken, we also live in an incredibly rural area and so there are few people around anyway. Have some [worries] regarding the economy but will cross that bridge when we come to it… think it is incredibly serious but many people aren’t approaching it as if it is*. *Videos every week of crowds of people, including emergency services clapping for the NHS infuriates me! Lovely to clap but in huge groups with the elderly and children??? just asking for death and sorrow… (*Northern European woman in her early 20s; Personal Action sub-score 33.1; #1-2411)

One additional qualitative code cluster of note is *Hard taking care of family (Code 1.a)*, which is correlated with the Personal Action subscore, and in fact the only subscore that is correlated with this cluster (which also includes codes for staying home and changes in education). Respondents with this code frequently mention the increased demands of caring for children at home during lockdown:

> *Je suis plus vigilante aux problèmes de santé dans ma famille…Nous sommes assignés à résidence depuis 41 jours alors que de grands espaces naturels tout proches pourraient nous permettre de respirer. Les enfants sont à la maison non-stop et tournent en rond. Nous avons malgré tout vu les grands parents une fois en faisant attention.* (I am more vigilant to health problems in my family … We have been under house arrest for 41 days while large natural spaces nearby could allow us to breathe. The children are at home nonstop and going in circles. We did see the grandparents once though, being careful. (Western Europe respondent; Personal Action sub-score 62.5; #1-3356)

Respondents effectively being able to respond to family needs at home with lower Personal Action often mentioned security in resources:

> *Total lockdown in Spain. Only going out to buy food and medicines minimum number of times possible. Wearing mask when out. Hand washing…There is a fairly positive interaction between my wife and I and our 2 early 20s sons who live in same house. We are not short of money which is an important issue…Am pretty stress free and almost enjoying not working*. *Have started writing a kind of memoir instead! We can ride this out financially although we’ll obviously take something of a hit - but hopefully not enough to make life complicated.* (Man in his 60s living in Southern Europe; Personal Action Score 55.5; #1-2087)

Finally, while some respondents indicated they did not perceive COVID-19 as impacting their lives, their Personal Action sub-score suggests they, in fact, have been impacted:

> *Cambio radical de estilo de vida …El coronavirus fue subestimado y ahora causa estragos que pudieran haber disminuidos con una buena acción del gobierno…Mi vida no ha cambiado mucho, mi forma de ingreso es de manera remota, asi que no afecta mis ingresos…Un poco deprimido y estresado, pero manteniendome firme… (*Radical lifestyle change … The coronavirus was underestimated and now it causes havoc that could be diminished with a good government action … My life has not changed much, my form of income is remotely, so it does not affect my income … A little depressed and stressed, but holding my ground.) (Man in his 20s living in South America; Personal Action Subscore 62.9; #1-4087)

#### Qualitative Code correlation with Supplies Impact SubScore

The Supplies Impact Subscore reflects the inability of respondents to obtain groceries, cleaning supplies, and medications. Shown in Table 7, the Supplies Subscore had the highest correlations with Qualitative Code Clusters among all four subscores and the Total Impact Score. These two areas of qualitative clustering (Struggle to access cleaning supplies (Code 6.e) and Struggle to access food (Code 6.a) incorporated other qualitative codes such as shortages, income insufficiency, and price gouging. The inability to obtain supplies – especially those necessary to help families prevent COVID-19 – created additional burdens and frustrations on respondents, reflected in the strongest correlation of qualitative experience with the Supplies Subscore:

> *I feel extremely depressed and like this will never end. Everything is always sold out. This is an absolutely disaster and I’m finding it harder and harder to stay positive and have any hope that things will get better any time soon.* (Woman in her early 20s living in Northern America; Supply Subscore 64.1; #1-2661)
>
> *J’ai peur pour mes amis et leurs familles qui ont été diagnostiqué positif au Covid-19. J’essaye de les joindre quotidiennement par tout le moyens mais sans espoir, Depuis 3 jours je n’ose même plus quitter mon lit et attendre un appel ou un message…Que les gérants de Supermarchés, Épicier, Marchés reviennent a être malade du Covid. J’ai peur de sortir, Peur de m’approcher des gens, Peur de ce qui pourrait arriver a mes amis et a ma famille. Les marchés sont surpeuplé et les stands beaucoup trop proche j’ai peur pour ma santé. A chaque fois que j’essaie le pharmacien me demande de revenir un autre jour.. je suppose qu’ils n’ont pas l’intention d’en vendre (gel) a des jeunes adultes.* (I fear for my friends and their families who have tested positive for Covid-19. I try to reach them daily by all means but without hope, for 3 days I don’t even dare to leave my bed and wait for a call or a message … That the managers of Supermarkets, Grocers, Markets come back to be sick of the Covid. I’m scared to go out, scared of approaching people, scared of what might happen to my friends and family. The markets are overcrowded and the stalls far too close I fear for my health. Every time I try, the pharmacist asks me to come back another day.. I guess they don’t intend to sell (gel) to young adults. (Man in his early 20s living in North Africa; Supplies Subscore 64.1; #2-1502)

Also, strongly correlated with Supplies Sub-score – perhaps with more urgency – are qualitative codes relating to procuring food, nearly always confounded by lack of income and availability, conveyed through these respondents with a very high Supplies Subscores:

> *Desespera la situación económica eso es mas preocupante que el coronavirus porque las deudas siguen creciendo y no hay trabajo…No hay dinero para hacer la despensa…No tengo dinero. Odio esta situación. Ya quiero ir q trabajar necesito dinero para darle de comer a la familia.* (The economic situation is desperate, that is more worrisome than the coronavirus because the debts continue to grow and there is no work … There is no money to stock the pantry … I have no money. I hate this situation. I already want to go to work - I need money to feed the family.) (Woman in her early 30s living in Central America; Supplies Subscore 80; #2-1821)
>
> *“Mon pays a un systeme de santé precaire,manque de materiels de réanimation, personnel non formé, capacités de lits limité…Dans mon pays on vit du jour au lendemain ce qu’on cherches c’est ce qu’on mange…on prefere sortir contracter corona que de mourrir de faim en restant à la maison!”* (My country has a precarious health system, lack of resuscitation equipment, untrained staff, limited bed capacity … In my country we live overnight - what we seek is what we eat…one would rather go out and contract corona than starve to death while staying at home!) (Man in his 30s living in Sub-Saharan Africa; Supplies Subscore 75.0; #2-1390)

Similarly, the complicity of income and access to resources on procuring food is clear throughout all regions:

> *In media anche nei paesi più piccoli si può sempre trovare quasi tutto. farmacie, generi alimentari, oggettistica. alcuni settori purtroppo stanno man mano scomparendo (contadini e allevatori che vendano direttamente il proprio prodotto)… Nizialmente Pasta, lievito, prodotti senza lattosio, farina. Ora rimane più difficile trovare il lievito, ma con un po di fortuna si trova. Frutta, Verdura e Carne sono scadenti e a caro prezzo, ma tuttavia disponibili. Ora mancano i prodotti più ricercati e di nicchia, ma ho rimediato acquistando la spesa su Amazon pantry. Cerco di rimanere serena e positiva, ma sarebbe sciocco negare che noto delle ripercussioni sulla mia salute: dormo male, ho problemi al tratto grastrointestinale, etc.* (On average, even in the smallest countries you can always find almost everything - pharmacies, foodstuffs, gifts. Unfortunately, some sectors are gradually disappearing (farmers and breeders who sell their products directly) … Initially pasta, yeast, lactose-free products, flour [were hard to find]. Now it remains more difficult to find the yeast, but with a little luck it is. Fruit, Vegetables and Meat are poor and expensive, but still available. Now the most sought after and niche products are missing, but I made up for it by buying groceries on Amazon pantry. I try to stay calm and positive, but it would be foolish to deny that I notice the repercussions on my health: I sleep badly, I have problems with the gastrointestinal tract, etc.) (Female in her 30s living in Southern Europe; Supplies Subscore 64.1; #1-3733)

In the African region, respondents with childcare responsibilities were twice as likely to (qualitatively) report struggles to access food; in Latin America and the Caribbean, respondents with elder care responsibilities were 50% more likely to mention struggles with accessing food (data not shown). Complicating the procurement of food and cleaning supplies is *Panic buying of supplies (Code 6.e.2).* Respondents were aware of larger economic and behavioral dynamics that created challenges with access to food and supplies throughout the world.

> *Nous n’avons aucunement les moyens de faire les provisions.. d’abord sans caronavirus on mangeait a peine deux fois par jour donc imaginez…* (We do not have the means to stock up … first, without coronavirus we barely ate twice a day, so imagine …) (West African respondent; Supplies Subscore 53.1; #2-1452)
>
> *Las personas exageraron y compraron mucha comida y hubo desabastecimiento de ciertos productos…* (People exaggerated and bought a lot of food and there was a shortage of certain products). (Woman in her 30s living in South America; Supplies Subscore 64.1; #1-3907)
>
> *El problema es que, la misma gente, irresponsable, se avalanzó en multitudes a comprar carros llenos de mercadería y tuvo que implementarse la medida de que, de acuerdo a los metros cuadrados de los locales, se restringió y aún hoy se hace, la entrada de 1, 2, 3 o 4 personas. Los demás, debemos hacer cola en la calle, con barbijos y respetando la distancia entre unos y otros. Me abastecí y me abastezco de lo necesario para una semana o diez días.*(The problem is that the same people, irresponsible, rushed in crowds to buy carts full of merchandise and the measure had to be implemented that, according to the square meters of the premises, the entrance was restricted - and is still done today - for 1, 2, 3 or 4 people. The rest of us must queue in the street, wearing masks and respecting the distance between each other. I stocked up, and I stock up on what I need for a week or ten days.) (Woman in her 60s living in South America; Supplies Subscore 42.2; #2-1934)
>
> *La chiusura forzata di un paese già sottomesso ad un regime predatorio in cui chi governa guadagna milioni di euro e i cittadini muoiono di fame, ci è stato tolto il diritto al lavoro. Chi non lavora non guadagna e non può vivere… La gente terrorizzata inizialmente ha depredato i supermercati che non sono riusciti a rimpiazzare i prodotti mancanti.* (The forced closure of a country already subjected to a predatory regime in which those who govern earn millions of euros and the citizens die of hunger, the right to work has been taken away. Those who do not work do not earn and cannot live … Terrified people initially pillaged supermarkets which failed to replace the missing products.) (Southern Europe respondent; Supplies Subscore 53.1; #2-1737)
>
> *Early in the pandemic panic buying resulted in no rice, pasta or meat in shops.* (Man in his 70s living in Oceania; Supplies Subscore 64.1; #2-393)
>
> *No work, no physical contact with family members, initial food shortages, travel ban… Not permitted to work as business deemed non-essential… Panic buying led to initial food shortages, Panic buying led to unavailability. I do admin so can work from home. I found scaremongering, so cut back on social media.* (Woman in her 50s living in Northern Europe; Supplies subscore 64.1; #1-1255)

The Supplies Subscore domain was one of two subscores correlated with accessing non-COVID-19-related medical services (*Struggle to Access Non-COVID-19 medical services (Code 6.b)*), which, while relating to different social dynamics that consumable supplies and food, still left respondents without access to something they felt they needed.

> *Sono diabetica, devo fare controlli periodici all’ospedale. Non posso vedere il mio medico e le prescrizioni mediche adesso vengono fatte per email.* (I am diabetic, I have to have periodic checks at the hospital. I can’t see my doctor and my prescriptions are now done by email.) (Woman in her 20s living in Southern Europe; Supplies Subscore 42.2; #1-3626)
>
> *Fear of the unknown… Shortage of equipment… No one knows exactly what can stop this totally. Losing income by closing business. (*Man in his 50s living in Southern Africa; Supplies subscore 42.2; #2-897)
>
> *I am a fulltime carer of my adult disabled daughter. We are used to isolation. It has impacted some important therapies for her tho.* (Woman in her 50s living in Oceania; Supplies Subscore 42.2; #2-151)
>
> Si me preocupa porque hay mucha gente, que no puede darse el lujo de quedarse en casa, en mi caso no tengo ese problema, pero se que hay muchos pasando hambre, eso traera como consecuencia gente en la calle y el virus se expandira mas…Mensualmente busco en un hospital medicamento para mi hermano, que vive como a 8 horas de aqui. y a raiz de esto no los he podido buscar y si los tuviera, tampoco los pudiera enviar. (Yes, I am concerned because there are many people who cannot afford to stay at home - in my case I do not have that problem, but I know that there are many going hungry that will result in people on the street and the virus will spread more… Monthly I look for medicine in a hospital for my brother, who lives about 8 hours from here and as a result of this I have not been able to search for them and if I had them, I could not send them either.) (Man in his 60s living in South America; Supplies Subscore 42.2; #1-4181)

#### Qualitative Code correlation with Cancellation Impact Subscore

The Cancellation Impact Subscore included items such as deciding not to travel or changing travel plans, postponing or cancelling a medical procedure or surgery, or cancelling plans to attend large gatherings (like concerts or sporting events). As with several other subscores and the Total Impact Score, *Worse Mental Health (Code 8)* was strongly correlated with Cancellation subscore, often linked with decreasing income and consequences of maintaining social-physical distancing:

> *Ansiedad, incertidumbre por el futuro propio y de mis hijos… Pérdida de ingresos, alteración del curso académico de mis hijos, no ver a mi familia. Cancelación de reservas de vacaciones. Dificultad para hacer planes de futuro, estudios y trabajos en curso se han visto alterados. Es una llamada a prestar atención a la globalización.* (Anxiety, uncertainty about my own future and that of my children … Loss of income, alteration of the academic course of my children, not seeing my family. Cancellation of vacation reservations. Difficulty making plans for the future, studies and work in progress have been altered. It is a call to pay attention to globalization.) (Woman in her 40s living in Southern Europe; Cancellation Subscore 62.3; #1-4032)
>
> *Classi interrotte al college…La vita ha dovuto fermarsi completamente per affrontare questa pandemia. Il distanziamento è essenziale per impedire l’avanzamento della pandemia. Ho provato molta ansia e panico. Molto preoccupante e disperato in tutto il mondo.”* (Classes interrupted in college …Life has had to come to a complete standstill to deal with this pandemic. Distancing is essential to prevent the pandemic from advancing. I experienced a lot of anxiety and panic. Very worrying and desperate all over the world.) (Male in his 20s living in South America; Cancellation Subscore 62.3; #1-3442)
>
> *He llegado a tener noches en las que no puedo dormir, debido a la ansiedad que me provoca el aislamiento social…Creo que esta situación se ha salido de las manos de cualquier gobierno, sea local o nacional…Me preocupa que la inseguridad y violencia aumente en el país, debido a que habrá más gente sin trabajo con la necesidad de comer…Me encuentro sin laborar en casa y sin posibilidades de encontrar algún trabajo durante algunas semanas o meses…Realmente no he hecho compras de pánico y he cancelado casi todos mis planes sociales. Mis sentimientos han sido demasiado ambiguos y ambivalentes, he tenido muchos altibajos emocionales, pero siento que todavía está todo bajo control.* (I have come to have nights when I cannot sleep, due to the anxiety caused by social isolation … I think this situation has gotten out of the hands of any government, be it local or national … I am concerned that the insecurity and violence increase in the country, because there will be more people without work with the need to eat … I find myself without working at home and without the possibility of finding a job for a few weeks or months … I have not really done shopping panic and have canceled almost all my social plans. My feelings have been too ambiguous and ambivalent, I have had many emotional ups and downs, but I feel like everything is still under control.) (Man in his 20s living in Central America; Cancellation subscore 51.4, #1-3942)

Some respondents indicated that – even while mental health was worsening – they were primarily coping and, as a result, had lower Cancellation subscores:

> *Fallimento totale delle mie attività imprenditoriale (attività di ricettività turistica, attività di consulenza in immobiliare). Ho annullato una visita per l’idoneità al porto d’armi sportivo. Ho annullato le mie regate di canottaggio. Ho annullato le visite per la mia operazione alle emorroidi. Fallimento totale delle mie attività…Sto cercando in tutti i modi ulteriori fonti di guadagno e confido nella mia possibilità di farlo. Mi tengo impegnato tutto il giorno e mi alleno tutti i giorni e questo mi aiuta molto a guardare avanti con positività. senza lo sport e senza la fiducia nelle mie capacità, proabilmente mi sarei depresso in questi 2 mesi*. (Total failure of my entrepreneurial activities (tourist accommodation activities, real estate consultancy activities). I canceled a visit to be eligible for a sports firearms license. I canceled my rowing regattas. I canceled visits for my hemorrhoid operation. Total failure of my businesses … I am looking for additional sources of income in every way and I am confident in my ability to do so. I keep busy all day and train every day and this helps me a lot to look forward positively. Without sport and without confidence in my abilities, I probably would have been depressed in these 2 months.) (Male in his 30s living in Southern Europe; Cancellation Subscore 40.6; #1-3749)

For some, cancellation of personally or culturally important events added to feelings of isolation and impact:

> *O medo de contrai infecção e contaminar alguem…Porque de facto ha separações de familias,redução dos rendimentos,desglobalização…Adiei viagem e festa de casamento da minha filha…Fique em casa.* (The fear of contracting infection and contaminating someone … Because in fact there are separations from families, reduced income, deglobalization … I postponed my daughter’s trip and wedding party … Stay at home.) (Woman, 50s, Sub-Saharan Africa, Cancellation subscore 40.6; #2-342)
>
> *More worried about my husband who is having difficulty sleeping and is anxious. He finds it difficult to relax… My husband and I are both retired…We paid for our children and grandchildren to accompany us on a holiday…We have to use it or lose it. This is worrying… We cannot visit our children and grandchildren. I have a heart condition and worry about shopping. I get stressed about my husband because he is a quiet man but is now constantly busy and tired. He appears unable to relax…* (Woman in her *70s,* living in Oceania; Cancellation subscore 51.4; #1-58)

One additional subgroup seemingly differentially impacted by Cancellation items was immigrant groups, most of whom are unable to travel:

> *The impossibility of returning to my country of origin stresses me. Feeling of being drifting, lonely and isolated. World economic [situation] is stressing me. Fear that this pandemic will generate a worse tragedy, hunger. We became hostages to a virus. Being in another country is even worse… Stores serve online and with deliveries. Most voluntarily closed or reduced the service that is scheduled. Parks, cinemas and interactive activities have all been canceled… I think that soon the world population will rebel against the lockdown decreed in the world*. *Especially if they’re hungry.* (East Asian respondent; Cancellation subscore 62.3; #2-919)

#### Qualitative Code correlation with Livelihood Impact Subscore

The Livelihood Impact Subscore contained items such as experiencing bullying because of coronavirus, quitting a job, and losing income from a job or business. Much of the correlation of qualitative codes with the Livelihood subscore surrounds issues of employment and work, and the changes brought about by COVID-19 prevention measures implemented by governments and employers. For some, these policies simply halted employment or educational options leading to livelihood shifts:

> *I quit my job… Due to long travel and there is no transportation available due to corona virus outbreak. (*Woman in her 30s living in South Asia; Livelihood subscore 67.6; #1-1108) *Con casi 8 semanas de encierro creo que he estado bajo de ánimos en algunos momentos, ademas que con las restricciones que hay en España las semanas anteriores estaba temeroso incluso de salir al supermercado…Estaba acostumbrado a tener una vida social muy activa y eso se ha visto bastante afectado. De momento no tengo trabajo, justo antes de la contingencia estaba por ser contratado en una empresa y eso se ha puesto en pausa…Me he sentido impotente de no poder hacer nada al ver a tantas personas pasando grandes dificultades.* (With almost 8 weeks of confinement I think I have been in low spirits at times, in addition to the restrictions that exist in [] in the previous weeks I was afraid even of going to the supermarket … I was used to having a very active social life and that it has been quite affected. At the moment I do not have a job, just before the shutdown I was about to be hired in a company and that has been put on hiatus … I have felt powerless of not being able to do anything when seeing so many people having great difficulties.) (Man living in Southern Europe; Livelihood subscale 67.6; #2-1767)
>
> *Perte de mes revenus (emploi étudiant) + annulation de mon stage + fermeture de mon université…*(Loss of my income (student job) + cancellation of my internship + closure of my university …) (Woman in her 20s living in Western Europ;, Livelihood subscale 67.6; #2-1576)
>
> *Ho avuto un forte calo delle mie entrate perché sono un lavoratore autonomo e non sono in grado di lavorare. Avevo un po ‘paura di ciò che riserva il futuro.* (I have had a sharp drop in my income because I am self-employed and am unable to work. I am a little afraid of what the future holds.) (Male in his 20s living in South America; Livelihood sub-score 61.0; #2-1659)

In the case of some respondents, the intersection of the positive association of *Changes in work/ employment (Code 7)* with the inverse association with *Government preparedness (Code 12)* reflects both the inability to work from home and an inflexibility that increases exposure and impacts livelihood:

> *La politica de estado es desinformar a la población y autoridad[e]s de salud de la region. Lo cual afecta a la comunidad nacional e internacional…Siguen programando clases de presencia fisica y exponen a trabajadores y alumnos. Minimizan las consecuencias de la pandemia… La vida ha cambiado. No hay fuentes de empleo y la estancia en casa es muy prolongad. Perd[í] mi empleo.* (The state policy is to misinform the population and health authorities of the region. Which affects the national and international community … They continue to schedule physical presence classes and expose workers and students. They minimize the consequences of the pandemic … Life has changed. There are no sources of employment and the stay at home is very long. I lost my job.) (Central American woman in her 40s; Livelihood subscore 54.5; 2-2019)
>
> *I worried due to lockdown most of the business are running in loss. It will certainly effect my job. Maybe my company will layoff employees to avoid further loss… The government is effective taking strict measures to implement social distancing. It has also helped to maintain the supply chain of essential goods… Apart from above, I also worried about the world economy and I assumed we might have to face an economic recession… As part of my wor[k], I had to travel a lot but after the coronavirus outbreak, my work and business has totally stopped. My work is my main source of income and after the lock-down I don’t have any reliable source of income… I’ m not earning after the lockdown as my office is closed. I have been staying a home all day. Avoiding all the social contact. I only go out to buy groceries wearing mask. My work often is related with outdoors marketing and it’s is not possible to do it from home.* (Man in his 30s living in South Asia; Livelihood subscore 54.5; #1-1514)
>
> *Ils ont des politiques de sante qui reste à desirer car ils sont plutot interesses par les profits qu’ils auraient grace a cette aux dons et autres appuis financiers internationnaux que de soucier reellement de leurs peuples… Le couvre feu l’etat d’urgence et le confinement ont impacté sur le revenu d’un ouvrier informel comme moi…* (They have health policies which remain to be desired because they are rather interested in the profits which they would have thanks to this with the donations and other international financial support than to really worry about their people … The curfew the state of emergency and confinement impacted the income of an informal worker like me …) (Sub-Saharan Africa respondent; Livelihood subscore 67.6; 2-1448)
>
> *Dato che si tratta di una situazione pandemica, a volte mi ritrovo a pensare che non finirà mai, che siamo in una situazione complicata e anche peggiore in [country] con un presidente che nega i fatti del covid-19 e non fa nulla per aiutare la sua popolazione…il governo nel mio paese non fa quasi alcuno sforzo per affrontare la pandemia, il presidente non si preoccupa della popolazione ed è contrario a tutte le misure di isolamento e di fronte alla covid…Ho dovuto interrompere i miei studi all’università e ho dovuto lasciare il mio tirocinio perché non volevano fermarsi ma per proteggermi ho deciso di rimanere a casa e lavorare a Mturk. il mio lavoro non poteva essere svolto a casa…* (Since this is a pandemic situation, I sometimes find myself thinking that it will never end, that we are in a complicated and even worse situation in with a president denying the facts of covid-19 and doing nothing to help its population… the government in my country makes almost no effort to deal with the pandemic, the president does not care about the population and is against all isolation measures and facing COVID … I had to stop my study at university and I had to leave my internship because they didn’t want to stop, but to protect myself I decided to stay home and work in mTurk. My work could not be done at home …) (Man in his 20s living in South America; Livelihood subscale 67.6; #1-3409)

Many respondents remain stressed about work, particularly those whose employment depends on in-person interaction with clients or markets:

> *My business may go bankrupt and with it my income and my home… The main business cannot be done from hom[e]. I’m in the high risk age group and I take care of a 90 year old, I can’t risk [shopping for groceries]… Unable to pay my debts and have money for food and needs.* (Woman in her 60s living in the Caribbean; Livelihood subscale 54.4; #1-1955)
>
> *Soy freelancer y estoy acostumbrado a pasar mucho tiempo en casa, ovbiamente hay diferencias pero estoy acostumbrado a traba[ja]r desde casa y pasar mucho tiempo en ella…La situación de la gente que depende del día a día y no está pudiendo desarrollar sus actividades…La bajada de cantidad de clientes y trabajo repercute en la economía y está un poco fuera de mi control resolverlo. La convivencia contínua también genera estrés y roces.* (I am a freelancer and I am used to spending a lot of time at home, obviously there are differences but I am used to working from home and spending a lot of time in it … The situation of people who depend on the day to day and are not being able to develop their activities…The decrease in the number of clients and work affects the economy and it is a little out of my control to solve it. The continuous coexistence also generates stress and friction.) (Man in his 20s living in South America; Livelihood subscore 67.6; #1-3854)
>
> *Difficulté de ce nourrir même d’électricité chez nous et si jamais je tombe malade d’un autre maladi même je ne peux pas me soigne par-ce-que je vie du jour au jour et que c’est le pire encore que je vie maintenant…Ma mère est partie au marché pour chercher de quoi a mangé et on lui a arrêté pour l’amène à la gendarmerie pendant 24h et payer avant d’être libéré…Ma profession ne peux pas me permettre car je suis un homme du terrain.* (Difficulty of even feeding electricity at home and if I ever fall ill with another disease even I cannot cure myself because I live from day to day and that it is the worst yet that I live now … My mother went to the market to look for something to eat and she was arrested to take her to the gendarmerie for 24 hours and pay before being released … My profession cannot allow it [work from home] because I am a man of the field.) (Man in his 30s living in Sub-Saharan Africa; Livelihood subscale 80.6; #2-1402)

A strong inverse correlation of *Primarily unaffected/coping (Code 9)* qualitative cluster with Livelihood subscore suggests that some respondents perceive that their lives have not dramatically changed or that they are adjusting to the changes sufficiently:

> *I trust in God’s presence in this epidemic, my gov’t is taking care of the medical and economic emergency, people are following safety protocols… Some people are willfully disobeying this protocol, endangering not only themselves but others who are doing it. This kind of irresponsible behavior is deplorable and should be sanctioned…* (Woman in her 50s living in Southeast Asia; Livelihood subscale 41.5; #1-1758)

Similarly, other respondents note that the origins of their livelihood shifts are unnecessary, and in some cases, considered oppressive or unfair:

> *I am 70 with no underlying health issues but because of my age I have to isolate for 12 weeks, this is having a massive impact on my life, health and mental health. Mainly really angry and worried at having to stay at home, I need to go out. Because of my age I feel as if I am being bullied by government lockdown requirements. I am doing what I have been asked to do, but extremely unhappy about not seeing my family, I am an adult capable of making my own decisions. I feel very, very stressed and angry having to stay at home, I need to go out to visit family and friends.* (Woman in her 70s living in Northern Europe; Livelihood subscale 67.6; #1-2237)

Conversely, essential workers are typically unable to work from home, and more intimately interact with the very COVID-19 risks that the rest of the community tries to avoid, and the elimination of which are the goals of stay-at-home orders and shutdowns for other workers:

> *I stayed home and got covid 19 anyway. My husband is front line medical and came home sick with it as well. If people will do all recommended then it would have been checked. But they won’t. Just because there are more beds and equipment doesn’t mean you should fill them up. We need to stay home and use protective equipment. People are dying. Young and old*. *Tattoos, massages, bowling are not worth dying for. People do not take it seriously. That is why it continues. That is why our health care workers get sick. God have mercy on us all. Yes some are bored. We all want to get out. We all want to go back to work. But please protect us all. God have mercy… We have it. So guess all the precautions we took still did not prevent it. But my husband who I think brought it home is a paramedic. All front line medical are more susceptible. Our friend a nurse has it too. Another medical worker tested positive… My husband is on a ventilator with covid 19. He is our major worker. Yes I am concerned for his health and our welfare. (*Northern America respondent in their 60s; Livelihood subscale 67.6; #1-2812)

#### Total Impact Score by Region

Non-medical impact as a consequence of COVID-19 and its prevention unfolds throughout the world in many similar ways – respondents in all regions note the difficulty in obtaining supplies and provision, the personal actions necessary to address and adapt to shutdowns and rapid change, cancellations of personally important and culturally relevant events, and livelihood shifts brought on by changes in job and school, even though the context and specifics of the ecological framework may vary considerably by region.

#### Africa

> *Just staying at home during lockdowns without working and earning at the end of the day, week or month… Restricted on my movements using public transport…. The loss of my current job and no proper business as before… Very little part of my job will be done at home….(*Man in his 30s living in Sub-Saharan Africa; Total Impact Score 67.5; #2-127)
>
> *Fear and anxiety engulfs me and my family thus, the emotional and psychological stress. This also has also drained us financially as family because we cannot go to work as before and market has also gone down drastically because people are financially handicap. Worried about closing down of schools and churches. Some female students are carelessly getting pregnant all because they are at home because of the Coronavirus. Financially down, getting less, spending more. More food is consumed when kids and family are at home. More light and water bills consumed. Work down and sales also down. I am a field officer. Without school in session, it would be impossible to move round to supervise activities and others as mandated.* (Man in his 30s living in West Africa; Total Impact Score 43.9; #2-895)
>
> *Changement radical dans façon de vivre et avenir incertain…Confinement inefficace, moyens de protections insuffisants, population mal informée …Ne peut plus aller voir mes parents, un enfant bloqué a l’ étranger, incertitude de l’ avenir, peur de violences si situation incontrôlée…Beaucoup moins important qu auparavant, une sorte d adaptation en cours…*(Radical change in way of life and uncertain future … Ineffective confinement, insufficient means of protection, poorly informed population … Can no longer go to see my parents, a child stranded abroad, uncertainty of the future, fear of violence if an uncontrolled situation … Much less important than before, a sort of adaptation in progress …) (Woman in her 50s living in Northern Africa; Total Impact Score 47.9; #1-3311)

#### Asia

> *Corona is creating a worldwide panic situation. It is increasing anxiety among people that may result in stress… I can’t go outside the home or be social with friends. This has a negative effect on my mental health… I have canceled my traveling plans and a meeting with my doctor too due to this situation…. In [], provinces are under lockdown so you can get the groceries within a given period of time. so it is hard to get everything. most people have stocked such items. So it is difficult for us to get supplies on time… Due to the disturbance in my social life, I was very much on stress.*(Man in his 20s living in South Asia; Total Impact Score 61.0; #1-1675)
>
> *When you worry so much, you won’t be able to think of the productive ways to start or spend your day… The system was caught unprepared for this pandemic. There is lack of PPEs. Given the novelty of the viral infection, the system is not backed up by adequate knowledge on how to combat it… I still continue working as a teacher while staying at home. I prepare lessons and activities for my students and send these through messenger and email. Ours is a boarding school and student teachers are handling classes using the activities that teachers prepared. I am only away from my family. I miss being together with them… You need to walk for an hour to go to grocery store. Another hour to get back. Public transportation is not allowed… I avoid going to the doctor to get prescriptions…(*Man in his 40s living in Southeast Asia; Total Impact Score 59.7; #1-1835)

#### Europe

> *Ineffectual management of the situation by [] governments is having a hugely negative impact on the economy. My business is in the construction industry. Business unable to trade, loss of revenue stream. Widespread food shortages. The lockdown measures led to stockpiling which the supermarkets and government were ill equipped to remedy. Sanitizer has been unavailable since February. Subject to verbal abuse for leaving home. Public hysteria is such that many now labour under the misguided notion that stepping outside will lead to certain death. The media have exacerbated this and the government have done nothing to curb it. Ineffectual and inappropriate responses by the [] government will have grave repercussions for the economy and health of the nation… I do not believe lockdown measures to be effective in the long term due to the economic and social ramifications and the fact that it merely delays the inevitable. I feel that the government should focus resources on increased testing and should look to emulate the strategies deployed in the countries that are controlling the spread rather than relying in flawed scientific assumption.* (Woman in her 40s living in Northern Europe; Total Impact Score 59.7; #1-2211)
>
> *I don’t worry. I’m enjoying the peace. I’m learning and working on my computer. There are no disturbing factors… I think the [] government is very effective in fighting the crisis. Restrictive measures were introduced at an early stage. 36 thousand beds are there in hospitals to receive the infected. The [neighboring] government was slow to react. That’s why there have been nearly 15 thousand infected people up to this moment but now the situation is managed well by the healthcare system. I’m not worried. I’m generally commuting between [] and []*. *Borders are now closed for private traffic. I have to pay the rent for an apartment in [] that I cannot use now until the border is closed. The contract cannot be cancelled because in these circumstances I cannot empty the apartment. A major exhibition of my daughter has been cancelled that would have been a large gathering. I did not have plans to visit other big events. Everything is available in the store opposite our house. There is an abundance of goods. I go out once in two weeks to buy food. I have not been irritated at all. I miss seeing my mother who is 90 years old and lives 200 km far from us.(*Woman in her 60s living in Western Europe; Total Impact Score 59.7; #1-109)
>
> *Always thinking at coronavirus-related things, constantly worried for friends/family, feeling like crying all the time, irascible, sad, helplessness feeling, unproductive… Not able to take my usual walks, not able to volunteer where I was, university closed, no alone time due to family staying home too, lost routine and usable to focus or be productive, chaotic sleep schedule*. *Going out to buy groceries only once a week so buying for the whole week, not panic-buying stocking up. Postponed ophthalmological consult and new glasses, routine health check and lungs check.* (Woman in her 20s living in Eastern Europe; Total Impact Score 59.3; #1-2005)

#### Latin America and the Caribbean

> *Las deudas siguen aumentando y muchos quedaremos en la calle…Soy independiente y al quedar encerradoy aislado mi economía esta en problemás más graves que antes. Me estan cobrando renta de vivienda y de donde carajo la saco. Vendo servicios legales enun país donde la brecha digital es grande…No tengo a nadie y ya estoy hastiadode esto. Estoy estresado,deprimido y con un miedo terrible de quedar en la calle.* (Debts continue to increase and many of us will remain on the street … I am independent and by being locked up and isolated, my economy is in more serious problems than before. They’re charging me housing rent and where the hell do I get it from? I sell legal services in a country where the digital divide is great … I have no one and I am already fed up with this. I am stressed, depressed and terribly afraid of being on the street.) (Central America respondent; Total Impact Score 55.7; #2-2319)
>
> *El mundo está paralizado, no hay trabajo, la situación económica de mi pais y del mundo entero esta en estado crítico y eso es lo que me preocupa…Mi pais es un desastre económico y sanitario debido a la incompetencia del régimen dictatorial en que vivimos…Los planes a corto y mediano han sido totalmente destruidos y nos ha tocado sobrevivir en un ambiente lleno de estrés y pesimismo…Trabajo como freelancer y no he tenido contratos o actividades de trabajo en lo que va de cuarentena… El presupuesto de mi familia era muy escaso cuando impusieron la cuarentena. El mundo exagera con esta enfermedad, es mas el pánico que genera que las muertes.* (The world is paralyzed, there is no work, the economic situation of my country and the whole world is in critical condition and that is what worries me … My country is an economic and health disaster due to the incompetence of the dictatorial regime in which we live … The short and medium plans have been totally destroyed and we have had to survive in an environment full of stress and pessimism … I work as a freelancer and I have not had contracts or work activities so far in quarantine … My family’s budget was very tight when the quarantine was imposed. The world exaggerates with this disease, it is more the panic that it generates than the deaths.) (Woman in her 20s living in South America; Total Impact Score 59.7; #1-4194)

#### Northern America

> *[Our region] has been wonderful, sharing information quickly, shutting down relatively quickly, and so on. The national government continues to refuse to order a nationwide shelter in place. The scientists are doing a good job of disseminating information but they are hampered and unable to implement plans for action. We have a national government who dragged its feet getting items from the national stockpile to hard hit areas then said the national stockpile is not for the country to use…??? I’m worried about local, small businesses. I’m worried about people who are in bad home situations that have to shelter in place. I’m worried about a second and third wave. I’m a K-12 teacher now teaching from home for the rest of the school year. My state is talking about if we open up in the fall, staggered schedules and more physical distancing. I like to go out in the world after work and on days I don’t work, soaking up art, culture, nature, and more. I travel frequently. All of that is gone for now. During the first two weeks of shelter in place I was anxious and fearful then I calmed down. I reminded myself that as long as I follow precautions when I’m out (which is really rare) and I shelter in place, I’ll be okay. I shifted my attitude and started looking at this time as a gift. I re-cultivated gratitude. (*Woman in her 40s living in Northern America; Total Impact Score 55.7; #1-2606)
>
> *We have two essential workers, so they wash hands frequently. One has more direct contact with public so he washes frequently and washes his clothes and takes a long hot shower after shift. I don’t mind the quarantine. I worry about my husband and son. [Our] governor has been fantastic. Informative, blunt. Federal government is promoting denial. Jobs closed, schools closed, stores closed and I haven’t left my house in three weeks. My elderly mom is used to being fairly active and I’m afraid it will age her more rapidly. (*Northern America respondent; Total Impact Score 58.1; #1-2488)
>
> *I have generalized anxiety and I am recovering from major depression. I know how to manage “regular” stress, but the pandemic is throwing a wrench in everything. My coping mechanisms are affected by the social distancing measures put in place, the constant news updates about deaths rising and dramatic repercussions are taking a heavy toll. I work in healthcare, so the stress is also heightened significantly at work. The routine is significantly modified too, which doesn’t help with anxiety management. Local government is reacting faster than national, but the measures are not always as effective as they should be. Some decisions are taken too late. Massive changes at work. Constant stress all the time. Routine is upside down. The lines in front of stores are long and since I am still working I have no time to waste waiting in line. I made do with what I have at home as much as I can and have to resign myself to not getting what I want… Some stores are starting to have cleaning supplies and hand sanitizer again, but it was out everywhere for over a month… Things are a mess, they will stay messy for a long time and no amount of rainbow will change anything about that. I understand the need for positive thinking, but it seems like a delusion in many cases…(*Woman in her 20s living in Northern America; Total Impact Score 55.7; #1-382)

#### Oceania

> *I’m afraid to go to work. And yet I have to work. I stayed at home, no work no pay company policy. In a hospital where there are many patients [and] coronavirus might be airborne…(* mid 50s, woman, Pacific Islands, Total Impact Score 57.3; #1-1623)
>
> *Fear of losing job and getting infected and death… I just lost my mother and I can’t travel and never see her again. She died alone in the hospital. This is very tragic and heartbreaking to us all in the family. I need to travel but I’m terrified with the virus and anxious to get quarantined in a prescribed place other than my house. I lost my dear mother and I couldn’t travel to be with her.* (Woman in her 50s living in the Pacific Islands; Total Impact Score 63.6; #1-1620)
>
> *Anxiety every now and then. Sleep has been totally off…because we are home all day… sometimes is hard to sleep at bedtime because I just think of everything….it’s harder like when we hear another person died….anxiety kicks in. Been praying more too… Worried about everything… This pandemic is real….we realize the world is hurting…we want to try and get back to normal…..then we have to think….will we ever get back to normal…is the new normal always wearing masks to be better protected….the fear of the unknown….. I teach….we now have to teach online. My children wanted to join 4th qtr. sports…that’s not happening.. I was also a teacher at church on Sunday…nobody can attend mass…we attend by watching it live on social media…The fear of the unknown….so many things we had planned but now we don’t know…. I chose to cancel my daughters follow-up appt. at our local hospital because we already had patients there with the virus….I was just scared and didn’t even want to risk us from getting it… My stress level was high when we 1st locked down or rather shutdown and had to stay home… social distancing…my stress was from not knowing what was to happen in the days to come…..I would say my stress level has subsided a little knowing that our Dept. of Education has a plan and we now have guidance.* (Woman in her 40s living in the Pacific Islands, Total Impact Score 59.6; #1-687)

## DISCUSSION

The personal, non-medical impact of COVID-19 is substantial around the world, disproportionately impacting people, households, and countries in a wide context of influences. The personal non-medical impact includes actions people have taken, cancellations of events and plans, inability to procure food and supplies, and personal economic/ employment/ social disruptions. The global policy and public health reactions to control spread and impact of COVID-19 have largely focused on shutting down public places, restricting movement and gathering of people, providing testing, screening, and treatment for the disease, and implementing infectious disease precautions (masking, social-physical distancing, handwashing). Although these measures are highly effective – and in some countries such as New Zealand, have contributed to an increase in trust among citizens – they have also exacerbated pre-existing inequalities from socio-economics to digital and technological inequalities. (56, 57) Lockdowns have affected socio-economics globally, including reductions for governmental revenues leading to social welfare reductions, thereby increasing societal poverty gaps. (58) Shown in Figure 5, our analysis contextualizes the constructs and dynamics of non-medical implications of COVID-19-related policies, behaviors, and influences within a medical ecological framework that operates at multiple levels and across multiple dimensions, allowing for a more comprehensive analysis of these phenomena.

**Figure 5:**
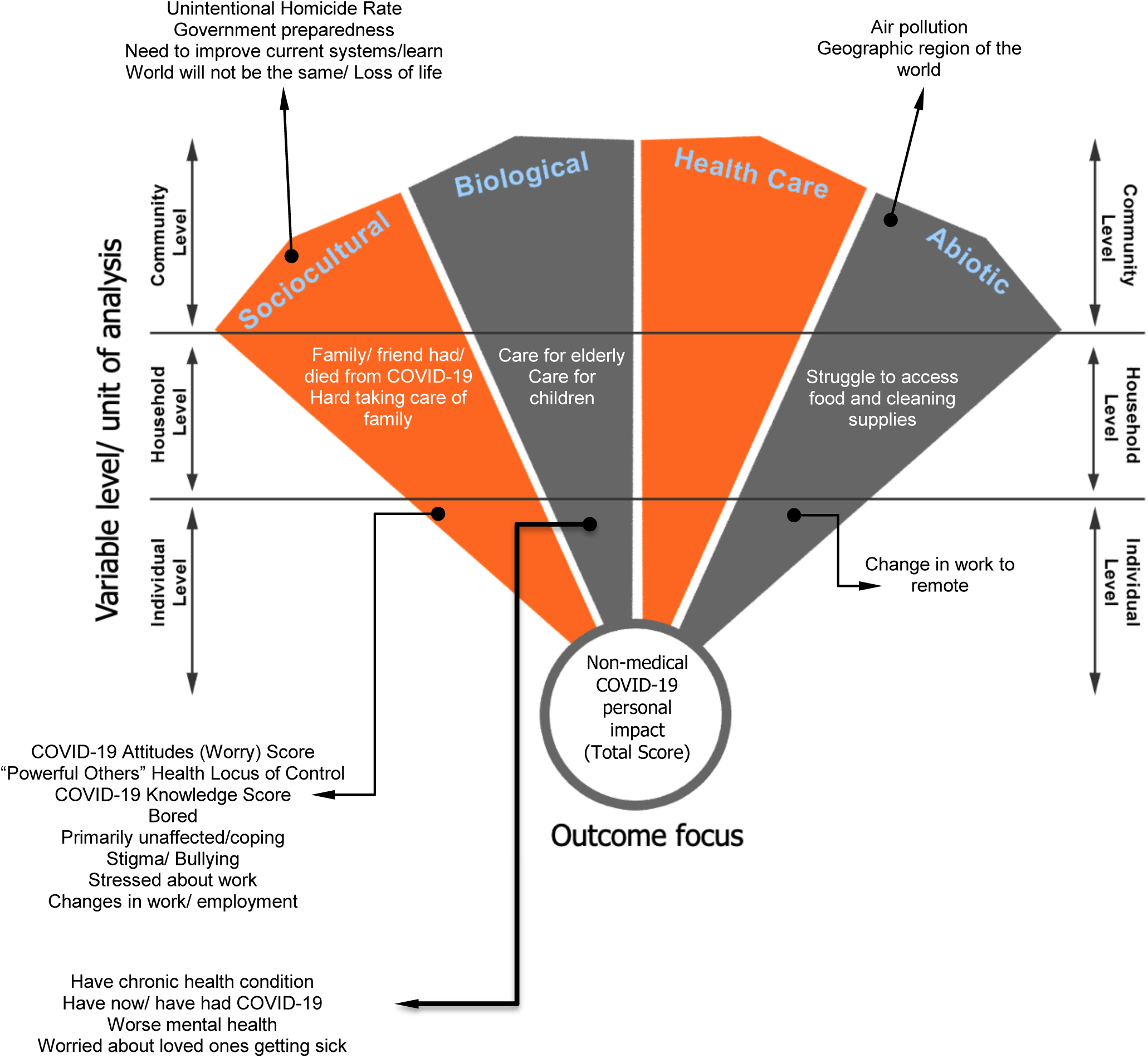
Significant quantitative and qualitative associations with non-medical COVID-19-related personal impact (Total Score) at the global level mapped to the Critical Medical Ecological mode.

Non-medical impact of COVID-19 varied significantly among regions, with respondents from Asia noting the highest impact, followed by Latin America and the Caribbean, Africa, Northern America, Oceania, and Europe. The subcomponents of impact often showed similar geographic patterns: Asia and Latin America had the highest levels of personal action, Asia, Latin America, and Africa had the highest levels of cancellations, and Asia, Latin America, and Africa had the highest levels of livelihood impact. The only subcomponent of impact that varied considerably from this general geographic pattern was “supply” impact, being the highest in Oceania and Northern America and lowest in Latin America and Africa. The cluster of countries in Asia, Latin America, and Africa also had the highest levels of “internal” and “powerful others” health locus of control, the highest levels of COVID-19-related worry, and the highest rates of difficulties accessing health care. While the COVID-19 rates of infection and death were lower in this cluster of countries during the time of this study, the differential non-medical impact of policies aimed at containing the pandemic - resulting in higher levels of total non-medical impact, personal action impact, cancellation impact, and livelihood impact - were greater. With many former colonies and historically subjugated peoples residing in this cluster of countries, contemporary effects of past colonization and the perpetual exploitation of one cluster of countries (Northern America, Europe, and Oceania) on the other (Africa, Asia, and Latin America) could account for some of the systems-level influences (including economic, social, political, and environmental) on non-medical impact. In our analysis, country-level influences accounted for more variation in non-medical COVID-19-related impact in Africa and Asia than they did in the other regions. That said, most global variation in non-medical impact (accounting for about 2/3rds of variation) was attributed to individual-level factors, with the remaining third split between household factors and country-level factors. Except in Oceania, country-level COVID-19 morbidity/mortality metrics remained insignificant predictors of non-medical impact after multilevel modeling.

Worsening mental health, stress, and COVID-19-related worry were the most prominently and consistently associated variables with non-medical impact in every region. COVID-19-related worry was significantly, positively associated with every impact score and subscore across every region: as COVID-19-related worry increased, so did levels of non-medical impact. Over 90% of respondents in all regions indicated that worry or stress related to COVID-19 had impacted their mental health, and respondents were at least twice as likely or more to worry about getting sick from COVID-19 than from dengue or Lyme disease. Though the main reason for this worry in each region was concern about becoming sick themselves, the remaining reasons for worry were economic: worried about income, investments, and not being able to afford to work/ stay home nor afford COVID testing. Qualitatively, mental health and worry directly accounted for three of the top five factors correlated with non-medical impact, and indirectly accounted for the other two (struggle to access food and supplies). Qualitative responses around stress and worry were very frequently coupled with concerns about the economic impact, loss of income, and inability to afford sustenance and prevention. Importantly, qualitative responses also indicated that respondents frequently questioned the purposes and consequences of policies requiring they work from home or shutting down worksites and schools when the economic impacts of such policies on families were not always addressed clearly.

In fact, it may be that the more distant a respondent is from the reality of the medical effects of COVID-19, the less likely they are to understand or to agree with national policies aimed at reducing its spread. Our findings suggest that the closer the respondent is to personally experiencing COVID-19 (being infected themselves, having a family or friend infected, having a family or friend die), the more likely they are to experience higher levels of non-medical impact. A strong, statistically significant relationship exists between lack of personal experience of COVID-19 (self, family, or friend) and considering oneself primarily unaffected, that one’s freedoms have been limited, and mentioning COVID-19-related conspiracy theories and misinformation; in general, the non-medical impact scores are lower and significant for respondents holding these beliefs. As mentioned previously, country-level COVID-19 outbreak magnitude and change over time had little impact on non-medical impact scores in most regions, but personal-level experience with COVID-19 relates to greater non-medical impact. For example, the vast majority of respondents had never been tested at the time of this study and did not believe they had been infected, and did not experience a friend or family member having or dying from COVID. While an important set of influences on people with this direct medical experience – most respondents do not have that medical experience and therefore, undergo the impacts of shutdown policies and others without that insight. Specifically, the majority (>50%) of respondents in all regions observed social-physical distancing, cancelled travel plans, bought or wore a mask, stocked up on supplies, or stayed home instead of going to work. More than 1/3^rd^ of participants lost income, and one in ten lost their jobs. Being, knowing, or interacting with someone COVID-19-infected itself is generally much less common than these non-medical impacts; for many, these are the dominant, lived dynamics of the pandemic.

The International Labour Organization estimates that during the COVID-19 pandemic, half of the global workforce (80% of workforces in Latin America and Africa, 70% in Central Asia and Europe, 22% in Asia and the Pacific) are at risk of losing their jobs. (59) COVID-19 preventive measures have had an impact on people’s loss of income such as seasonal jobs increasing instability and insecurity in many towns.(60-62) The effect of job loss significantly impacts migrant employees.(63) Additionally, migrant, rural, and ethnic minority children have been disproportionally affected by the pandemic preventive measures of school closures. The shifting from typical educational approaches to the use of social-physical distance learning has magnified pre-existing global digital inequalities. Marginalized communities lacking access to technology aggravates pre-existing children’s academic disparities.(57, 64) The school closures have faced secondary societal impacts including food access challenges where in places such as India and the United States, closing schools have represented another barrier of inequalities because many children are dependent on school meals, exacerbating food insecurity.(65, 66)

Lockdowns have also affected global travel. The COVID-19 pandemic has represented geopolitical challenges in which policies have been made to protect their country’s constituents including travel restrictions and closing access to borders as preventive measures. For example, the European Union banned United States’ citizens from entering their countries because of the inability of the United States government to control the spread of pandemic cases.(67-69) The travel restrictions imposed worldwide have also generated a chain effect in the global economy pushing arguments for a global recession.(70)

Compounding these personal experiences of non-medical impact are significant challenges in maintaining households. The COVID-19 pandemic has not been an exception in severely disrupting the global supply chain. (71) The pandemic has disproportionately affected the dependency on economic commerce across countries, reducing both supply and demand, making it challenging for communities to find and meet basic needs, such as medications, fuel (e.g., gasoline, petrol, oil), and food.(72-74) As we’ve shown, having children under 18 in the household or responsibilities for caring for elders were responsibilities significantly and independently associated with greater non-medical COVID-19-related impact. Caring for children was especially associated with higher “supplies” and “livelihood” impact, while caring for elderly was significantly and independently associated with all non-medical impact scores. Respondents indicating they had child-care or elder-care responsibilities were more likely to report (qualitatively) challenges with coping. In the African region, respondents with childcare responsibilities were twice as likely to (qualitatively) report struggles to access food; in Latin America and the Caribbean, respondents with elder care responsibilities were 50% more likely to mention struggles with accessing food. Community behaviors in response to the pandemic can be categorized by three phases, including reaction, coping, and long-term adaptation. These behaviors altered the balance of the supply chain system.(75) Access to certain protective products has become a necessity, like the accessibility of handwashing facilities and disinfectant household supplies.(76, 77) In addition, other needs have been politicized such as the use of facemasks.(78) This societal crisis has polarized communities, which have then incurred community altercations between people, such as healthcare workers being discriminated and bullied, thus aggravating mental health of first respondents and their relatives.(79, 80)

Our study is limited in several ways. Firstly, we could not access all countries of the world (e.g., China) given that we used Facebook and mTurk to recruit participants. The inability of citizens from China to participate in this project limits our complete analysis of global lived experience, but also allows other nations in Asia disproportionate visibility in their own experience. Similarly, the respondents in our sample drawn from these social media sources are likely to have better access to technology, literacy, and, likely, education than their non-responding counterparts. Secondly, our study was conducted during the early months of the pandemic; since then, much of the world has experienced at least an initial wave if not multiple waves of COVID-19, which likely could change the relative impact of variables in our models. Thirdly, we experienced high levels of missing demographic data, which limit or exclude our ability to analyze such variables as age, gender, and education, particularly in multivariate models. In using four languages, close monitoring of recruitment, and through integrating quantitative and qualitative data, we have strengthened our approach to generate more useful, novel analyses that could enhance public health effectiveness. Our large global sample with all geographic regions of the world represented allows for integrated and multilevel analyses that help us address the ecological interpretation of our data.

Respondents, households, and communities are not without assets with which to confront the challenges facing them from the COVID-19 pandemic, and COVID-related policies and prevention strategies. COVID-19-related knowledge is high: most respondents relatively accurately understood COVID-19 prevention strategies, types of transmission, and risk groups. Most respondents indicated they had sufficient information to protect themselves and their families from COVID-19 infection. “Internal” health locus of control levels exceeded “Chance” or “Powerful Others” in each region, suggesting that respondents had a strong sense of controlling their own health. Respondents indicated having mostly strong social support systems, and most reported using social media regularly. Many respondents indicated their health was good to excellent. COVID-19-related knowledge, internal health locus of control, sufficient COVID-19 information, and strong support systems could help moderate some of the challenges brought on by worry, coping, and practical impact of shutdowns. Indeed, COVID-19 prevention policies and strategies could leverage these and other assets to reduce the burden of non-medical impact and – perhaps – enhance the effectiveness of policies to prevent disease transmission and improve adherence with prevention strategies.

## CONCLUSIONS

Individual-level sociocultural variables (representing social roles, power dynamics, and stressors) were most strongly associated with non-medical COVID-19-related impact in most regions of the world. Country-level non-medical impact globally (especially in Africa and Asia) demonstrated the complex effect of social conditions, country policy, and environmental factors on individual experience, which needs further exploration to prevent unintended consequences of COVID-19 control. Household-level variables demonstrated the challenging difficulties household units face in addressing supplies, income, health, and employment.

While the cumulative medical consequences (incidence, morbidity, mortality) of COVID-19 commands priority of systems globally, the lived experiences of these policies aimed at prevention and reduction of COVID-19’s health and epidemiological consequences affect a far greater portion of the global population directly. For many, COVID-19 infection and its medical consequences remain somewhat distant while the non-medical impact of policies aimed at controlling COVID-19 impact them at the individual-level. Attending to non-medical impact of COVID-19 helps balance prevention of disease with prevention of unintended consequences of policy measures, and in fact could strengthen the ability of individuals, households, and systems to adhere to prevention and reduce the health consequences of COVID-19.

Security measures such as lockdowns and social-physical distancing have resulted in a constant adaptation by communities without taking into account pre-existing societal disparities, inequalities, and injustices, perpetuating the stigmatization of certain community groups.(81) Measures of isolation have resulted in barriers for non-COVID-19 health prevention and health conditions management (e.g., diabetes and mental health).(82-84) Social-physical confinement has also increased family and domestic violence altercations and alcohol consumption.(85-87) The social isolation measures have had negative consequences such as reduced physical activity and increased smoking. Previous work in Brazil and the United Kingdom have suggested that these behaviors could negatively impact people’s lives, particularly in those with pre-existing health conditions such as cardiovascular disease.(88, 89)

Although social confinements and social-physical distancing measures have been affecting communities, they have also contributed positively to ecological emergencies by improving the global climate such as air quality, biodiversity, and noise reduction across many metropolitan cities.(90-93) While the pandemic has changed people’s lives worldwide, it has especially impacted the most vulnerable and marginalized populations including migrant communities, refugees, indigenous groups, and ethnic minority groups that were already living with an amalgam of pre-existing social inequalities.(94-97)

Best practice in public health and infectious disease prevention can – eventually – collide with human culture and generate stress and influence that challenges the very effectiveness of public health and policy interventions. Human communities evolve to sustain themselves through family, culture, and economy – with elements of household structures, individual roles and expectations, shared community events and celebrations, food systems, economic systems, and personal relationships among many social mechanisms. Disruptions in these systems and mechanisms challenge the ability of communities – and their component households and individuals – to adhere to policies aimed at controlling disease. In short, people will – eventually – choose maintaining their families and culture over prevention when such a choice is required, and will prioritize obtaining the basic requirements of life through economic activity over what they perceive as unnecessary and very possible sickness and death in their families – from other non-COVID-19 causes - as a result.

Attending to how to balance prevention of disease with managing and sustaining culture and social expectations will increase the probability of adhering to both. In countries/regions with seemingly intransigent inequities and unbalanced global economic relationships, inability to adhere to prevention and the potential disruption of culture and society are strong. For many communities, households, and individuals, it is impossible to adhere to prevention policies given existing inequities and maldistribution of resources within countries and around the world. Through the words of one Sub-Saharan African man, *“In my country we live overnight - what we seek is what we eat…one would rather go out and contract corona than starve to death while staying at home!*” Widespread upheaval of societies and mass disruptions of social dynamics through human tragedy, natural disasters, or pandemics are likely the biggest medical ecological stressors (and social determinants of health) that exist. Monitoring and accounting for non-medical impact in policy and public health strategies while simultaneously focusing on medical impact of COVID-19 will improve the opportunity for disease reduction, through potentially moderating the unintended consequences on families, households, and countries.

## Data Availability

Though relevant data (tables and qualitative excerpts) are within the manuscript additional data cannot be shared publicly because of the risk of stigmatization of marginalized groups and countries.

## ACKNOWLEDGEMENTS

We are grateful for the assistance of Connor DeAndrea-Lazarus, Kathleen Buckwell, Cody Gardner, and Carrie Dykes for logistical assistance with developing and implementing aspects of this project.

## REFERENCES

1. World Health Organization. COVID-19 Strategy Update. Geneva, Switzerland: World Health Organization Printing Office. 2020.

2. van Dorn A, Cooney RE, Sabin ML. COVID-19 exacerbating inequalities in the US. Lancet (London, England). 2020;395(10232):1243.

3. Ahmed F, Ahmed Ne, Pissarides C, Stiglitz J. Why inequality could spread COVID-19. The Lancet Public Health. 2020;5(5):e240.

4. Marmot M. Economic and social determinants of disease. Bull World Health Organ. 2001;79(10):988–9.

5. Rose G. Sick individuals and sick populations. Int J Epidemiol. 1985;14(1):32–8.

6. Braveman P, Egerter S, Williams DR. The social determinants of health: coming of age. Annu Rev Public Health. 2011;32:381–98.

7. Cheng VC, Lau SK, Woo PC, Yuen KY. Severe acute respiratory syndrome coronavirus as an agent of emerging and reemerging infection. Clin Microbiol Rev. 2007;20(4):660–94.

8. Oxford COVID-19 Government Response Tracker [Internet]. 2020. Available from: https://www.bsg.ox.ac.uk/research/research-projects/coronavirus-government-response-tracker.

9. World Health Organization. Novel coronavirus (2019 nCoV) Advice for the Public. World Health Organization; Geneva: Switzerland. 2020.

10. 2019 GPMB. A World at Risk: Annual Report on Global Preparedness for Health Emergencies. Geneva: World Health Organization 2019.

11. U.S. Department of Health and Human Services OoDPaHP. Healthy People 2020 Washington, DC [4 Aug 2020]. Available from: https://www.healthypeople.gov/.

12. Czyzewski K. Colonialism as a broader social determinant of health. International Indigenous Policy Journal. 2011;2(1).

13. Fisher MB, Emma. As Coronavirus Deepens Inequality, Inequality Worsens Its Spread. The New York Times. 2020 March 15, 2020.

14. Alegria M, NeMoyer A, Falgas Bague I, Wang Y, Alvarez K. Social Determinants of Mental Health: Where We Are and Where We Need to Go. Curr Psychiatry Rep. 2018;20(11):95.

15. Dungca N AJ, Bhattarai A, Kornfield M On the front lines of the pandemic, grocery workers are in the dark about risks. The Washington Post. 2020.

16. Rollston R, Galea S. COVID-19 and the Social Determinants of Health. Am J Health Promot. 2020;34(6):687–9.

17. Turner-Musa J, Ajayi O, Kemp L. Examining Social Determinants of Health, Stigma, and COVID-19 Disparities. Healthcare (Basel). 2020;8(2).

18. Ndegwa S. Religion and Culture Plague Africa’s Fight Against COVID-19: China Global TV Network; 2020 [Available from: https://news.cgtn.com/news/2020-04-26/Religion-and-culture-plague-Africa-s-fight-against-COVID-19-PZNaKEiRFe/index.html.

19. Jennings R. How Cultural Differences Help Asian Countries Beat COVID-19, While US Struggles. Voice of America. 2020 July 22, 2020.

20. Yen MY, Chiu AW, Schwartz J, King CC, Lin YE, Chang SC, et al. From SARS in 2003 to H1N1 in 2009: lessons learned from Taiwan in preparation for the next pandemic. J Hosp Infect. 2014;87(4):185-93.

21. Hille KW, Edward. Containing coronavirus: lessons from Asia. Financial Times. 2020 March 16, 2020.

22. McGinnis JM, Williams-Russo P, Knickman JR. The case for more active policy attention to health promotion. Health Aff (Millwood). 2002;21(2):78–93.

23. Organization. WH. Key facts from JMP 2015 report. 2015.

24. Bavel JJV, Baicker K, Boggio PS, Capraro V, Cichocka A, Cikara M, et al. Using social and behavioural science to support COVID-19 pandemic response. Nat Hum Behav. 2020;4(5):460–71.

25. Cialdini RB, Goldstein NJ. Social influence: compliance and conformity. Annu Rev Psychol. 2004;55:591–621.

26. Normile D. Coronavirus cases have dropped sharply in South Korea. What’s the secret to its success? Science. 2020 March 17, 2020.

27. Sui C. What Taiwan can teach the world on fighting the coronavirus: NBC News; 2020 [updated March 13, 2020. Available from: https://www.nbcnews.com/health/health-news/what-taiwan-can-teach-world-fighting-coronavirus-n1153826?fbclid=IwAR3RxQndOSGbH2lW3wVg2fFJ9KkRxv0ko2tQ7X-ZY7oJs0195VOxVXlkQB0.

28. Walker P, C. Whittaker, O. Watson, M. Baguelin, K. Ainslie, S. Bhatia, S. Bhatt, et al. Report 12: The Global Impact of COVID-19 and Strategies for Mitigation and Suppression. 2020 March 26, 2020. Report No.: 12.

29. Ferguson N, D. Laydon, G. Nedjati Gilani, N. Imai, K. Ainslie, M. Baguelin, S. Bhatia, et al.. Report 9: Impact of non-pharmaceutical interventions (NPIs) to reduce COVID-19 mortality and healthcare demand. 2020. Report No.: 9.

30. 2020 WA. WHO ramps up preparedness for novel coronavirus in the African region WHO Regional Office for Africa2020 [January 31, 2020]. Available from: https://www.afro.who.int/news/who-ramps-preparedness-novel-coronavirus-african-region.

31. Aengus Collins M-VFOR. COVID-19 risk governance: drivers, responses and lessons to be learned. Journal of Risk Research. 2020.

32. Goodman PS. Sweden Has Become the World’s Cautionary Tale. The New York Times. 2020 July 7, 2020.

33. Vandell P. Pinal County Sheriff Mark Lamb responds to testing positive for COVID-19, says he wouldn’t enforce mask mandate. AZCentral. 2020 June 20, 2020.

34. Seale H, Heywood AE, Leask J, Sheel M, Thomas S, Durrheim DN, et al. COVID-19 is rapidly changing: Examining public perceptions and behaviors in response to this evolving pandemic. PLoS One. 2020;15(6):e0235112.

35. Fetzer T, Witte, M., Hensel, L., Jachimowicz, J., Haushofer, J., Ivchenko, A., … Yoeli, E. Perceptions of an Insufficient Government Response at the Onset of the COVID-19 Pandemic are Associated with Lower Mental Well-Being. PREPRINT. 2020.

36. De Ver Dye T, Muir E, Farovitch L, Siddiqi S, Sharma S. Critical medical ecology and SARS-COV-2 in the urban environment: a pragmatic, dynamic approach to explaining and planning for research and practice. Infectious Diseases of Poverty. 2020;9(1):1–7.

37. Dye TD, Sy A, Albert P, Cash H, Hadley J, Tomeing T, et al. Critical medical ecological perspectives on diabetes in the Pacific Islands: colonialism, power, and balance in human-environment interaction over time. The Lancet Global Health. 2018;6:S36.

38. Benchimol EI, Smeeth L, Guttmann A, Harron K, Moher D, Petersen I, et al. The REporting of studies Conducted using Observational Routinely-collected health Data (RECORD) statement. PLoS Med. 2015;12(10):e1001885.

39. Tong A, Sainsbury P, Craig J. Consolidated criteria for reporting qualitative research (COREQ): a 32-item checklist for interviews and focus groups. International Journal for Quality in Health Care. 2007;19(6):349–57.

40. Difallah D, Filatova E, Ipeirotis P, editors. Demographics and dynamics of mechanical Turk workers. Proceedings of the Eleventh ACM International Conference on Web Search and Data Mining; 2018.

41. Shaver LG, Khawer A, Yi Y, Aubrey-Bassler K, Etchegary H, Roebothan B, et al. Using Facebook advertising to recruit representative samples: Feasibility assessment of a cross-sectional survey. Journal of Medical Internet Research. 2019;21(8):e14021.

42. United Nations Statistics Division. Methodology–standard country or area codes for statistical use (M49). 2019 [

43. International Organization for Standardization. ISO Country Codes Collection: Online Browsling Platform 2020 [Available from: https://www.iso.org/obp/ui/ - search.

44. Wallston KA, Wallston BS, DeVellis R. Development of the multidimensional health locus of control (MHLC) scales. Health Education & Behavior. 1978;6(1):160–70.

45. Cohen S, Kamarck T, Mermelstein R. Perceived stress scale. Measuring stress: A guide for health and social scientists. 1994;10:1–2.

46. Zimet GD, Dahlem NW, Zimet SG, Farley GK. The multidimensional scale of perceived social support. Journal of Personality Assessment. 1988;52(1):30–41.

47. Andresen EM, Catlin TK, Wyrwich KW, Jackson-Thompson J. Retest reliability of surveillance questions on health related quality of life. Journal of Epidemiology & Community Health. 2003;57(5):339–43.

48. Fahimi M, Link M, Mokdad A, Schwartz DA, Levy P. Tracking chronic disease and risk behavior prevalence as survey participation declines: Statistics from the behavioral risk factor surveillance system and other national surveys. Preventing Chronic Disease. 2008;5(3).

49. Hamel L, Lopez L, Muñana C, Kates J, Michaud J, Brodie M. KFF Health Tracking Poll March 2020: https://www.kff.org/coronavirus-covid-19/poll-finding/kff-coronavirus-poll-march-2020/; 2020 [

50. Streiner DL, Norman GR, Cairney J. Health measurement scales: a practical guide to their development and use: Oxford University Press, USA; 2015.

51. UN Office of the High Representative for the Least Developed Countries. Country Profiles http://unohrlls.org/about-sids/country-profiles/2020 [Available from: http://unohrlls.org/about-sids/country-profiles/.

52. World Health Organization. World health statistics 2016: monitoring health for the SDGs sustainable development goals: World Health Organization; 2016.

53. Dong E, Du H, Gardner L. An interactive web-based dashboard to track COVID-19 in real time. The Lancet infectious diseases. 2020;20(5):533–4.

54. The World Bank. Countries and Economies 2020 [Available from: https://data.worldbank.org/country.

55. Fetters MD, Curry LA, Creswell JW. Achieving integration in mixed methods designs— principles and practices. Health Services Research. 2013;48(6pt2):2134-56.

56. Sibley CG, Greaves LM, Satherley N, Wilson MS, Overall NC, Lee CHJ, et al. Effects of the COVID-19 pandemic and nationwide lockdown on trust, attitudes toward government, and well-being. Am Psychol. 2020;75(5):618–30.

57. Beaunoyer E, Dupere S, Guitton MJ. COVID-19 and digital inequalities: Reciprocal impacts and mitigation strategies. Comput Human Behav. 2020:106424.

58. Bonaccorsi G, Pierri F, Cinelli M, Flori A, Galeazzi A, Porcelli F, et al. Economic and social consequences of human mobility restrictions under COVID-19. Proc Natl Acad Sci U S A. 2020;117(27):15530–5.

59. The International Labour Organization. Nearly half of global workforce at risk as job losses increase due to COVID-19: UN labour agency: United Nations; 2020 [Available from: https://news.un.org/en/story/2020/04/1062792.

60. Douglas M, Katikireddi SV, Taulbut M, McKee M, McCartney G. Mitigating the wider health effects of covid-19 pandemic response. Bmj. 2020;369.

61. Manderson L, Levine S. COVID-19, risk, fear, and fall-out. Taylor & Francis; 2020.

62. Tran PB, Hensing G, Wingfield T, Atkins S, Annerstedt KS, Kazibwe J, et al. Income security during public health emergencies: the COVID-19 poverty trap in Vietnam. BMJ Global health. 2020;5(6):e002504.

63. Che L, Du H, Chan KW. Unequal pain: a sketch of the impact of the Covid-19 pandemic on migrants’ employment in China. Eurasian Geography and Economics. 2020:1–16.

64. Sintema EJ. Effect of COVID-19 on the performance of grade 12 students: Implications for STEM education. Eurasia Journal of Mathematics, Science and Technology Education. 2020;16(7):em1851.

65. Van Lancker W, Parolin Z. COVID-19, school closures, and child poverty: a social crisis in the making. The Lancet Public Health. 2020;5(5):e243–e4.

66. Alvi M, Gupta M. Learning in times of lockdown: how Covid-19 is affecting education and food security in India. Food Security. 2020:1–4.

67. Stevis-Gridneff M. E.U. Formalizes Reopening, Barring Travelers From U.S.: The New York Times; 2020 [Available from: https://www.nytimes.com/2020/06/30/world/europe/eu-reopening-blocks-us-travelers.html.

68. Salcedo A, Sanam Y, G C. Coronavirus Travel Restrictions, Across the Globe: The New York Times; 2020 [Available from: https://www.nytimes.com/article/coronavirus-travel-restrictions.html.

69. Chinazzi M, Davis JT, Ajelli M, Gioannini C, Litvinova M, Merler S, et al. The effect of travel restrictions on the spread of the 2019 novel coronavirus (COVID-19) outbreak. Science. 2020;368(6489):395-400.

70. Ozili PK, Arun T. Spillover of COVID-19: impact on the Global Economy. Available at SSRN 3562570. 2020.

71. Singh S, Kumar R, Panchal R, Tiwari MK. Impact of COVID-19 on logistics systems and disruptions in food supply chain. International Journal of Production Research. 2020:1–16.

72. Inoue H, Todo Y. The propagation of the economic impact through supply chains: The case of a mega-city lockdown against the spread of COVID-19. Available at SSRN 3564898. 2020.

73. Laguna L, Fiszman S, Puerta P, Chaya C, Tárrega A. The impact of COVID-19 lockdown on food priorities. Results from a preliminary study using social media and an online survey with Spanish consumers. Food Quality and Preference. 2020:104028.

74. Nicola M, Alsafi Z, Sohrabi C, Kerwan A, Al-Jabir A, Iosifidis C, et al. The socio-economic implications of the coronavirus pandemic (COVID-19): A review. International journal of surgery (London, England). 2020;78:185.

75. Kirk CP, Rifkin LS. I’ll Trade You Diamonds for Toilet Paper: Consumer Reacting, Coping and Adapting Behaviors in the COVID-19 Pandemic. Journal of Business Research. 2020.

76. Schmidt CW. Lack of Handwashing Access: A Widespread Deficiency in the Age of COVID-19. Environmental Health Perspectives. 2020;128(6):064002.

77. Clemens KS, Matkovic J, Faasse K, Geers AL. Determinants of safety-focused product purchasing in the United States at the beginning of the global COVID-19 pandemic. Safety Science. 2020;130:104894.

78. Lyu W, Wehby GL. Community Use Of Face Masks And COVID-19: Evidence From A Natural Experiment Of State Mandates In The US: Study examines impact on COVID-19 growth rates associated with state government mandates requiring face mask use in public. Health Affairs. 2020:10.1377/hlthaff. 2020.00818.

79. Makino M, Kanie A, Nakajima A, Takebayashi Y. Mental health crisis of Japanese health care workers under COVID-19. Psychological Trauma: Theory, Research, Practice, and Policy. 2020;12(S1):S136.

80. Griffiths MD, Mamun MA. COVID-19 suicidal behavior among couples and suicide pacts: Case study evidence from press reports. Psychiatry Research. 2020.

81. Sotgiu G, Carta G, Suelzu L, Carta D, Migliori G. How to demystify COVID-19 and reduce social stigma. Int J Tuberc Lung Dis. 2020;24(6):640–2.

82. Odriozola-González P, Planchuelo-Gómez Á, Irurtia-Muñiz MJ, de Luis-García R. Psychological symptoms of the outbreak of the COVID-19 crisis and confinement in the population of Spain. 2020.

83. Hartmann-Boyce J, Morris E, Goyder C, Kinton J, Perring J, Nunan D, et al. Diabetes and COVID-19: Risks, Management, and Learnings From Other National Disasters. Diabetes Care. 2020;43(8):1695–703.

84. Rodgers RF, Lombardo C, Cerolini S, Franko DL, Omori M, Fuller-Tyszkiewicz M, et al. The impact of the COVID-19 pandemic on eating disorder risk and symptoms. Int J Eat Disord. 2020;53(7):1166–70.

85. Mohler G, Bertozzi AL, Carter J, Short MB, Sledge D, Tita GE, et al. Impact of social distancing during COVID-19 pandemic on crime in Los Angeles and Indianapolis. J Crim Justice. 2020;68:101692.

86. Campbell AM. An increasing risk of family violence during the Covid-19 pandemic: Strengthening community collaborations to save lives. Forensic Science International: Reports. 2020.

87. Da BL, Im GY, Schiano TD. COVID-19 hangover: a rising tide of alcohol use disorder and alcohol-associated liver disease. Hepatology. 2020.

88. Peçanha T, Goessler KF, Roschel H, Gualano B. Social isolation during the COVID-19 pandemic can increase physical inactivity and the global burden of cardiovascular disease. American Journal of Physiology-Heart and Circulatory Physiology. 2020;318(6):H1441–H6.

89. Patwardhan P. COVID-19: Risk of increase in smoking rates among England’s 6 million smokers and relapse among England’s 11 million ex-smokers. BJGP open. 2020.

90. Nakada LYK, Urban RC. COVID-19 pandemic: Impacts on the air quality during the partial lockdown in Sao Paulo state, Brazil. Sci Total Environ. 2020;730:139087.

91. Ogen Y. Assessing nitrogen dioxide (NO2) levels as a contributing factor to the coronavirus (COVID-19) fatality rate. Science of The Total Environment. 2020:138605.

92. Pinder AC, Raghavan R, Britton JR, Cooke S. COVID-19 and biodiversity: The paradox of cleaner rivers and elevated extinction risk to iconic fish species. Aquatic Conservation: Marine and Freshwater Ecosystems. 2020;30(6):1061–2.

93. Velásquez RMA, Lara JVM. Gaussian approach for probability and correlation between the number of COVID-19 cases and the air pollution in Lima. Urban Climate. 2020:100664.

94. Matache M, Bhabha J. Anti-Roma Racism is Spiraling During COVID-19 Pandemic. Health and Human Rights. 2020;22(1):379.

95. Meneses-Navarro S, Freyermuth-Enciso MG, Pelcastre-Villafuerte BE, Campos-Navarro R, Meléndez-Navarro DM, Gómez-Flores-Ramos L. The challenges facing indigenous communities in Latin America as they confront the COVID-19 pandemic. International Journal for Equity in Health. 2020;19:1–3.

96. Kluge HHP, Jakab Z, Bartovic J, D’Anna V, Severoni S. Refugee and migrant health in the COVID-19 response. The Lancet. 2020;395(10232):1237–9.

97. Hooper MW, Nápoles AM, Pérez-Stable EJ. COVID-19 and racial/ethnic disparities. Jama. 2020.

